# Unveiling the Landscape of Reportable Genetic Secondary Findings in the Spanish Population: A Comprehensive Analysis Using the Collaborative Spanish Variant Server Database

**DOI:** 10.1101/2024.08.01.24311343

**Authors:** Rosario Carmona, Javier Pérez-Florido, Gema Roldán, Carlos Loucera, Virginia Aquino, Noemí Toro-Barrios, José L. Fernández-Rueda, Gerrit Bostlemann, Daniel López-López, Francisco M. Ortuño, Beatriz Morte, María Peña-Chilet, Joaquín Dopazo, CSVS Crowdsourcing Group

**Affiliations:** Andalusian Platform for Computational Medicine, Andalusian Public Foundation Progress and Health-FPS, 41013, Seville, Spain; Centro de Investigación Biomédica en Red en Enfermedades Raras (CIBERER), ISCIII, Madrid, Spain; Institute of Biomedicine of Seville, IBiS, University Hospital Virgen del Rocío/CSIC/University of Seville, Seville, Spain; Departamento de Ingeniería de Computadores, Automática y Robótica, University of Granada, 18071, Granada, Spain; Platform for Big Data, IA and Biostatistics, La Fe Health Research Institute, 46026, Valencia, Spain; Navarrabiomed-IdiSNA, Complejo Hospitalario de Navarra, Universidad Pública de Navarra (UPNA), IdiSNA (Navarra Institute for Health Research), 31008, Pamplona, Spain; Department of Genetics, Instituto de Investigación Sanitaria-Fundación Jiménez Díaz University Hospital, Universidad Autónoma de Madrid (IIS-FJD, UAM), 28040, Madrid, Spain; Fundación para la Investigación y Docencia Sant Joan de Deu, 08830, Barcelona, Spain; Department of Maternofetal Medicine, Genetics and Reproduction, Institute of Biomedicine of Seville (IBIS), University Hospital Virgen del Rocío/CSIC/University of Seville, 41013, Seville, Spain; Fundación Pública Galega de Medicina Xenómica, SERGAS, IDIS, 15706, Santiago de Compostela, Spain; Asociación Instituto de Investigación Sanitaria de Biocruces, Vizcaya, 48903, Vizcaya, Spain; Hospital Univ. 12 de Octubre, 28041, Madrid, Spain; Centro de Investigación Príncipe Felipe, 46012, Valencia, Spain; Universidad de Barcelona, 08193, Barcelona, Spain; Hospital Virgen de la Arrixaca, 30120, Murcia, Spain; Center for Biomedical Network Research on Rare Diseases (CIBERER), ISCIII, 28040, Madrid, Spain; Otology & Neurotology Group CTS 495, Department of Genomic Medicine, Centre for Genomics and Oncological Research (GENYO), Pfizer University of Granada, 18016, Granada, Spain; Vall d’Hebron Institut de Recerca, 08035, Barcelona, Spain; Fundación para la Investigación del Hospital la Fe, 46026, Valencia, Spain; Servicio de Genética, Ramón y Cajal Institute of Health Research (IRYCIS), 28034, Madrid, Spain; Undiagnosed Rare Diseases Programme (ENoD). Center for Biomedical Research on Rare Diseases (CIBERER), ISCIII, 28029, Madrid, Spain; Universidad de Murcia, 30100, Murcia, Spain; Fundación IDIBELL, 08005, Barcelona, Spain; Fundación para la Investigación y Docencia Sant Joan de Deu, 8830, Barcelona, Spain; Universidad Autónoma de Madrid, 28049, Madrid, Spain; Agencia Estatal Consejo Superior de Investigaciones Científcas, 28006, Madrid, Spain; Universidad de Zaragoza, 50009, Zaragoza, Spain; Hospital Clínico y Provincial de Barcelona, 08036, Barcelona, Spain; Fundación Instituto de Investigación Sanitaria Illes Baleares (IdISBa), 07120, Palma de Mallorca, Spain

**Keywords:** secondary findings, local population, genomes, pathogenic variants, ACGM recommendations

## Abstract

The escalating adoption of Next Generation Sequencing (NGS) in clinical diagnostics reveals genetic variations, termed secondary findings (SFs), with health implications beyond primary diagnoses. The Collaborative Spanish Variant Server (CSVS), a crowdsourced database, contains genomic data from more than 2100 unrelated Spanish individuals. Following the American College of Medical genetics (ACMG) guidelines, CSVS was analyzed, identifying pathogenic or likely pathogenic variants in 78 actionable genes (ACMG list v3.1) to ascertain SF prevalence in the Spanish population. Among 1129 samples, 60 reportable SFs were found in 5% of individuals, impacting 32 ACMG-listed genes, notably associated with cardiovascular disease (59.4%), cancer (25%), inborn errors of metabolism (6.3%), and other miscellaneous phenotypes (9.4%). The study emphasizes utilizing dynamic population databases for periodic SF assessment, aligning with evolving ACMG recommendations. These findings illuminate the prevalence of significant genetic variants, enriching understanding of secondary findings in the Spanish population.

## Background

In contemporary clinical practice, the utilization of whole genome sequencing (WGS), whole exome sequencing (WES), and clinical exome sequencing (CES) has become increasingly commonplace. These techniques play a pivotal role in the accurate diagnosis of hereditary and rare diseases [1] as well as in providing tailored treatment recommendations for cancer patients [2]. Furthermore, the availability of an individual’s genetic sequence not only facilitates targeted disease diagnosis and treatment but also presents the intriguing opportunity to uncover pathogenic variants unrelated to the primary reason for medical consultation. These findings, detected in opportunistic screening, known as secondary findings (SF), hold substantial clinical significance. The identification of SF relies on patient consent, as the informed disclosure of unexpected genetic insights contributes to a more comprehensive understanding of an individual’s health profile, potentially offering opportunities for proactive interventions and improved patient care.

The American College of Medical Genetics (ACMG) has periodically issued guidance and updates concerning the reporting of secondary findings within the realm of clinical genome and exome sequencing [3]. Notably, the ACMG outlines a list of genes linked to highly penetrant diseases for which actionable treatments and prevention guidelines are available, aiming to curtail mortality and morbidity in genetically predisposed individuals. Over time, this list of actionable genes recommended for secondary findings exploration has expanded, with the most recent iteration at the time of this study (ACMG v3.1 list) encompassing 78 such genes [3]. While the ACMG’s approach seeks to harness genetic insights to inform medical intervention, the European Society of Human Genetics (ESHG) takes in contrast a more cautious stance, advocating for a conservative approach towards opportunistic screening and advising the minimization of unsolicited findings [4].

Several studies have been published that adhere to ACMG recommendations and engage in the analysis of SFs across diverse populations (refer to Table 1). The initial studies assessed SFs predicated on the ACMG 1.0 list, which encompassed 56 genes [5] and are detailed below. An examination of SFs within the 1,902 unrelated individuals of all the world, corresponding to Phase 1 of the 1000 Genomes dataset unveiled a prevalence of 1% [6]. Most of the findings were found in European and American individuals, while there were few in groups of African or Asian ancestry. Subsequently, a unique exome-based study deduced a prevalence rate of 2% in a cohort composed of 4,300 individuals of European ancestry, and 1.1% within a subset of 2,203 African individuals in the analysis of 112 medically actionable genes [7], while if focusing only on pathogenic variants in the gene subset of the ACMG 1.0 list, these prevalences were 0.7% in the European cohort and 0.5% in the African group. They state that this lower rate of pathogenic variants in the African ancestry group could be due to the underrepresentation of this population in research and clinical studies. A broader initiative was carried out in the DiscovEHR study, which combined next-generation sequencing with electronic health record data, with the analysis of 50,726 whole exome sequencing (WES) profiles, in search of deleterious variants in genes clinically actionable from the ACMG 1.0 list. They estimated that 3.5% of individuals had pathogenic or probably pathogenic variants suitable for clinical action [8]. In parallel, a study encompassing 377 Southeast Asian individuals, employing either WES or whole genome sequencing (WGS), yielded a SFs rate of 1.6% [9]. Lastly, a consolidated study in individuals that had not been selected for family history of disease probed SFs, revealing a prevalence of 1.1% within 462 WES profiles of European American individuals, and a corresponding 1% rate among 3223 African-American profiles [10]. This work took advantage of related clinical characteristics determined from the participants’ medical history and demonstrated an increased aggregate risk of developing clinical characteristics associated with the corresponding diseases.

**Table 1.** Summary of published secondary findings studies.

| ACMG SF List Version | Number of genes | Year of publication | Ethnicity or origin | Cohort size | Secondary Findings rate (%) | Sequencing Technology* | Reference |
| --- | --- | --- | --- | --- | --- | --- | --- |
| 1.0 | 56 | 2015 | 1000 Genomes (mix) | 1092 | 1 | WGS | [6] |
| 1.0 | 56 | 2015 | European | 4300 | 0.7 | WES | [7] |
| 1.0 | 56 | 2015 | African | 2203 | 0.5 | WES | [7] |
| 1.0 | 56 | 2016 | Mix | 50726 | 3.5 | WES | [8] |
| 1.0 | 56 | 2016 | South East Asia | 377 | 1.6 | WES / WGS | [9] |
| 1.0 | 56 | 2018 | European American | 462 | 1.1 | WES | [10] |
| 1.0 | 56 | 2018 | African American | 3223 | 1 | WES | [10] |
| 2.0 | 59 | 2017 | East Asian | 959 | 2.5 | WGS | [12] |
| 2.0 | 59 | 2018 | Chinese | 421 | 2.85 | WES | [13] |
| 2.0 | 59 | 2019 | Dutch | 1640 | 2.7 | WES | [14] |
| 2.0 | 59 | 2020 | Hong kong Chinese | 1116 | 1.52 | WES | [15] |
| 2.0 | 59 | 2020 | UK Biobank (mix) | 49960 | 2 | WES | [16] |
| 2.0 | 59 | 2020 | Taiwanese | 161 | 1.86 | WES | [17] |
| 2.0 | 59 | 2020 | Lebanese | 280 | 8.6 | WES | [18] |
| 2.0 | 59 | 2020 | Mix | 21915 | 2.54 | PANEL | [19] |
| 2.0 | 59 | 2021 | Turkish | 622 | 2.1 | CES | [20] |
| 2.0 | 59 | 2021 | Saudi Arabia | 1254 | 8 | WES | [21] |
| 2.0 | 59 | 2021 | Qatar (mix) | 6045 | 2.3 | WGS | [22] |
| 2.0 | 59 | 2022 | Chinese | 2020 | 3.02 | CES | [24] |
| 2.0 | 59 | 2022 | Colombian | 160 | 8.13 | WES | [23] |
| 2.0 | 59 | 2022 | Spanish | 740 | 3.6 | WES / WGS | [25] |
| 3.0 | 73 | 2022 | Thai | 1559 | 5.5 | WES | [27] |
| 3.0 | 73 | 2022 | Chinese | 2987 | 5.3 | WES | [28] |
| 3.1 | 78 | 2023 | Pakistani | 863 | 2.7 | WGS | [29] |

\*WGS: whole genome sequencing; WES: whole exome sequencing; CES: clinical exome sequencing

Most of the published studies have focused on the analysis of SFs utilizing the ACMG v2.0 list, encompassing 59 genes [11]. Among them, various SF prevalence rates have been reported. One of them refined the previously published rate of secondary findings on East Asians through a large-scale WGS study, showing that WGS provides broader coverage of the 59 actionable genes than WES. They determined a prevalence of 2.5% in 954 whole genome sequencing (WGS) profiles from East Asian populations and described genetic heterogeneity by ethnicity [12]; an assessment of 421 whole exome sequencing (WES) profiles from Chinese children examined how racial diversity influences the identification of secondary findings unveiling a SF prevalence of 2.85% [13]. The participants in the latter study were young individuals, so some phenotypes of a future disease may not have emerged yet (adult or age of onset), which could lead to misidentifying variants responsible for the disease as secondary findings, so special attention must be paid when reporting SFs in children or young patients. An analogous SF rate of 2.7% was observed within a cohort of 1640 Dutch WES profiles of anonymized healthy Dutch individuals, corresponding to 44 subjects carrying a medically actionable dominant variant in any of the 59 genes in the ACMG v2.0 list [14]. They also identified 36 individuals (2.2%) as carrying a high-risk disease allele in actionable recessive genes, but not reportable, as none of them were homozygous or compound heterozygous for these alleles and ACMG recommends returning only pathogenic variants. biallelic, so they do not count towards the rate of SFs. Another study involved a screening of 1116 Hong Kong Chinese WES profiles that yielded a frequency of 1.52% for SFs and identified some pathogenic or likely pathogenic variants as founder mutations or recurrent mutations in Asians or Chinese [15]. The largest study to our knowledge involving 49960 individual WES profiles, of European genetic ancestry (93.6%), but also Africans (1.49%) and East Asians (0.54%), from the UK Biobank conveyed a SF prevalence rate of 2% [16]. A SF rate of 1.86% was observed within 161 Taiwanese WES profiles, emphasizing the high carrier rate of pathogenic and probably pathogenic variants in the ATP7B gene, which coincides with the higher prevalence of Wilson’s disease described in the East Asian population [17]. A cohort of 280 Lebanese WES profiles presenting a wide spectrum of genetic disorders were examined and, in addition to molecular diagnosis, were screened for SFs and showed a rate of 8.6% [18]. In a collective of 21915 eMERGE network participants representing diverse ethnic backgrounds, a SF prevalence of 2.54% was ascertained using panels [19]. A study encompassing 622 Turkish clinical exome sequencing (CES) profiles identified a rate of 2.1% for reportable variations [20]. The investigation into the Saudi population disclosed a SF rate of 8% across 1254 WES profiles [21]. According to the authors, this higher rate of SF compared to other studies could be due to the identification of new Saudi pathogenic or probably pathogenic variants never reported in other populations. Another study focused on the analysis of SF in Arab and other Middle Eastern populations involved the examination of 6045 participants from the Qatar Genome Program unveiling a SF prevalence rate of 2.3% [22]. Also, recent studies revealed a SF rate of 8.13% within 160 Colombian WES profiles [23]. In the China Neonatal Genome Project, a total of 2020 CES profiles were analyzed for secondary finding variants from the ACMG v2.0 list and revealed a SF rate of 3.02% [24]. More recently, a Spanish study documented a SF prevalence rate of 3.6% within a cohort primarily consisting of 740 WES profiles [25].

As far as our knowledge extends, the ACMG v3.0 list of 73 genes [26] has been utilized by only two studies for the purpose of reporting SFs. One study examined 1,559 Thai exomes, revealing a SF rate of 5.5%, most of them related to cardiovascular and cancer phenotypes [27], while another analyzed a Chinese cohort of 2,987 individuals, and a SF prevalence of 5.3% was reported [28]. Finally, a very recent study has been published that employed the ACMG v3.1 list, encompassing 78 genes [3], focusing on the pursuit of SFs within 863 Pakistani individuals. In this study, a SF rate of 2.7% was reported [29]. Interestingly, an unexpectedly high proportion of biallelic variants was observed, consistent with the high level of consanguinity in this population.

Reporting SFs in these actionable genes imparts valuable information to the individuals and their families, thereby enabling potential preventive interventions, as well as timely and appropriate clinical monitoring and disease management. However, this prevalence, as illustrated in the results reported above, displays significant differences depending on ethnicity and geographic region [30–32]. Also sequencing technologies and cohort sizes can impact the results. Country-level analysis of SFs would ideally capture the genetic background of the country population, avoiding biases caused by subsamples from hospital-level studies or the dilution of local variants observed in multi-country studies. The Collaborative Spanish Variant Server (CSVS) has been established as a crowdsourcing initiative aimed at furnishing the scientific and medical community with information pertaining to the genomic variability of the Spanish population [31]. CSVS encompasses over 2100 exomes and genomes of unrelated individuals of Spanish ancestry, evenly distributed across various geographical regions of Spain. Consequently, the availability of a country-level, population-specific database for the investigation of SFs presents itself as a valuable instrument to direct healthcare policy endeavors. This enables the formulation of preventive and therapeutic approaches tailored to the predominant diseases prevalent within the specific country or region.

The objective of this study resides in harnessing the latent potential of a country-scale database of genomic variation to assess the prevalence of pathogenic variants and to ascertain secondary findings within the Spanish population. As an additional objective, the availability of prevalence values of variants that could be considered SF in some patients or individuals is another relevant practical asset of the CSVS. Given that the criteria set forth by the ACGM for classifying a variant as pathogenic or likely pathogenic are contingent not solely on the isolated variant but rather on its interaction with other variants, including their prevalence within a population, the incorporation of this metric within the CSVS markedly enhances its clinical utility.

## Methods

### Data

The Collaborative Spanish Variant Server (CSVS) is a crowdsourcing initiative to provide information about the genomic variability of the Spanish population to the scientific and medical community [31]. CSVS contains genomic data from different projects such as the Medical Genome Project [30, 33], the EnoD, (Undiagnosed Rare Diseases programme), the Spanish Network for Research in Rare Diseases (CIBERER), the Project Genome 1000 Navarra, The RareGenomics from Madrid, and other research groups and initiatives across Spain [34–36]. Each submitter is responsible for obtaining the informed consent from each participant submitted to CSVS.

Currently, CSVS stores information (WES or WGS) on 2105 unrelated Spanish individuals (version 5.0.0) and will continue to increase with new contributions of samples. Among these, 1129 individuals pertain to discrete samples, comprising 666 exomes and 463 genomes, while the remaining 976 individuals stem from aggregate samples. The samples were sequenced using Illumina technology in different sequencing facilities. Apart from the aggregate samples, that were processed in origin and submitted to CSVS, the rest of samples underwent a common processing pipeline [37] (see details in the original CSVS publication [31]).

### Genes and individuals analyzed

In this study, the CSVS population was surveyed to assess the occurrence of pathogenic variants and secondary findings within the Spanish population, adhering to the criteria set forth by the ACMG (American College of Medical Genetics). At the time this study was carried out, the most recent ACMG list (v3.1) suggested the scrutiny of 78 actionable genes [3], a compilation that offers insights into several distinct disease categories, including cancer, cardiovascular phenotypes, congenital metabolic disorders, and other related conditions, providing a framework for understanding the genetic underpinnings of a broad range of health conditions.

### Potential secondary findings variants

All variants present in the 78 ACMG genes in CSVS and having allele frequency (MAF) under 0.1 in 1KG-EUR and 1KG-ALL, as well as in Gnomad v2.1.1 [38], were recovered and annotated with ClinVar database [39] (version 20221027). All pathogenic or likely pathogenic variants according to ClinVar were registered. Besides, variants with conflicting interpretations of pathogenicity in ClinVar with at least one interpretation as pathogenic or likely pathogenic were classified according to ACMG guidelines using InterVar [40] with the default values and only those finally labeled as pathogenic or likely pathogenic were accounted for. In addition, variants not annotated in ClinVar and impacting coding regions through mechanisms such as the introduction of stop codons, loss or alteration of stop/start codons, modifications in splice acceptor/donor sequences, or induction of frameshift mutations, were subjected to interpretation using InterVar. Variants classified as pathogenic (P) or likely pathogenic (LP) by InterVar were additionally considered, expanding the scope of variant analysis. For each variant, only healthy individuals, or individuals with ICD10 categories non-overlapping with phenotypes corresponding to the variant were considered [31].

The identification of secondary findings within CSVS adhered to the established criteria outlined by the ACMG [3].

The estimation of carriers bearing Pathogenic or Likely Pathogenic variants on a per-variant basis is carried out using all the individuals in CSVS (individual and aggregated data). Since aggregated samples do not provide information on variant heterozygosity, only individual samples were used to estimate rates of potential SF at the population level.

## Results

### Identification of potential secondary findings variants

Inclusive of both individual and aggregate samples, a total of 90,654 variants encompassing the compendium of 78 ACMG recommended genes was identified within the complete CSVS database (see Figure 1). These variants underwent a frequency-based filtering process and were subjected to comprehensive analysis via reference to the ClinVar database and ACMG’s classification guidelines, as delineated in the methodology section. Following this evaluation, a total of 541 variants emerged as potential Secondary Findings (SFs). This subset is delineated as follows: 75 variants were ascribed pathogenic or likely pathogenic designations according to ClinVar annotations, while 14 variants exhibited conflicting interpretations within ClinVar yet aligned with pathogenic or likely pathogenic categorizations guided by ACMG criteria. Additionally, 452 variants, unaccounted for in ClinVar annotations, were deemed pathogenic or likely pathogenic based on ACMG assessments due to their propensity to induce protein truncation or modify splice sites (see Supplementary Table 1 for details).

**Figure 1.**
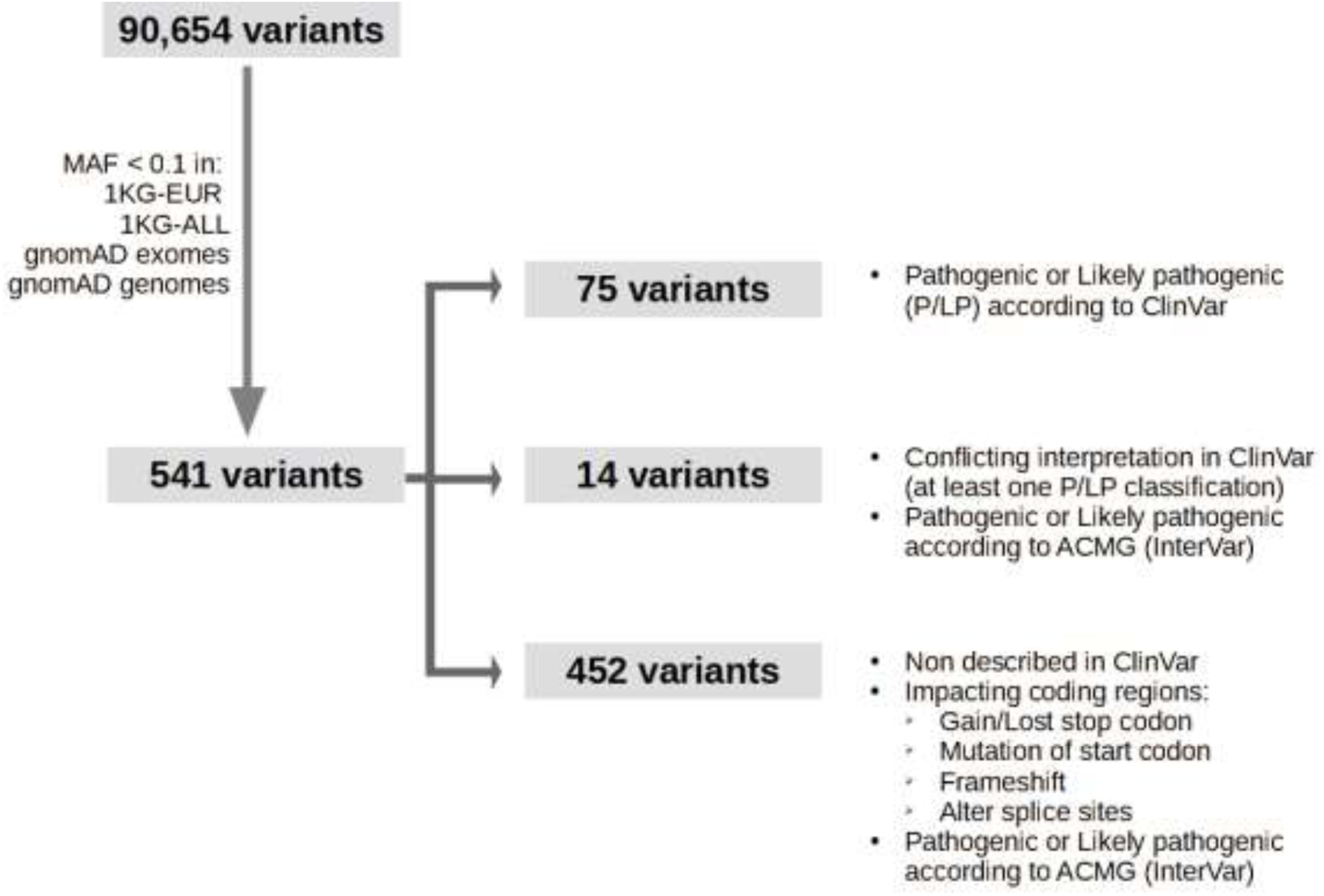
Summary diagram of the variant filtering process. P: pathogenic. LP: likely pathogenic.

### Variant-based analysis of potential secondary findings variants

Considering all the samples, both the individual sample dataset and the aggregated dataset, this ensemble of 541 variants were observed in 66 out of the 78 genes delineated by ACMG as pivotal for consideration. A comprehensive depiction of the count of Pathogenic or Likely Pathogenic variants attributed to each gene, along with the corresponding associated conditions, is provided in Supplementary Table 2. Notably, several genes exhibited an accumulation of variants, with the most prominent instances including titin (*TTN*) with 125 variants (constituting 23% of the total), followed by breast cancer genes 1 and 2 (*BRCA1* and *BRCA2*) harboring 20 (3.7%) and 66 (12.2%) variants, respectively. Furthermore, ryanodine receptors 1 and 2 (*RYR1* and *RYR2*) were also notable, presenting 18 (3.3%) and 22 (4%) variants, respectively.

While the majority of variants were observed in a heterozygous state within carrier individuals, specific exceptions were observed: *TTN* c.22148_22149insAG (p.Asn7383fs) occurred in a homozygous state within a unique individual, GLA c.427G>A (p.Ala143Thr, rs104894845) manifested in a hemizygous state within an individual, and the distinct instance of the HFE c.845G>A (p.Cys282Tyr, rs1800562) variant displayed a notable presence of heterozygous carriers (110 out of 2078, accounting for 5.39%) alongside two homozygous carriers (see Supplementary Table 1 for further information).

### Analysis of potential secondary findings at the level of individual samples

Considering the cohort of 1129 individual samples, a total of 105 (9.3%) individuals exhibited the presence of one or more of the 77 designated pathogenic or likely pathogenic variants. Of this total, 60 variants were identified as reportable secondary findings (refer to Table 2), while the remaining 17 variants did not qualify for reporting due to their heterozygous nature within genes associated with autosomal disorders, without the simultaneous presence of a second variant to elucidate a compound heterozygous state. Within the CSVS dataset, out of the 1129 individual samples, 57 instances (5%) showcased at least one reportable secondary finding variant. Among these individuals, the majority (53 individuals, 93%) manifested a singular secondary finding variant, with two variants constituting the most prominent observation. Notably, no secondary finding variant was found in more than one individual, except for SCN5A c.4671del (p.Asn1557fs), which was shared between two unrelated individuals.

**Table 2.**
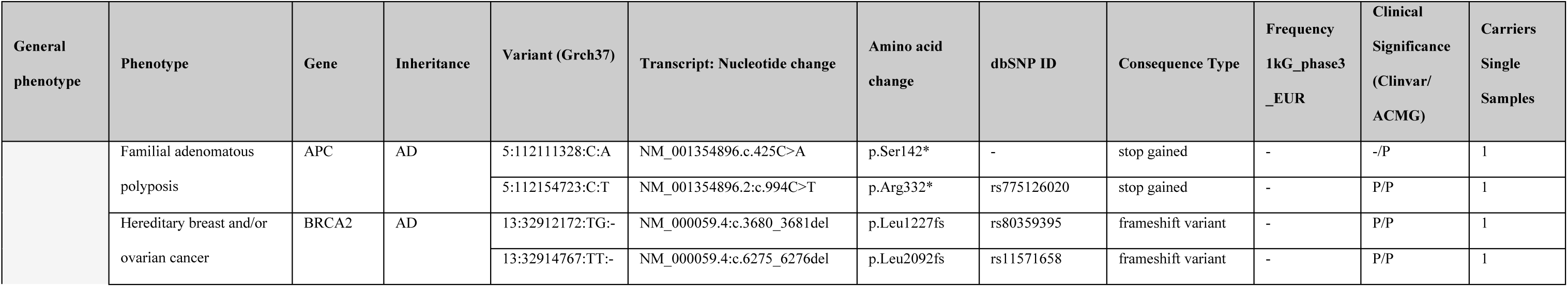
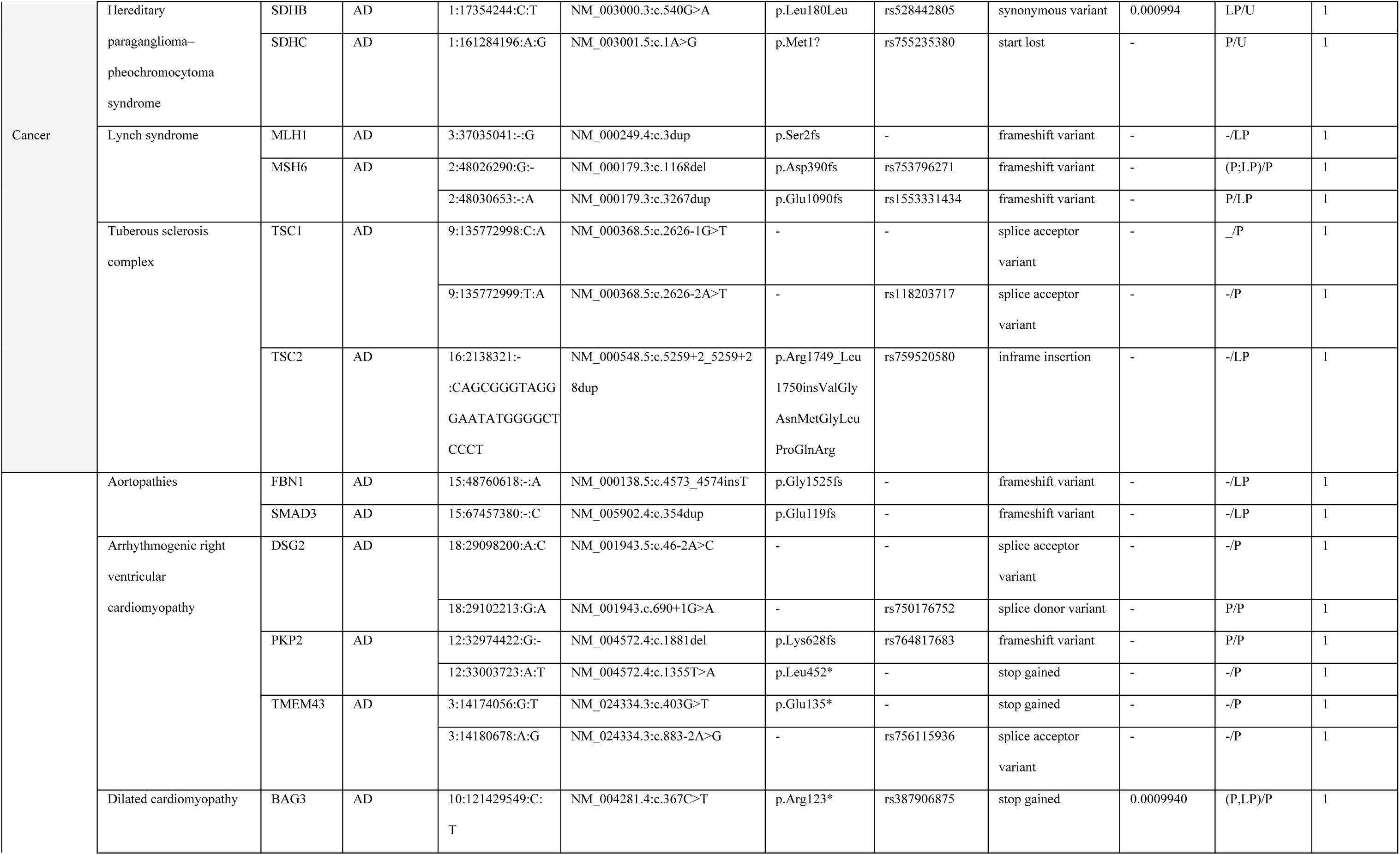
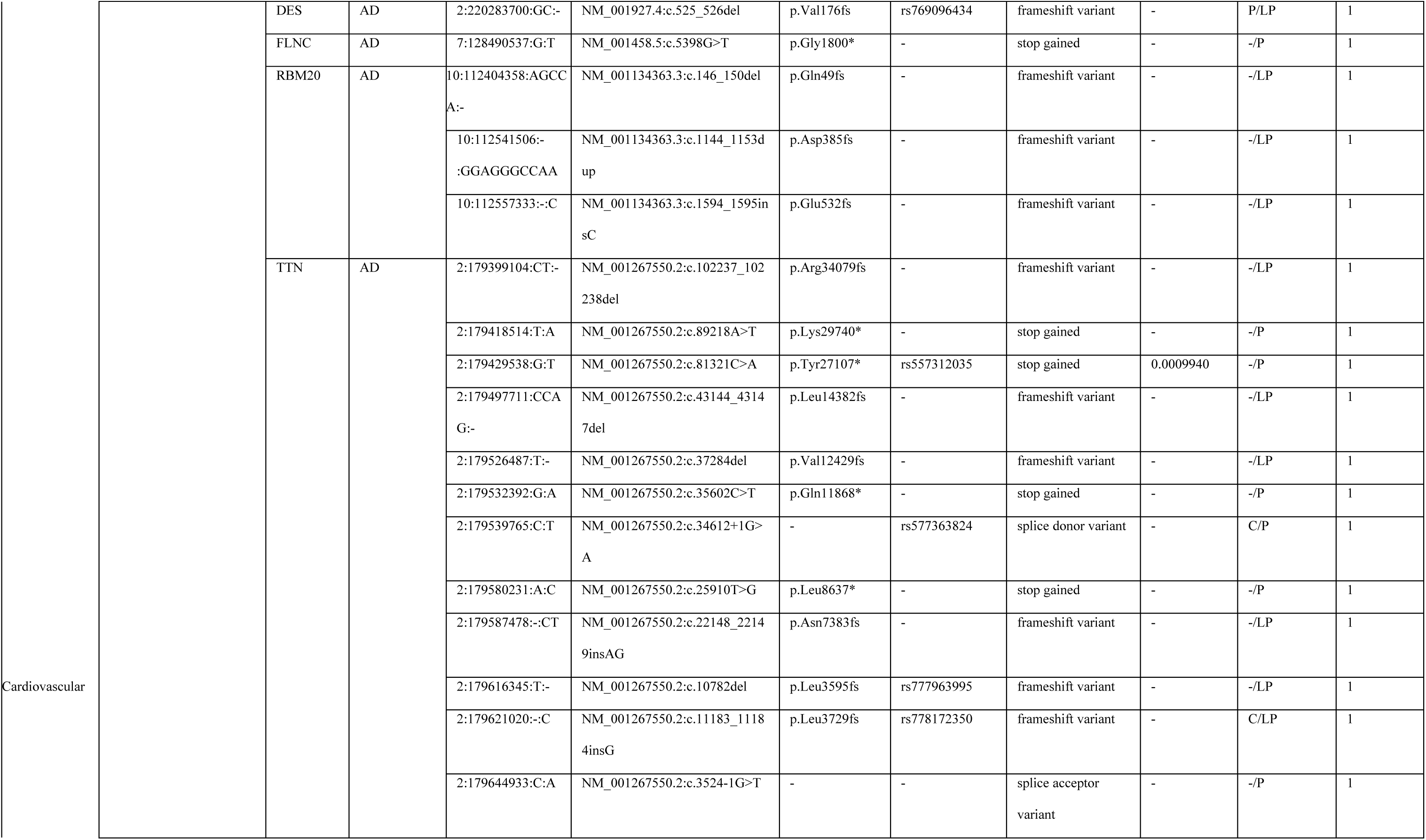
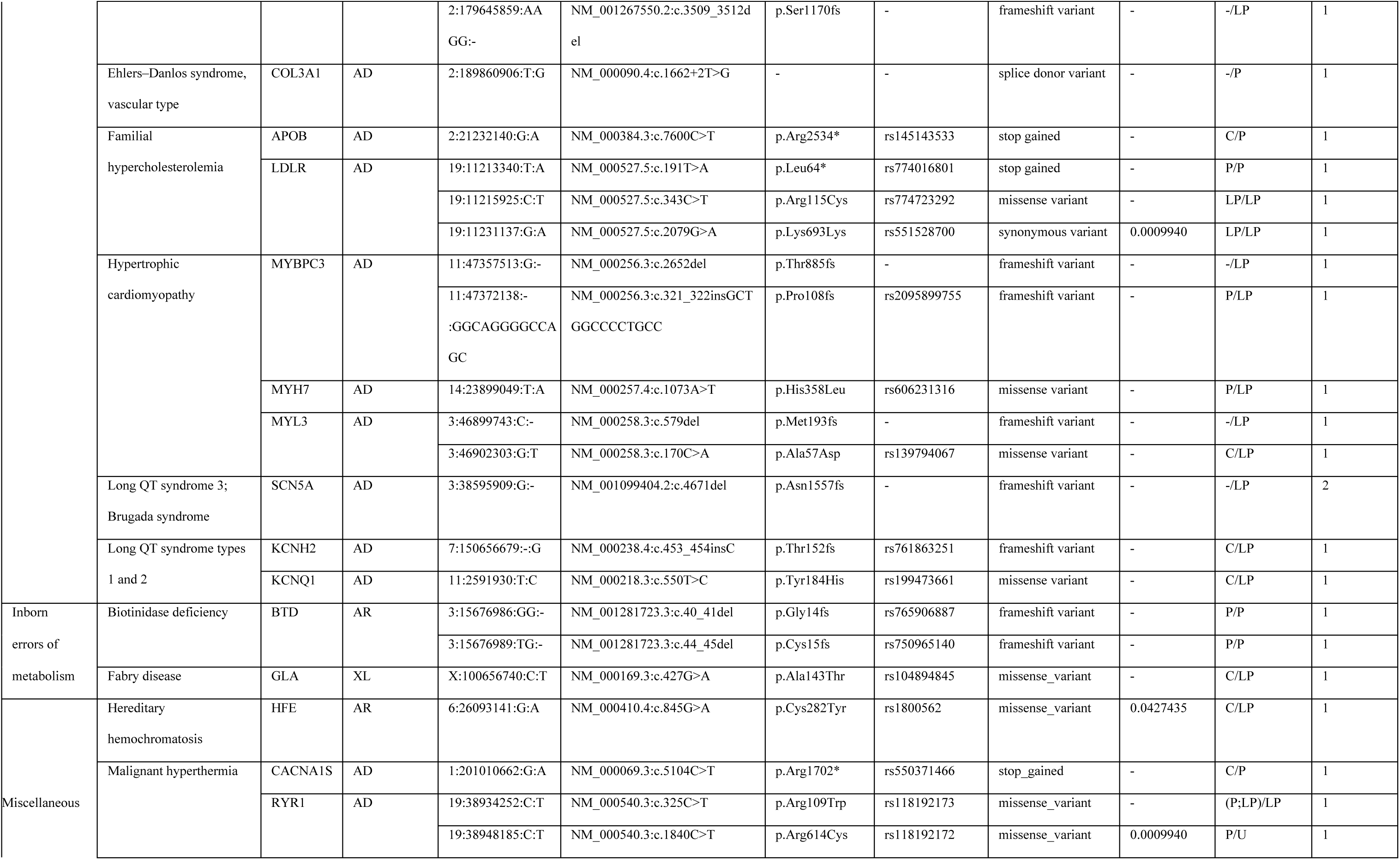
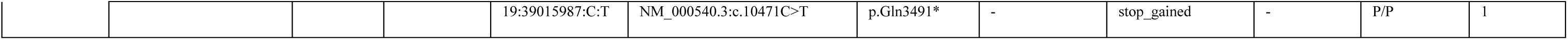
Secondary findings variants in 78 ACMG recommended genes (v3.1) detected in individual samples of CSVS database. Columns are as follows: General phenotype aggregates general categories listed in the next column. General Phenotype category, are General categories of diseases defined by ACMG for reporting secondary findings [3]. Phenotype, are General phenotype associated with the gene. Gene, list the gene in which the variant is located. Inheritance, define the inheritance mode for the disease (AD, autosomal dominant; AR, autosomal recessive; SD, semidominant; XL, X-linked). Variant (Grch37), in format “chromosome : position : reference allele : alternative allele”, according to genome reference Grch37. Nucleotide change contains change at DNA level according to HGVS nomenclature referred to the canonical transcript in RefSeq. Amino acid change, lists change at protein level according to HGVS nomenclature. dbSNP ID, contains variant identifier in dbSNP database (rs). Consequence Type, lists the worst consequence type found among all transcripts. Frequency 1kG_phase3_EUR, contains the frequency recorded in 1000G for the variant selected for the European population. ClinicalSignificance(Clinvar/ACMG), contains the clinical significance of the variant according to both. ClinVar database and ACMG classification (P=pathogenic, LP=likely pathogenic). Carriers Single Samples, contains the number of individuals carrying the variant considering only individual samples.

The encompassing spectrum of these secondary findings spanned 32 of the 78 genes proposed by ACMG (see Figure 2). These genes included various phenotypic categories, with 19 associated with cardiovascular phenotypes (such as *FBN1*, *SMAD3*, *DSG2*, *PKP2, TMEM43, BAG3, DES, FLNC, RBM20, TTN, COL3A1, APOB, LDLR, MYBPC3, MYH7, MYL3, SCN5A, KCNH2* and *KCNQ1*), 8 related to cancer (*APC, BRCA2, SDHB, SDHC, MLH1, MSH6, TSC1* and *TSC2*), 2 linked to inborn errors of metabolism (*BTD* and *GLA*), and 3 attributed to diverse phenotypes (such as *CACNA1S* and *RYR1* associated with malignant hyperthermia and *HFE* responsible for hereditary hemochromatosis). These genes collectively underlie 17 distinct disorders, characterized by 14 instances of autosomal dominant inheritance patterns, 2 autosomal recessive diseases (biotinidase deficiency and hereditary hemochromatosis), and 1 X-linked disorder (Fabry disease).

**Figure 2.**
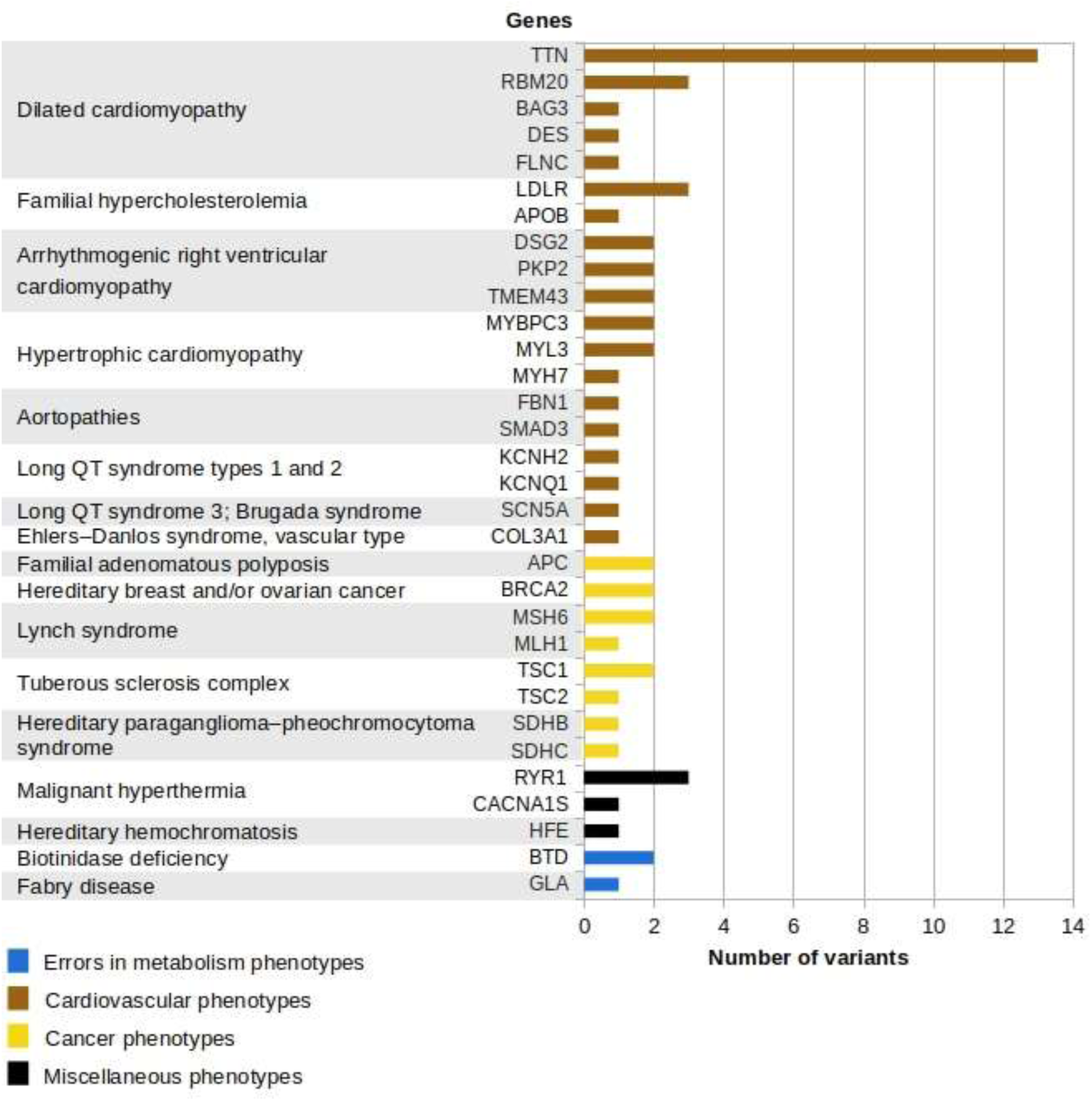
Schematic representation of distribution of the 60 variants identified as actionable secondary findings in 1129 individual samples of CSVS.

With the exception of specific cases, most secondary findings were observed in a heterozygous state. Notably, *TTN* c.22148_22149insAG (p.Asn7383fs) presented as homozygous in a single individual, *GLA* c.427G>A (p.Ala143Thr, rs104894845) emerged as hemizygous in another individual, and the variant *HFE* c.845G>A (p.Cys282Tyr, rs1800562) exhibited homozygosity in one individual. An intriguing instance of potential compound heterozygosity appeared, where one individual carried two distinct pathogenic heterozygous variants in the biotinidase gene *BTD*: c.40_41del (p.Gly14fs, rs765906887) and c.44_45del (p.Cys15fs, rs750965140), associated with biotinidase deficiency, a recessive disorder.

### Information on secondary findings is publicly available and updated on regular basis in CSVS

The insights garnered concerning secondary findings within the Spanish population are accessible through the CSVS web interface [31]. This repository designates variants that meet the criteria outlined in the methodology presented in this study, conforming to the ACMG guidelines pertaining to the 78 specified actionable genes (ACMG list v3.1). For each of these identified variants, the CSVS platform also furnishes pertinent information including the linked phenotype, the OMIM disorder terminology corresponding to the relevant gene, and the inheritance pattern associated with the implicated disease. As new genomic data enter in CSVS the information on secondary findings is updated. This guarantees an increasingly precise survey of the secondary finding in the Spanish population.

## Discussion

In this study, CSVS was analyzed for secondary findings in 78 actionable genes (ACMG v3.1). The prevalence of pathogenic or likely pathogenic variants at individual and population levels was assessed. The carrier prevalence for 541 variants, cataloged within the curated list of 78 ACMG genes, as referenced by ClinVar and/or ACMG criteria (refer to Supplementary Table 1 for details), ranged from 0.048% to 5.39% within the CSVS dataset. Comprehensive analysis revealed prevalent variants, including a subset of 14 exceeding 1% frequency. This exploration provided insights into the collective occurrence of secondary findings in the Spanish population.

By far, the most prevailing pathogenic variant within the dataset was the *HFE* c.845G>A (p.Cys282Tyr, rs1800562) allele, observed in 112 out of 2078 individuals (constituting 5.39% of the cohort). Among these instances, 110 individuals exhibited heterozygosity, while 2 individuals manifested homozygosity. The *HFE* gene, governing homeostatic iron regulation, is linked to hereditary hemochromatosis [41], an autosomal recessive disorder characterized by anomalous iron metabolism, resulting in the accumulation of surplus iron within diverse organs, thereby precipitating their malfunction. The notable prevalence of the *HFE* c.845G>A allele within our cohort aligns harmoniously with prior investigations, which have highlighted its prominent presence, particularly among individuals of European ancestry [42].

Among the variants exceeding a 1% prevalence rate, a mere two remain cataloged within the dbSNP database and were classified as pathogenic as per both ClinVar and ACMG standards: *MUTYH* c.1103G>A (p.Gly368Asp, rs36053993) and *ATP7B* c.1934T>G (p.Met645Arg, rs121907998). The mutY DNA glycosylase gene (*MUTYH*) is linked to autosomal recessive colorectal adenomatous polyposis [43], and the ATPase copper transporting beta gene (*ATP7B*) underpins Wilson disease [44]. Interestingly, for the remaining variants boasting a prevalence surpassing 1% within the Spanish population, no frequency data has been described in population databases (dbSNP or ClinVar). Remarkably, this subset consists exclusively of frameshift variants, all classified as Pathogenic or Likely Pathogenic in accordance with ACMG criteria. These variants exert their impact on the following genes: three are linked to cardiovascular phenotypes—titin (*TTN*) associated with dilated cardiomyopathy [45], desmoplakin (*DSP*) associated with arrhythmogenic right ventricular cardiomyopathy [46], and ryanodine receptors 2 (*RYR2*) associated with Catecholaminergic polymorphic ventricular tachycardia [47]. Notably, the fourth gene in this category is *BRCA2*, pertinently associated with hereditary breast and/or ovarian cancer. The notable contrast between their relatively high prevalence within CSVS and their conspicuous absence within other population frequency databases renders these variants exceptionally intriguing for shedding light on the distinctive genetic landscape of the Spanish population. It also stresses the importance of collecting genomic data on local populations [30, 31].

When exploring CSVS at individual-level, out of the 57 distinct individual cases harboring reportable SF variants (as outlined in Table 2), 39 cases (68%) were identified as carriers of reportable variants within genes associated with the cardiovascular system. The spectrum of most prevalent cardiovascular conditions included: dilated cardiomyopathy, encompassing genes such as *BAG3*, *DES*, *FLNC*, *RBM20*, and *TTN*; arrhythmogenic right ventricular cardiomyopathy, encompassing genes *DSG2*, *PKP2* and *TMEM43*; and hypertrophic cardiomyopathy, encompassing genes *MYBPC3*, *MYH7*, and *MYL3*. This implies that more than half of individuals harboring secondary findings may potentially be susceptible to conditions associated with sudden death or heart failure, dependent on the specific implicated gene. Of the 57 documented cases, eleven instances (19%) corresponded to individuals bearing variations within genes linked to cancer-related disorders. Predominant among these was Lynch syndrome, with pathogenic variants detected in the *MSH6* and *MLH1* genes. Additional cancer-related disorders included: hereditary breast and/or ovarian cancer (*BRCA2* gene), hereditary paraganglioma– pheochromocytoma syndrome (*SDHB* and *SDHC* genes), tuberous sclerosis complex (*TSC1* and *TSC2* genes), and familial adenomatous polyposis (*APC* gene). The early identification of cancer predisposition through the analysis of secondary findings bears unequivocal advantages in the realms of disease prevention, treatment, and disease progression management.

Also, two cases (3.5%) demonstrated reportable variants within genes associated with inborn errors of metabolism phenotypes including a likely pathogenic variant in hemizygous state within the alpha-galactosidase gene (*GLA*), linked to Fabry disease and two distinct pathogenic heterozygous variants situated within the biotinidase gene (*BTD*), implicated in an autosomal recessive metabolic disorder wherein the deficiency of the enzyme results in cutaneous and neurological abnormalities.

Finally, 5 of the cases (8.8%) exhibited reportable variants tied to two diverse and distinct phenotypic categories: i) malignant hyperthermia susceptibility, with three individuals bearing diverse pathogenic variants within the ryanodine receptor 1 gene (*RYR1*) and another one showing a reportable variant within the calcium channel gene *CACNA1S* [48], and ii) hereditary hemochromatosis, an autosomal recessive disorder characterized by the pathological accumulation of excess iron within various organs, with one individual harboring homozygosity for the variant rs1800562 (p.Cys282Tyr) within the *HFE* gene. Significantly, this variant is the sole exception within the ACMG criteria for the *HFE* gene, mandating its inclusion in reporting [49].

Of notable interest among secondary findings (SFs) are also variants bearing pharmacogenetic significance [50]. However, these variants fall beyond the scope of this study because our group already conducted an exhaustive analysis of actionable pharmacogenes within the Spanish population, unveiling a staggering statistic: 98% of the Spanish population carries at least one allele associated with the potential need for therapeutic adjustments. This statistic translates to an average requirement for a therapeutic alteration across 3.31 out of the 64 implicated drugs [51].

The prevalence of SFs has been estimated in numerous global populations, showcasing a notable degree of variability, spanning from 0.5% to 8.13% (refer to Table 1). These variations stem from a multitude of factors, as elucidated earlier. Divergences can arise from disparate sequencing methodologies, sequencing depth (which can influence detection sensitivity), varying cohort sizes, ethnic diversity, genetic backgrounds, differential filtering criteria, distinct interpretations, variations in ACMG classification approaches (including InterVar, Franklin, or Varsome, as well as different versions of these repositories), and disparities in the employed clinical databases (such as ClinVar or HGMD).

The observed SF prevalence within CSVS (5%) marginally exceeds that noted in the majority of studies tabulated in Table 1. This outcome is anticipated, primarily because most of those studies are predicated upon the ACMG lists 1.0 or 2.0, encompassing 56 and 59 genes, respectively. By contrast, this study explores 78 genes designated within the ACMG v3.1 list in the CSVS repository. Remarkably, the prevalence rate found (5%) aligns closely with findings from studies based on the predecessor ACMG list (ACMG v3.0 list, encompassing 73 genes). Noteworthy examples include a Thai study (5.5%) [27] and a Chinese counterpart (5.3%) [28]. Conversely, to our knowledge, the singular published study grounded in the same ACMG list as CSVS, an examination of 863 Pakistani individuals, revealed a SF rate of 2.7% [29]. This lower prevalence is likely attributed to the smaller cohort size and divergent interpretation criteria employed in that study as compared to CSVS.

Recently, a study conducted by Codina-Solà et al., focusing on SFs within the Spanish population has been published [25]. While it parallels this study in its calculation of SFs in the Spanish population, the primary emphasis of this work lies in investigating the rate of acceptance pertaining to SFs disclosure, along with the influencing factors, within a cohort of individuals afflicted by rare genetic disorders within a hospital setting. Their investigation centered around the ACMG v2 list (encompassing only 59 genes) [11], and their cohort comprised 740 individuals, most of them exomes (736 out of 740), which hinders the detection of important non-coding SFs (e.g. splicing, etc.). The Codina-Solà study yielded pathogenic/likely pathogenic variants in only 18 genes, ultimately culminating in a SFs rate of 3.6% within their cohort, slightly lower than the SF prevalence reported in the present study (5%). This discrepancy can be attributed, at least in part, to the larger size of the CSVS cohort, the broader scope of actionable genes investigated (59 genes in ACMG v2 vs. 78 genes in this case), and the greater number of genomes included within our cohort. that yield a more uniform and comprehensive coverage and therefore a better discovery rate of variants, as previously described [29].

One of the limitations of this study comes from the short-read sequencing technologies used for the sequencing and the unavailability of parental sequences that preclude systematic phase resolution. In addition, the variant-centric character of CSVS, as well as other variant repositories makes unfeasible representing anything that is conditional to combinations with other neighbor variants, which underestimates the estimation of compound heterozygous SFs. Also, some batch effect is expected derived from the fact that, despite more recent attempts of country-wide homogenization of procedures lead by the IMPaCT project [52], sequences come from different sequencing facilities across Spain (see the original CSVS publication for details [31])

Another common limitation of SF studies, from which this study is not exempt, is the absence of copy number variants (CNVs). Given that CNVs constitute factors in hereditary diseases and can contribute as SFs, there exists the potential for an underestimation of the real SF rate. Consequently, the consideration of CNVs becomes imperative for more accurate SF estimation within the Spanish population. Notably, SPACNACS houses data pertaining to CNVs among the Spanish population [53], derived from a subset of CSVS samples. Future endeavors entail the utilization of SPACNACS to gauge the contribution of CNVs to the secondary findings rate.

## Conclusions

The presence of pathogenic or likely pathogenic variants within the Spanish Variant Server CSVS has been examined, conforming to ACMG criteria for a list of 78 actionable genes (ACMG v3.1), as part of the pursuit of secondary findings in the Spanish population. The SF rate, which stands at 5%, has been computed within CSVS and aligns with the range documented in analogous studies. The annotation for these SF variants is accessible through the CSVS website.

Therefore, population variability databases such as CSVS represent excellent platforms for the prospective exploration of SFs. Owing to its ongoing expansion through the incorporation of new individuals, the representational accuracy of Spanish variation is poised for augmentation. Furthermore, the architecture of CSVS enables periodic reevaluation of SF prevalence in tandem with the evolution of knowledge and ACMG criteria.

The outcomes of this study summarize the prevalence of pathogenic and likely pathogenic variants in the Spanish population, thereby contributing to the amplification of existing insights concerning secondary findings within this demographic. The entirety of this information can potentially empower clinicians in their endeavors to enhance the healthcare outcomes of the population.

## Data Availability

Part of the dataset analyzed during the current study are available in the European Genome Archive (EGA) repository, https://www.ebi.ac.uk/ega/datasets/EGAD00001003101.
The rest of data can be queried in the Collaborative Spanish Variant Server (CSVS) http://csvs.babelomics.org/. Aggregated data can be requested in CSVS (see downloads section)

http://csvs.babelomics.org/

https://www.ebi.ac.uk/ega/datasets/EGAD00001003101

## List of abbreviations

ACGM: American College of Medical Genetics
CES: Clinical Exome Sequencing
CNV: copy number variants
CSVS: Collaborative Spanish Variant Server
SF: Secondary finding
WES: Whole Exome Sequencing
WGS: Whole Genome Sequencing

## Ethics approval and consent to participate

Not applicable

## Availability of data and materials

Part of the dataset analyzed during the current study are available in the European Genome Archive (EGA) repository, https://www.ebi.ac.uk/ega/datasets/EGAD00001003101.

The rest of data can be queried in the Collaborative Spanish Variant Server (CSVS) http://csvs.babelomics.org/. Aggregated data can be requested in CSVS (see downloads section)

## Author Contributions

RC carried out most of the analysis and wrote the manuscript; VA, NTB, MPC helped with the analysis of the data; JPF, GB contributed to the primary analysis of the sequences; JLFR and GR developed software to query the data from the CSVS database; CL, DLL, FMO developed the applications for testing ancestry and potential kinship; BM coordinates the EnoD project and the collection of data; JD coordinated the work and wrote the final version of the manuscript. Members of the CSVS Crowdsourcing Group have contributed with human genome sequences to the CSVS.

## References

1. Lavelle TA, Feng X, Keisler M, Cohen JT, Neumann PJ, Prichard D, Schroeder BE, Salyakina D, Espinal PS, Weidner SB: Cost-effectiveness of exome and genome sequencing for children with rare and undiagnosed conditions. Genetics in Medicine 2022, 24(6):1349–1361.

2. Shen T, Pajaro-Van de Stadt SH, Yeat NC, Lin JC-H: Clinical applications of next generation sequencing in cancer: from panels, to exomes, to genomes. Frontiers in genetics 2015, 6:215.

3. Miller DT, Lee K, Abul-Husn NS, Amendola LM, Brothers K, Chung WK, Gollob MH, Gordon AS, Harrison SM, Hershberger RE: ACMG SF v3. 1 list for reporting of secondary findings in clinical exome and genome sequencing: A policy statement of the American College of Medical Genetics and Genomics (ACMG). In., vol. 24: Elsevier; 2022: 1407–1414.

4. de Wert G, Dondorp W, Clarke A, Dequeker EM, Cordier C, Deans Z, van El CG, Fellmann F, Hastings R, Hentze S: Opportunistic genomic screening. Recommendations of the European society of human genetics. European Journal of Human Genetics 2021, 29(3):365–377.

5. Green RC, Berg JS, Grody WW, Kalia SS, Korf BR, Martin CL, McGuire AL, Nussbaum RL, O’Daniel JM, Ormond KE et al: ACMG recommendations for reporting of incidental findings in clinical exome and genome sequencing. Genet Med 2013, 15(7):565–574.

6. Olfson E, Cottrell CE, Davidson NO, Gurnett CA, Heusel JW, Stitziel NO, Chen L-S, Hartz S, Nagarajan R, Saccone NL: Identification of medically actionable secondary findings in the 1000 genomes. PLOS one 2015, 10(9):e0135193.

7. Amendola LM, Dorschner MO, Robertson PD, Salama JS, Hart R, Shirts BH, Murray ML, Tokita MJ, Gallego CJ, Kim DS: Actionable exomic incidental findings in 6503 participants: challenges of variant classification. Genome research 2015, 25(3):305–315.

8. Dewey FE, Murray MF, Overton JD, Habegger L, Leader JB, Fetterolf SN, O’Dushlaine C, Van Hout CV, Staples J, Gonzaga-Jauregui C: Distribution and clinical impact of functional variants in 50,726 whole-exome sequences from the DiscovEHR study. Science 2016, 354(6319):aaf6814.

9. Jamuar SS, Kuan JL, Brett M, Tiang Z, Tan WLW, Lim JY, Liew WKM, Javed A, Liew WK, Law HY: Incidentalome from genomic sequencing: a barrier to personalized medicine? EBioMedicine 2016, 5:211–216.

10. Natarajan P, Gold NB, Bick AG, McLaughlin H, Kraft P, Rehm HL, Peloso GM, Wilson JG, Correa A, Seidman JG: Aggregate penetrance of genomic variants for actionable disorders in European and African Americans. Science translational medicine 2016, 8(364):364ra151–364ra151.

11. Kalia SS, Adelman K, Bale SJ, Chung WK, Eng C, Evans JP, Herman GE, Hufnagel SB, Klein TE, Korf BR et al: Recommendations for reporting of secondary findings in clinical exome and genome sequencing, 2016 update (ACMG SF v2. 0): a policy statement of the American College of Medical Genetics and Genomics. Genetics in medicine 2017, 19(2):249.

12. Tang CS-m, Dattani S, So M-t, Cherny SS, Tam PK, Sham PC, Garcia-Barcelo M-M: Actionable secondary findings from whole-genome sequencing of 954 East Asians. Human genetics 2018, 137:31–37.

13. Chen W, Li W, Ma Y, Zhang Y, Han B, Liu X, Zhao K, Zhang M, Mi J, Fu Y: Secondary findings in 421 whole exome-sequenced Chinese children. Human Genomics 2018, 12:1–6.

14. Haer-Wigman L, van der Schoot V, Feenstra I, Vulto-van Silfhout AT, Gilissen C, Brunner HG, Vissers LE, Yntema HG: 1 in 38 individuals at risk of a dominant medically actionable disease. European Journal of Human Genetics 2019, 27(2):325–330.

15. Yu MHC, Mak CCY, Fung JLF, Lee M, Tsang MHY, Chau JFT, Chung PH-Y, Yang W, Chan GCF, Lee SL: Actionable secondary findings in 1116 Hong Kong Chinese based on exome sequencing data. Journal of Human Genetics 2021, 66(6):637–641.

16. Van Hout CV, Tachmazidou I, Backman JD, Hoffman JD, Liu D, Pandey AK, Gonzaga-Jauregui C, Khalid S, Ye B, Banerjee N: Exome sequencing and characterization of 49,960 individuals in the UK Biobank. Nature 2020, 586(7831):749–756.

17. Kuo CW, Hwu WL, Chien YH, Hsu C, Hung MZ, Lin IL, Lai F, Lee NC: Frequency and spectrum of actionable pathogenic secondary findings in Taiwanese exomes. Molecular genetics & genomic medicine 2020, 8(10):e1455.

18. Jalkh N, Mehawej C, Chouery E: Actionable exomic secondary findings in 280 Lebanese participants. Frontiers in genetics 2020, 11:208.

19. Gordon AS, Zouk H, Venner E, Eng CM, Funke BH, Amendola LM, Carrell DS, Chisholm RL, Chung WK, Denny JC: Frequency of genomic secondary findings among 21,915 eMERGE network participants. Genetics in Medicine 2020, 22(9):1470–1477.

20. Arslan Ateş E, Türkyilmaz A, Yıldırım Ö, Alavanda C, Polat H, Demir Ş, Çebi AH, Geçkinli BB, Güney Aİ, Ata P: Secondary findings in 622 Turkish clinical exome sequencing data. Journal of Human Genetics 2021, 66(11):1113–1119.

21. Aloraini T, Alsubaie L, Alasker S, Al Muitiri A, Alswaid A, Eyiad W, Al Mutairi F, Ababneh F, Alfadhel M, Alfares A: The rate of secondary genomic findings in the Saudi population. American Journal of Medical Genetics Part A 2022, 188(1):83–88.

22. Elfatih A, Mifsud B, Syed N, Badii R, Mbarek H, Abbaszadeh F, Consortium QGPR, Estivill X, Management QGP, Ismail S: Actionable genomic variants in 6045 participants from the Qatar Genome Program. Human Mutation 2021, 42(12):1584–1601.

23. Rodríguez-Salgado LE, Silva-Aldana CT, Medina-Méndez E, Bareño-Silva J, Arcos-Burgos M, Silgado-Guzmán DF, Restrepo CM: Frequency of actionable Exomic secondary findings in 160 Colombian patients: Impact in the healthcare system. Gene 2022, 838:146699.

24. Xiao H, Zhang J-T, Dong X-R, Lu Y-L, Wu B-B, Wang H-J, Zhao Z-Y, Yang L, Zhou W-H: Secondary genomic findings in the 2020 China Neonatal Genomes Project participants. World Journal of Pediatrics 2022, 18(10):687–694.

25. Codina-Solà M, Trujillano L, Abulí A, Rovira-Moreno E, Muñoz-Cabello P, Campos B, Fernández-Álvarez P, Palau D, Carrasco E, Valenzuela I: An spanish study of secondary findings in families affected with mendelian disorders: choices, prevalence and family history. European Journal of Human Genetics 2023, 31(2):223–230.

26. Miller DT, Lee K, Chung WK, Gordon AS, Herman GE, Klein TE, Stewart DR, Amendola LM, Adelman K, Bale SJ: ACMG SF v3. 0 list for reporting of secondary findings in clinical exome and genome sequencing: a policy statement of the American College of Medical Genetics and Genomics (ACMG). Genetics in medicine 2021, 23(8):1381–1390.

27. Chetruengchai W, Shotelersuk V: Actionable secondary findings in the 73 ACMG-recommended genes in 1559 Thai exomes. Journal of human genetics 2022, 67(3):137–142.

28. Huang Y, Liu B, Shi J, Zhao S, Xu K, Sun L, Chen N, Tian W, Zhang J, Wu N: Landscape of Secondary Findings in Chinese Population: A Practice of ACMG SF v3. 0 List. Journal of Personalized Medicine 2022, 12(9):1503.

29. Skrahin A, Cheema HA, Hussain M, Rana NN, Rehman KU, Kumar R, Oprea G, Ameziane N, Rolfs A, Skrahina V: Secondary findings in a large Pakistani cohort tested with whole genome sequencing. Life Science Alliance 2023, 6(3).

30. Dopazo J, Amadoz A, Bleda M, Garcia-Alonso L, Alemán A, García-García F, Rodriguez JA, Daub JT, Muntané G, Rueda A et al: 267 Spanish Exomes Reveal Population-Specific Differences in Disease-Related Genetic Variation. Molecular Biology and Evolution 2016, 33(5):1205–1218.

31. Peña-Chilet M, Roldán G, Perez-Florido J, Ortuño FM, Carmona R, Aquino V, Lopez-Lopez D, Loucera C, Fernandez-Rueda JL, Gallego A et al: CSVS, a crowdsourcing database of the Spanish population genetic variability. Nucleic Acids Research 2020, 49(D1):D1130–D1137.

32. Mathieson I, McVean G: Differential confounding of rare and common variants in spatially structured populations. Nat Genet 2012, 44(3):243–246.

33. The Medical Genome Project [http://www.medicalgenomeproject.com/]

34. Gui H, Schriemer D, Cheng WW, Chauhan RK, Antiňolo G, Berrios C, Bleda M, Brooks AS, Brouwer RW, Burns AJ: Whole exome sequencing coupled with unbiased functional analysis reveals new Hirschsprung disease genes. Genome biology 2017, 18(1):1–13.

35. Gallego-Martinez A, Lopez-Escamez JA: Genetic architecture of Meniere’s disease. Hearing research 2020, 397:107872.

36. Torrent-Vernetta A, Gaboli M, Castillo-Corullón S, Mondéjar-López P, Sanz Santiago V, Costa-Colomer J, Osona B, Torres-Borrego J, de la Serna-Blázquez O, Bellón Alonso S et al: Incidence and Prevalence of Children’s Diffuse Lung Disease in Spain. Archivos de bronconeumologia 2022, 58(1):22–29.

37. Germline pipeline [https://www.clinbioinfosspa.es/content/germline-pipeline]

38. Chen S, Francioli LC, Goodrich JK, Collins RL, Kanai M, Wang Q, Alföldi J, Watts NA, Vittal C, Gauthier LD: A genomic mutational constraint map using variation in 76,156 human genomes. Nature 2024, 625(7993):92-100.

39. Landrum MJ, Lee JM, Benson M, Brown GR, Chao C, Chitipiralla S, Gu B, Hart J, Hoffman D, Jang W: ClinVar: improving access to variant interpretations and supporting evidence. Nucleic acids research 2018, 46(D1):D1062–D1067.

40. Li Q, Wang K: InterVar: clinical interpretation of genetic variants by the 2015 ACMG-AMP guidelines. The American Journal of Human Genetics 2017, 100(2):267–280.

41. Feder J, Gnirke A, Thomas W, Tsuchihashi Z, Ruddy D, Basava A, Dormishian F, Domingo Jr R, Ellis M, Fullan A: A novel MHC class I–like gene is mutated in patients with hereditary haemochromatosis. Nature genetics 1996, 13(4):399–408.

42. Gallego CJ, Burt A, Sundaresan AS, Ye Z, Shaw C, Crosslin DR, Crane PK, Fullerton SM, Hansen K, Carrell D: Penetrance of Hemochromatosis in HFE Genotypes Resulting in p. Cys282Tyr and p.[Cys282Tyr];[His63Asp] in the eMERGE Network. The American Journal of Human Genetics 2015, 97(4):512–520.

43. Al-Tassan N, Chmiel NH, Maynard J, Fleming N, Livingston AL, Williams GT, Hodges AK, Davies DR, David SS, Sampson JR: Inherited variants of MYH associated with somatic G: C→ T: A mutations in colorectal tumors. Nature genetics 2002, 30(2):227–232.

44. Bull PC, Thomas GR, Rommens JM, Forbes JR, Cox DW: The Wilson disease gene is a putative copper transporting P–type ATPase similar to the Menkes gene. Nature genetics 1993, 5(4):327–337.

45. Herman DS, Lam L, Taylor MR, Wang L, Teekakirikul P, Christodoulou D, Conner L, DePalma SR, McDonough B, Sparks E: Truncations of titin causing dilated cardiomyopathy. New England Journal of Medicine 2012, 366(7):619–628.

46. Rampazzo A, Nava A, Malacrida S, Beffagna G, Bauce B, Rossi V, Zimbello R, Simionati B, Basso C, Thiene G: Mutation in human desmoplakin domain binding to plakoglobin causes a dominant form of arrhythmogenic right ventricular cardiomyopathy. The American Journal of human genetics 2002, 71(5):1200–1206.

47. Priori SG, Napolitano C, Tiso N, Memmi M, Vignati G, Bloise R, Sorrentino V, Danieli GA: Mutations in the cardiac ryanodine receptor gene (hRyR2) underlie catecholaminergic polymorphic ventricular tachycardia. Circulation 2001, 103(2):196–200.

48. Monnier N, Procaccio V, Stieglitz P, Lunardi J: Malignant-hyperthermia susceptibility is associated with a mutation of the a1-subunit of the human dihydropyridine-sensitive L-type voltage-dependent calcium-channel receptor in skeletal muscle. The American Journal of Human Genetics 1997, 60(6):1316–1325.

49. Miller DT, Lee K, Gordon AS, Amendola LM, Adelman K, Bale SJ, Chung WK, Gollob MH, Harrison SM, Herman GE: Recommendations for reporting of secondary findings in clinical exome and genome sequencing, 2021 update: a policy statement of the American College of Medical Genetics and Genomics (ACMG). Genetics in Medicine 2021, 23(8):1391–1398.

50. Relling M, Klein T: CPIC: clinical pharmacogenetics implementation consortium of the pharmacogenomics research network. Clinical Pharmacology & Therapeutics 2011, 89(3):464–467.

51. Nunez-Torres R, Pita G, Peña-Chilet M, López-López D, Zamora J, Roldán G, Herráez B, Álvarez N, Alonso MR, Dopazo J et al: A Comprehensive Analysis of 21 Actionable Pharmacogenes in the Spanish Population: From Genetic Characterisation to Clinical Impact. Pharmaceutics 2023, 15(4).

52. Infraestructura de apoyo al diagnóstico con tecnologías genómicas de alta complejidad [https://genomica-impact.es/]

53. López-López D, Roldán G, Fernández-Rueda JL, Bostelmann G, Carmona R, Aquino V, Perez-Florido J, Ortuño F, Pita G, Núñez-Torres R: A crowdsourcing database for the copy-number variation of the Spanish population. Human Genomics 2023, 17(1):1–12.

