## Supplementary Table 1 for "Unveiling the Landscape of Reportable Genetic Secondary Findings in the Spanish Population: A Comprehensive Analysis Using the Collaborative Spanish Variant Server Database"

**Supplementary Table 1. Pathogenic or likely pathogenic variants according with ClinVar and/or ACMG detected in 78 actionable genes (ACMG v3.1) in the analysis of individual and aggregate samples of CSVS database**. The column labels are:

General Phenotype category, which corresponds to General categories of diseases defined by ACMG for reporting secondary findings [9]. Phenotype, are General phenotype associated with the gene; Gene, designates the gene in which the variant is located; Inheritance, informs on the inheritance mode for the disease (AD, autosomal dominant; AR, autosomal recessive; SD, semidominant; XL, X-linked); Variant(Grch37), is the variant in format “chromosome : position : reference allele : alternative allele”, according to genome reference Grch37; dbSNP ID is the variant identifier in dbSNP database (rs); Genotype counts, represent genotype counts for: 0/0: homozygous reference, 0/1: heterozygous, 1/1: homozygous alternative and ./.: missing; Allele Frequency in CSVS lists allele frequencies in the CSVS population: refFreq: allele frequency for reference (0), altFreq: allele frequency for alternative (1), MAF: minor allele frequency, i.e. the lowest value between refFreq and altFreq; Carriers AllSamplesCSVS, contains the number of individuals carrying the variant taking into account both aggregate and individual samples; Carriers SingleSamplesCSVS contains the number of individuals carrying the variant considering only individual samples; Consequence Type describes the worst consequence type found among all transcripts, calculated according the Ensembl variation [71];. Frequency in 1000 Genomes Project: Alternate Allele Frequency in 1000 genomes project database (phase 3), 1kG_phase3_EUR: Frequency recorded in 1000G for the variant selected for the European population, 1kG_phase3_ALL: Frequency recorded in 1000G for the variant selected for the entire population; Prevalence in AllSamplesCSVS (%) is the percentage of carriers of the variant; GERP contains GERP score estimates the level of conservation of positions. Positive scores represent a substitution deficit and this indicate that a site may be under evolutionary constraint. Negative scores indicate that a site is probably evolving neutrally. Some author suggests that scores >=2 indicate evolutionary constraint and >=3 indicate purifying selection; SIFT contains SIFT score predicts whether an amino acid substitution affects protein function. SIFT value less than 0.05 represents a 'deleterious' prediction. SIFT value greater than or equal to 0.05 represents a 'tolerated' prediction; Polyphen contains Polyphen score predicts the possible impact of an aminoacidic substitution on the structure and function of a protein. Polyphen scores can be benign (<0.446), possibly damaging (0.446-0.908) or probably damaging (>0.908); CADD contains the CADD tool scores indicating the deleteriousness of SNVs and indels, where higher values indicate more likely to have deleterious effects. CADD scaled score is provided.; ClinVarID is the variation ID for the variant in the clinical variation database ClinVar [42]; ClinVarInterpretation is the clinical interpretation for the variant in the clinical variation database ClinVar [42]; ClinicalSignificance (Clinvar/ACMG) is the clinical significance of the variant according to ClinVar database if applicable or ACMG classification (P=pathogenic, LP=likely pathogenic). Those variants with conflicting interpretations of pathogenicity in ClinVar with at least one interpretation as P or LP were classified according to ACMG guidelines using InterVar [44] and only those labeled as pathogenic or likely pathogenic were retained; Reportable: describes if the variant reportable according to ACMG criteria (Yes or No) [9] and the criterion for the decision: a) For genes associated with autosomal dominant phenotypes, all pathogenic or likely pathogenic variants should be reported, b) For genes associated with autosomal recessive phenotypes, one likely pathogenic and/or pathogenic variant in homozygous or (*) two different likely pathogenic and/or pathogenic variants in the gene (compound heterozygous) are needed to be reportable, c) In the case of X-linked phenotypes that are apparently hemizygous, heterozygous or homozygous, any pathogenic or likely pathogenic variant should be reported, d) For the gene HFE (hereditary hemochromatosis, with autosomal recessive inheritance), ACMG recommends reporting only homozygotes for the rs1800562 (p.Cys282Tyr) variant.

| General Phenotype category | Phenotype | Gene | Inheritance | Variant(Grch37) | dbSNP ID | 0/0 | Genotype counts 0/1 | 1/1 | ./. | Allele Frequency in CSVS refFreq | altFreq | MAF | Carriers AllSamplesCSVS | Carriers SingleSamplesCSVS | Prevalence in AllSamplesCSVS (%) | Consequence Type | Frequency in 1000 Genomes Project 1kG_phase3_EUR | 1kG_phase3_ALL | GERP | SIFT | Polyphen | CADD | ClinVarID | ClinVarInterpretation | ClinicalSignificance (Clinvar/ACMG) | Reportable |
|---|---|---|---|---|---|---|---|---|---|---|---|---|---|---|---|---|---|---|---|---|---|---|---|---|---|---|
| Genes related to cardiovascular phenotypes | Catecholaminergic polymorphic ventricular tachycardia | CASQ2 | AR | 1:116269614:T:A | rs1342655330 | 2019 | 1 | 0 | 4 | 1.0 | 0.0 | 0.0 | 1 | 0 | 0.050 | stop_gained | - | - | 4.46999979019165 | - | - | 38.0 | - | - | P | NO, b) |
| Genes related to cancer phenotypes | Hereditary paraganglioma–pheochromocytoma syndrome | SDHC | AD | 1:161284196:A:G | rs755235380 | 2009 | 1 | 0 | 0 | 1.0 | 0.0 | 0.0 | 1 | 1 | 0.050 | start_lost | - | - | 5.230000019073486 | 0.01 | 0.596 | 23.700000762939453 | 407060 | Pathogenic | P | YES, a) |
| Genes related to cancer phenotypes | Hereditary paraganglioma–pheochromocytoma syndrome | SDHB | AD | 1:17349184:CT:- | rs762812025 | 2021 | 1 | 0 | 0 | 1.0 | 0.0 | 0.0 | 1 | 0 | 0.049 | frameshift_variant | - | - | 4.380000114440918 | - | - | - | 438428 | Pathogenic | P | YES, a) |
| Genes related to cancer phenotypes | Hereditary paraganglioma–pheochromocytoma syndrome | SDHB | AD | 1:17354244:C:T | rs528442805 | 2021 | 1 | 0 | 0 | 1.0 | 0.0 | 0.0 | 1 | 1 | 0.049 | synonymous_variant | 0.0009940357 | 0.004672897 | 5.380000114440918 | - | - | 19.25 | 438418 | Likely pathogenic | LP | YES, a) |
| Genes related to miscellaneous phenotypes | Malignant hyperthermia | CACNA1S | AD | 1:201010662:G:A | rs550371466 | 2095 | 1 | 0 | 0 | 1.0 | 0.0 | 0.0 | 1 | 1 | 0.048 | stop_gained | - | - | 3.5999999046325684 | - | - | 38.0 | 402469 | Conflicting interpretations of p | P | YES, a) |
| Genes related to miscellaneous phenotypes | Malignant hyperthermia | CACNA1S | AD | 1:201021747:-:TTTG | - | 2095 | 1 | 0 | 0 | 1.0 | 0.0 | 0.0 | 1 | 0 | 0.048 | frameshift_variant | - | - | 4.679999828338623 | - | - | - | - | - | LP | YES, a) |
| Genes related to miscellaneous phenotypes | Malignant hyperthermia | CACNA1S | AD | 1:201021751:T:- | - | 2095 | 1 | 0 | 0 | 1.0 | 0.0 | 0.0 | 1 | 0 | 0.048 | frameshift_variant | - | - | 4.679999828338623 | - | - | - | - | - | LP | YES, a) |
| Genes related to miscellaneous phenotypes | Malignant hyperthermia | CACNA1S | AD | 1:201021754:A:- | - | 2095 | 1 | 0 | 0 | 1.0 | 0.0 | 0.0 | 1 | 0 | 0.048 | frameshift_variant | - | - | 4.679999828338623 | - | - | - | - | - | LP | YES, a) |
| Genes related to miscellaneous phenotypes | Malignant hyperthermia | CACNA1S | AD | 1:201021757:-:TT | - | 2095 | 1 | 0 | 0 | 1.0 | 0.0 | 0.0 | 1 | 0 | 0.048 | frameshift_variant | - | - | 3.75 | - | - | - | - | - | LP | YES, a) |
| Genes related to miscellaneous phenotypes | Malignant hyperthermia | CACNA1S | AD | 1:201021758:AGGC:- | - | 2095 | 1 | 0 | 0 | 1.0 | 0.0 | 0.0 | 1 | 0 | 0.048 | frameshift_variant | - | - | 4.550000190734863 | - | - | - | - | - | LP | YES, a) |
| Genes related to miscellaneous phenotypes | Malignant hyperthermia | CACNA1S | AD | 1:201029886:TTTTT:- | - | 2095 | 1 | 0 | 0 | 1.0 | 0.0 | 0.0 | 1 | 0 | 0.048 | frameshift_variant | - | - | 4.949999809265137 | - | - | - | - | - | LP | YES, a) |
| Genes related to miscellaneous phenotypes | Malignant hyperthermia | CACNA1S | AD | 1:201029891:-:GGGGG | - | 2095 | 1 | 0 | 0 | 1.0 | 0.0 | 0.0 | 1 | 0 | 0.048 | frameshift_variant | - | - | 4.949999809265137 | - | - | - | - | - | LP | YES, a) |
| Genes related to cardiovascular phenotypes | Dilated cardiomyopathy | TNNT2 | AD | 1:201335991:TTGGG:- | - | 2080 | 1 | 0 | 0 | 1.0 | 0.0 | 0.0 | 1 | 0 | 0.048 | frameshift_variant | - | - | 0.7630000114440918 | - | - | - | - | - | LP | YES, a) |
| Genes related to cardiovascular phenotypes | Dilated cardiomyopathy | TNNT2 | AD | 1:201335996:-:AACCC | - | 2080 | 1 | 0 | 0 | 1.0 | 0.0 | 0.0 | 1 | 0 | 0.048 | frameshift_variant | - | - | 0.7630000114440918 | - | - | - | - | - | LP | YES, a) |
| Genes related to cardiovascular phenotypes | Catecholaminergic polymorphic ventricular tachycardia | RYR2 | AD | 1:237711731:TT:- | - | 2080 | 1 | 0 | 0 | 1.0 | 0.0 | 0.0 | 1 | 0 | 0.048 | frameshift_variant | - | - | 5.809999942779541 | - | - | - | - | - | LP | YES, a) |
| Genes related to cardiovascular phenotypes | Catecholaminergic polymorphic ventricular tachycardia | RYR2 | AD | 1:237796883:CACC:- | - | 2080 | 1 | 0 | 0 | 1.0 | 0.0 | 0.0 | 1 | 0 | 0.048 | frameshift_variant | - | - | 0.7039999961853027 | - | - | - | - | - | LP | YES, a) |
| Genes related to cardiovascular phenotypes | Catecholaminergic polymorphic ventricular tachycardia | RYR2 | AD | 1:237862276:-:AACG | - | 2080 | 1 | 0 | 0 | 1.0 | 0.0 | 0.0 | 1 | 0 | 0.048 | frameshift_variant | - | - | -10.300000190734863 | - | - | - | - | - | LP | YES, a) |
| Genes related to cardiovascular phenotypes | Catecholaminergic polymorphic ventricular tachycardia | RYR2 | AD | 1:237862284:-:G | - | 2080 | 1 | 0 | 0 | 1.0 | 0.0 | 0.0 | 1 | 0 | 0.048 | frameshift_variant | - | - | 5.46999979019165 | - | - | - | - | - | LP | YES, a) |
| Genes related to cardiovascular phenotypes | Catecholaminergic polymorphic ventricular tachycardia | RYR2 | AD | 1:237862287:CAAC:- | - | 2080 | 1 | 0 | 0 | 1.0 | 0.0 | 0.0 | 1 | 0 | 0.048 | frameshift_variant | - | - | 5.46999979019165 | - | - | - | - | - | LP | YES, a) |
| Genes related to cardiovascular phenotypes | Catecholaminergic polymorphic ventricular tachycardia | RYR2 | AD | 1:237870339:-:CC | - | 2080 | 1 | 0 | 0 | 1.0 | 0.0 | 0.0 | 1 | 0 | 0.048 | frameshift_variant | - | - | 5.71999979019165 | - | - | - | - | - | LP | YES, a) |
| Genes related to cancer phenotypes | MUTYH-associated polyposis | MUTYH | AR | 1:45796205:G:A | rs932830392 | 2021 | 1 | 0 | 0 | 1.0 | 0.0 | 0.0 | 1 | 0 | 0.049 | stop_gained | - | - | 4.349999904632568 | - | - | 39.0 | 620207 | Conflicting interpretations of p | P | NO, b) |
| Genes related to cancer phenotypes | MUTYH-associated polyposis | MUTYH | AR | 1:45796229:C:A | rs587782228 | 2021 | 1 | 0 | 0 | 1.0 | 0.0 | 0.0 | 1 | 0 | 0.049 | missense_variant | - | - | 5.5 | 0.0 | 0.993 | 31.0 | 219953 | Pathogenic/Likely pathogenic | P/LP | NO, b) |
| Genes related to cancer phenotypes | MUTYH-associated polyposis | MUTYH | AR | 1:45797372:G:- | rs587778536 | 2021 | 1 | 0 | 0 | 1.0 | 0.0 | 0.0 | 1 | 1 | 0.049 | frameshift_variant | - | - | 2.430000066757202 | - | - | - | 134860 | Conflicting interpretations of p | LP | NO, b) |
| Genes related to cardiovascular phenotypes | Familial hypercholesterolemia | PCSK9 | AD | 1:55525158:G:A | - | 2078 | 1 | 0 | 0 | 1.0 | 0.0 | 0.0 | 1 | 0 | 0.048 | splice_acceptor_variant | - | - | 3.630000114440918 | - | - | 22.399999618530273 | - | - | P | YES, a) |
| Genes related to miscellaneous phenotypes | RPE65-related retinopathy | RPE65 | AR | 1:68904671:A:T | rs61752905 | 1991 | 1 | 0 | 0 | 1.0 | 0.0 | 0.0 | 1 | 1 | 0.050 | missense_variant | 0.0009940357 | 0.004672897 | 4.869999885559082 | 0.0 | 0.998 | 25.700000762939453 | 98900 | Likely pathogenic | LP | NO, b) |
| Genes related to miscellaneous phenotypes | RPE65-related retinopathy | RPE65 | AR | 1:68904938:-:AG | - | 1991 | 1 | 0 | 0 | 1.0 | 0.0 | 0.0 | 1 | 0 | 0.050 | frameshift_variant | - | - | -1.590000033378601 | - | - | - | - | - | LP | NO, b) |
| Genes related to miscellaneous phenotypes | RPE65-related retinopathy | RPE65 | AR | 1:68904945:-:C | - | 1991 | 1 | 0 | 0 | 1.0 | 0.0 | 0.0 | 1 | 0 | 0.050 | frameshift_variant | - | - | 5.610000133514404 | - | - | - | - | - | LP | NO, b) |
| Genes related to miscellaneous phenotypes | RPE65-related retinopathy | RPE65 | AR | 1:68905251:C:A | rs192907397 | 1991 | 1 | 0 | 0 | 1.0 | 0.0 | 0.0 | 1 | 1 | 0.050 | missense_variant | - | - | 5.829999923706055 | 0.12 | 0.973 | 26.899999618530273 | 559520 | Likely pathogenic | LP | NO, b) |
| Genes related to miscellaneous phenotypes | RPE65-related retinopathy | RPE65 | AR | 1:68915577:C:G | - | 1991 | 1 | 0 | 0 | 1.0 | 0.0 | 0.0 | 1 | 0 | 0.050 | splice_donor_variant | - | - | 5.690000057220459 | - | - | 24.700000762939453 | - | - | P | NO, b) |
| Genes related to cardiovascular phenotypes | Dilated cardiomyopathy | RBM20 | AD | 10:112404358:AGCCA:- | - | 2053 | 1 | 0 | 0 | 1.0 | 0.0 | 0.0 | 1 | 1 | 0.049 | frameshift_variant | - | - | -3.740000009536743 | - | - | - | - | - | LP | YES, a) |
| Genes related to cardiovascular phenotypes | Dilated cardiomyopathy | RBM20 | AD | 10:112541459:-:CC | - | 2080 | 1 | 0 | 0 | 1.0 | 0.0 | 0.0 | 1 | 0 | 0.048 | frameshift_variant | - | - | 4.800000190734863 | - | - | - | - | - | LP | YES, a) |
| Genes related to cardiovascular phenotypes | Dilated cardiomyopathy | RBM20 | AD | 10:112541506:-:GGAG | - | 2080 | 1 | 0 | 0 | 1.0 | 0.0 | 0.0 | 1 | 1 | 0.048 | frameshift_variant | - | - | 1.9299999475479126 | - | - | - | - | - | LP | YES, a) |
| Genes related to cardiovascular phenotypes | Dilated cardiomyopathy | RBM20 | AD | 10:112557333:-:C | - | 2080 | 1 | 0 | 0 | 1.0 | 0.0 | 0.0 | 1 | 1 | 0.048 | frameshift_variant | - | - | 5.659999847412109 | - | - | - | - | - | LP | YES, a) |
| Genes related to cardiovascular phenotypes | Dilated cardiomyopathy | RBM20 | AD | 10:112572050:G:- | - | 2067 | 1 | 0 | 1 | 1.0 | 0.0 | 0.0 | 1 | 0 | 0.048 | frameshift_variant | - | - | 5.690000057220459 | - | - | - | - | - | LP | YES, a) |
| Genes related to cardiovascular phenotypes | Dilated cardiomyopathy | RBM20 | AD | 10:112572056:-:G | - | 2067 | 1 | 0 | 1 | 1.0 | 0.0 | 0.0 | 1 | 0 | 0.048 | frameshift_variant | - | - | 2.4600000381469727 | - | - | - | - | - | LP | YES, a) |
| Genes related to cardiovascular phenotypes | Dilated cardiomyopathy | BAG3 | AD | 10:121429482:TG:- | - | 2080 | 1 | 0 | 0 | 1.0 | 0.0 | 0.0 | 1 | 0 | 0.048 | frameshift_variant | - | - | 5.400000095367432 | - | - | - | - | - | LP | YES, a) |
| Genes related to cardiovascular phenotypes | Dilated cardiomyopathy | BAG3 | AD | 10:121429487:-:GA | - | 2080 | 1 | 0 | 0 | 1.0 | 0.0 | 0.0 | 1 | 0 | 0.048 | frameshift_variant | - | - | 5.400000095367432 | - | - | - | - | - | LP | YES, a) |
| Genes related to cardiovascular phenotypes | Dilated cardiomyopathy | BAG3 | AD | 10:121429549:C:T | rs387906875 | 2080 | 1 | 0 | 0 | 1.0 | 0.0 | 0.0 | 1 | 1 | 0.048 | stop_gained | 0.0009940357 | 0.004672897 | 3.4600000381469727 | - | - | 37.0 | 30397 | Pathogenic/Likely pathogenic | P/LP | YES, a) |
| Genes related to cancer phenotypes | Familial medullary thyroid cancer | RET | AD | 10:43596096:TC:- | - | 2021 | 1 | 0 | 0 | 1.0 | 0.0 | 0.0 | 1 | 0 | 0.049 | frameshift_variant | - | - | 2.3299999237060547 | - | - | - | - | - | LP | YES, a) |
| Genes related to cancer phenotypes | Familial medullary thyroid cancer | RET | AD | 10:43609097:G:T | rs79781594 | 2021 | 1 | 0 | 0 | 1.0 | 0.0 | 0.0 | 1 | 0 | 0.049 | missense_variant | - | - | 4.920000076293945 | 0.0 | 0.315 | 24.0 | 24902 | Pathogenic | P | YES, a) |
| Genes related to cancer phenotypes | Familial medullary thyroid cancer | RET | AD | 10:43613879:GGTC:- | - | 2021 | 1 | 0 | 0 | 1.0 | 0.0 | 0.0 | 1 | 0 | 0.049 | frameshift_variant | - | - | 4.670000076293945 | - | - | - | - | - | LP | YES, a) |
| Genes related to cancer phenotypes | Familial medullary thyroid cancer | RET | AD | 10:43613884:-:ACAC | - | 2021 | 1 | 0 | 0 | 1.0 | 0.0 | 0.0 | 1 | 0 | 0.049 | frameshift_variant | - | - | 5.579999923706055 | - | - | - | - | - | LP | YES, a) |
| Genes related to cancer phenotypes | PTEN hamartoma tumor syndrome | PTEN | AD | 10:89653808:G:C | rs786204854 | 2021 | 1 | 0 | 0 | 1.0 | 0.0 | 0.0 | 1 | 0 | 0.049 | missense_variant | - | - | 5.190000057220459 | 0.05 | 0.813 | 25.299999237060547 | 189400 | Conflicting interpretations of p | LP | YES, a) |
| Genes related to cancer phenotypes | PTEN hamartoma tumor syndrome | PTEN | AD | 10:89711897:GGCGC:- | - | 1961 | 1 | 0 | 2 | 1.0 | 0.0 | 0.0 | 1 | 0 | 0.051 | frameshift_variant | - | - | 4.840000152587891 | - | - | - | - | - | LP | YES, a) |
| Genes related to cancer phenotypes | PTEN hamartoma tumor syndrome | PTEN | AD | 10:89711902:-:TTATA | - | 1961 | 1 | 0 | 2 | 1.0 | 0.0 | 0.0 | 1 | 0 | 0.051 | frameshift_variant | - | - | 4.840000152587891 | - | - | - | - | - | LP | YES, a) |
| Genes related to cardiovascular phenotypes | Long QT syndrome types 1 and 2 | KCNQ1 | AD | 11:2591930:T:C | rs199473661 | 2080 | 1 | 0 | 0 | 1.0 | 0.0 | 0.0 | 1 | 1 | 0.048 | missense_variant | - | - | 4.400000095367432 | 0.0 | 0.995 | 27.0 | 67081 | Conflicting interpretations of p | LP | YES, a) |
| Genes related to cardiovascular phenotypes | Long QT syndrome types 1 and 2 | KCNQ1 | AD | 11:2610011:C:- | - | 2080 | 1 | 0 | 0 | 1.0 | 0.0 | 0.0 | 1 | 0 | 0.048 | frameshift_variant | - | - | 1.8700000047683716 | - | - | - | - | - | LP | YES, a) |
| Genes related to cardiovascular phenotypes | Long QT syndrome types 1 and 2 | KCNQ1 | AD | 11:2610020:-:A | - | 2080 | 1 | 0 | 0 | 1.0 | 0.0 | 0.0 | 1 | 0 | 0.048 | frameshift_variant | - | - | 5.019999980926514 | - | - | - | - | - | LP | YES, a) |
| Genes related to cardiovascular phenotypes | Hypertrophic cardiomyopathy | MYBPC3 | AD | 11:47357513:G:- | - | 2080 | 1 | 0 | 0 | 1.0 | 0.0 | 0.0 | 1 | 1 | 0.048 | frameshift_variant | - | - | 1.090000033378601 | - | - | - | - | - | LP | YES, a) |
| Genes related to cardiovascular phenotypes | Hypertrophic cardiomyopathy | MYBPC3 | AD | 11:47372138:-:GGCAG | rs2095899755 | 2080 | 1 | 0 | 0 | 1.0 | 0.0 | 0.0 | 1 | 1 | 0.048 | frameshift_variant | - | - | 2.440000057220459 | - | - | - | 940607 | Pathogenic | P | YES, a) |
| Genes related to cancer phenotypes | Hereditary paraganglioma–pheochromocytoma syndrome | SDHAF2 | AD | 11:61205292:G:A | rs113560320 | 1975 | 1 | 0 | 0 | 1.0 | 0.0 | 0.0 | 1 | 0 | 0.051 | missense_variant | - | - | 5.260000228881836 | 0.0 | 1.0 | 31.0 | 401 | Pathogenic | P | YES, a) |
| Genes related to cardiovascular phenotypes | Hypertrophic cardiomyopathy | MYL2 | AD | 12:111348894:T:C | rs397516407 | 2080 | 1 | 0 | 0 | 1.0 | 0.0 | 0.0 | 1 | 0 | 0.048 | missense_variant | - | - | 4.949999809265137 | 0.03 | 0.952 | 28.700000762939453 | 181421 | Likely pathogenic | LP | YES, a) |
| Genes related to cardiovascular phenotypes | Arrhythmogenic right ventricular cardiomyopathy | PKP2 | AD | 12:32974422:G:- | rs764817683 | 2068 | 1 | 0 | 0 | 1.0 | 0.0 | 0.0 | 1 | 1 | 0.048 | frameshift_variant | - | - | 0.15700000524520874 | - | - | - | 202022 | Pathogenic | P | YES, a) |
| Genes related to cardiovascular phenotypes | Arrhythmogenic right ventricular cardiomyopathy | PKP2 | AD | 12:33003723:A:T | - | 2068 | 1 | 0 | 0 | 1.0 | 0.0 | 0.0 | 1 | 1 | 0.048 | stop_gained | - | - | 4.699999809265137 | - | - | 41.0 | - | - | P | YES, a) |
| Genes related to miscellaneous phenotypes | Hereditary hemorrhagic telangiectasia | ACVRL1 | AD | 12:52306935:C:- | - | 2080 | 1 | 0 | 0 | 1.0 | 0.0 | 0.0 | 1 | 0 | 0.048 | frameshift_variant | - | - | -0.7160000205039978 | - | - | - | - | - | LP | YES, a) |
| Genes related to miscellaneous phenotypes | Hereditary hemorrhagic telangiectasia | ACVRL1 | AD | 12:52306941:-:C | - | 2078 | 1 | 0 | 2 | 1.0 | 0.0 | 0.0 | 1 | 0 | 0.048 | frameshift_variant | - | - | 1.9700000286102295 | - | - | - | - | - | LP | YES, a) |
| Genes related to cancer phenotypes | Hereditary breast and/or ovarian cancer | BRCA2 | AD | 13:32910616:-:A | - | 2021 | 1 | 0 | 0 | 1.0 | 0.0 | 0.0 | 1 | 0 | 0.049 | frameshift_variant | - | - | -3.1600000858306885 | - | - | - | - | - | LP | YES, a) |
| Genes related to cancer phenotypes | Hereditary breast and/or ovarian cancer | BRCA2 | AD | 13:32910619:G:- | - | 2021 | 1 | 0 | 0 | 1.0 | 0.0 | 0.0 | 1 | 0 | 0.049 | frameshift_variant | - | - | 1.7999999523162842 | - | - | - | - | - | LP | YES, a) |
| Genes related to cancer phenotypes | Hereditary breast and/or ovarian cancer | BRCA2 | AD | 13:32910868:C:A | rs80358503 | 2021 | 1 | 0 | 0 | 1.0 | 0.0 | 0.0 | 1 | 0 | 0.049 | stop_gained | - | - | -1.159999966621399 | - | - | 28.299999237060547 | 51274 | Pathogenic | P | YES, a) |
| Genes related to cancer phenotypes | Hereditary breast and/or ovarian cancer | BRCA2 | AD | 13:32910932:CCCA:- | - | 2021 | 1 | 0 | 0 | 1.0 | 0.0 | 0.0 | 1 | 0 | 0.049 | frameshift_variant | - | - | -10.100000381469727 | - | - | - | - | - | LP | YES, a) |
| Genes related to cancer phenotypes | Hereditary breast and/or ovarian cancer | BRCA2 | AD | 13:32910937:-:TTGC | - | 2021 | 1 | 0 | 0 | 1.0 | 0.0 | 0.0 | 1 | 0 | 0.049 | frameshift_variant | - | - | -0.628000020980835 | - | - | - | - | - | LP | YES, a) |
| Genes related to cancer phenotypes | Hereditary breast and/or ovarian cancer | BRCA2 | AD | 13:32910947:CAAG:- | - | 2021 | 1 | 0 | 0 | 1.0 | 0.0 | 0.0 | 1 | 0 | 0.049 | frameshift_variant | - | - | 2.8299999237060547 | - | - | - | - | - | LP | YES, a) |
| Genes related to cancer phenotypes | Hereditary breast and/or ovarian cancer | BRCA2 | AD | 13:32910952:-:CCTC | - | 2021 | 1 | 0 | 0 | 1.0 | 0.0 | 0.0 | 1 | 0 | 0.049 | frameshift_variant | - | - | 4.400000095367432 | - | - | - | - | - | LP | YES, a) |
| Genes related to cancer phenotypes | Hereditary breast and/or ovarian cancer | BRCA2 | AD | 13:32910962:-:C | - | 2021 | 1 | 0 | 0 | 1.0 | 0.0 | 0.0 | 1 | 0 | 0.049 | frameshift_variant | - | - | 1.5099999904632568 | - | - | - | - | - | LP | YES, a) |
| Genes related to cancer phenotypes | Hereditary breast and/or ovarian cancer | BRCA2 | AD | 13:32911025:CCTT:- | - | 2021 | 1 | 0 | 0 | 1.0 | 0.0 | 0.0 | 1 | 0 | 0.049 | frameshift_variant | - | - | 3.0299999713897705 | - | - | - | - | - | LP | YES, a) |
| Genes related to cancer phenotypes | Hereditary breast and/or ovarian cancer | BRCA2 | AD | 13:32911031:-:GGAC | - | 2021 | 1 | 0 | 0 | 1.0 | 0.0 | 0.0 | 1 | 0 | 0.049 | frameshift_variant | - | - | -1.9500000476837158 | - | - | - | - | - | LP | YES, a) |
| Genes related to cancer phenotypes | Hereditary breast and/or ovarian cancer | BRCA2 | AD | 13:32911473:CAGGA:- | - | 2021 | 1 | 0 | 0 | 1.0 | 0.0 | 0.0 | 1 | 0 | 0.049 | frameshift_variant | - | - | -1.4900000095367432 | - | - | - | - | - | LP | YES, a) |
| Genes related to cancer phenotypes | Hereditary breast and/or ovarian cancer | BRCA2 | AD | 13:32911483:AG:- | - | 2021 | 1 | 0 | 0 | 1.0 | 0.0 | 0.0 | 1 | 0 | 0.049 | frameshift_variant | - | - | -3.069999933242798 | - | - | - | - | - | LP | YES, a) |
| Genes related to cancer phenotypes | Hereditary breast and/or ovarian cancer | BRCA2 | AD | 13:32911488:-:TTATTG | - | 2021 | 1 | 0 | 0 | 1.0 | 0.0 | 0.0 | 1 | 0 | 0.049 | frameshift_variant | - | - | 4.090000152587891 | - | - | - | - | - | LP | YES, a) |
| Genes related to cancer phenotypes | Hereditary breast and/or ovarian cancer | BRCA2 | AD | 13:32912172:TG:- | rs80359395 | 2021 | 1 | 0 | 0 | 1.0 | 0.0 | 0.0 | 1 | 1 | 0.049 | frameshift_variant | - | - | -1.059999942779541 | - | - | - | 51504 | Pathogenic | P | YES, a) |
| Genes related to cancer phenotypes | Hereditary breast and/or ovarian cancer | BRCA2 | AD | 13:32912186:GAAG:- | - | 2021 | 1 | 0 | 0 | 1.0 | 0.0 | 0.0 | 1 | 0 | 0.049 | frameshift_variant | - | - | 3.0899999141693115 | - | - | - | - | - | LP | YES, a) |
| Genes related to cancer phenotypes | Hereditary breast and/or ovarian cancer | BRCA2 | AD | 13:32912194:-:C | - | 2021 | 1 | 0 | 0 | 1.0 | 0.0 | 0.0 | 1 | 0 | 0.049 | frameshift_variant | - | - | 4.610000133514404 | - | - | - | - | - | LP | YES, a) |
| Genes related to cancer phenotypes | Hereditary breast and/or ovarian cancer | BRCA2 | AD | 13:32912612:AA:- | - | 2021 | 1 | 0 | 0 | 1.0 | 0.0 | 0.0 | 1 | 0 | 0.049 | frameshift_variant | - | - | -4.949999809265137 | - | - | - | - | - | LP | YES, a) |
| Genes related to cancer phenotypes | Hereditary breast and/or ovarian cancer | BRCA2 | AD | 13:32912617:-:AG | - | 2021 | 1 | 0 | 0 | 1.0 | 0.0 | 0.0 | 1 | 0 | 0.049 | frameshift_variant | - | - | 3.2799999713897705 | - | - | - | - | - | LP | YES, a) |
| Genes related to cancer phenotypes | Hereditary breast and/or ovarian cancer | BRCA2 | AD | 13:32912904:-:CG | - | 2021 | 1 | 0 | 0 | 1.0 | 0.0 | 0.0 | 1 | 0 | 0.049 | frameshift_variant | - | - | 0.1080000028014183 | - | - | - | - | - | LP | YES, a) |
| Genes related to cancer phenotypes | Hereditary breast and/or ovarian cancer | BRCA2 | AD | 13:32913147:-:TACCT | - | 2021 | 1 | 0 | 0 | 1.0 | 0.0 | 0.0 | 1 | 0 | 0.049 | frameshift_variant | - | - | -5.260000228881836 | - | - | - | - | - | LP | YES, a) |
| Genes related to cancer phenotypes | Hereditary breast and/or ovarian cancer | BRCA2 | AD | 13:32913801:CTGG:- | - | 2021 | 1 | 0 | 0 | 1.0 | 0.0 | 0.0 | 1 | 0 | 0.049 | frameshift_variant | - | - | -0.33799999952316284 | - | - | - | - | - | LP | YES, a) |
| Genes related to cancer phenotypes | Hereditary breast and/or ovarian cancer | BRCA2 | AD | 13:32913806:-:CAAC | - | 2021 | 1 | 0 | 0 | 1.0 | 0.0 | 0.0 | 1 | 0 | 0.049 | frameshift_variant | - | - | 0.3930000066757202 | - | - | - | - | - | LP | YES, a) |
| Genes related to cancer phenotypes | Hereditary breast and/or ovarian cancer | BRCA2 | AD | 13:32914111:-:TC | - | 2021 | 1 | 0 | 0 | 1.0 | 0.0 | 0.0 | 1 | 0 | 0.049 | frameshift_variant | - | - | 2.7699999809265137 | - | - | - | - | - | LP | YES, a) |
| Genes related to cancer phenotypes | Hereditary breast and/or ovarian cancer | BRCA2 | AD | 13:32914112:-:T | - | 2021 | 1 | 0 | 0 | 1.0 | 0.0 | 0.0 | 1 | 0 | 0.049 | frameshift_variant | - | - | -0.9679999947547913 | - | - | - | 1071769 | Pathogenic | P | YES, a) |
| Genes related to cancer phenotypes | Hereditary breast and/or ovarian cancer | BRCA2 | AD | 13:32914767:TT:- | rs11571658 | 2021 | 1 | 0 | 0 | 1.0 | 0.0 | 0.0 | 1 | 1 | 0.049 | frameshift_variant | - | - | 1.3600000143051147 | - | - | - | 9318 | Pathogenic | P | YES, a) |
| Genes related to cancer phenotypes | Hereditary breast and/or ovarian cancer | BRCA2 | AD | 13:32914789:ACAAA:- | - | 2021 | 1 | 0 | 0 | 1.0 | 0.0 | 0.0 | 1 | 0 | 0.049 | frameshift_variant | - | - | 1.600000023841858 | - | - | - | - | - | LP | YES, a) |
| Genes related to cancer phenotypes | Hereditary breast and/or ovarian cancer | BRCA2 | AD | 13:32914795:-:GTTTT | - | 2021 | 1 | 0 | 0 | 1.0 | 0.0 | 0.0 | 1 | 0 | 0.049 | frameshift_variant | - | - | -1.6200000047683716 | - | - | - | - | - | LP | YES, a) |
| Genes related to cancer phenotypes | Hereditary breast and/or ovarian cancer | BRCA2 | AD | 13:32915052:-:A | - | 2021 | 1 | 0 | 0 | 1.0 | 0.0 | 0.0 | 1 | 0 | 0.049 | frameshift_variant | - | - | 4.070000171661377 | - | - | - | - | - | LP | YES, a) |
| Genes related to cancer phenotypes | Hereditary breast and/or ovarian cancer | BRCA2 | AD | 13:32969014:AGTC:- | - | 2021 | 1 | 0 | 0 | 1.0 | 0.0 | 0.0 | 1 | 0 | 0.049 | frameshift_variant | - | - | 5.889999866485596 | - | - | - | - | - | LP | YES, a) |
| Genes related to cancer phenotypes | Hereditary breast and/or ovarian cancer | BRCA2 | AD | 13:32969019:-:TGAA | - | 2021 | 1 | 0 | 0 | 1.0 | 0.0 | 0.0 | 1 | 0 | 0.049 | frameshift_variant | - | - | 5.889999866485596 | - | - | - | - | - | LP | YES, a) |
| Genes related to cancer phenotypes | Retinoblastoma | RB1 | AD | 13:49030368:AAAAA:- | - | 2021 | 1 | 0 | 0 | 1.0 | 0.0 | 0.0 | 1 | 0 | 0.049 | frameshift_variant | - | - | 4.800000190734863 | - | - | - | - | - | LP | YES, a) |
| Genes related to cancer phenotypes | Retinoblastoma | RB1 | AD | 13:49030373:-:GGGGG | - | 2021 | 1 | 0 | 0 | 1.0 | 0.0 | 0.0 | 1 | 0 | 0.049 | frameshift_variant | - | - | 4.800000190734863 | - | - | - | - | - | LP | YES, a) |
| Genes related to miscellaneous phenotypes | Wilson disease | ATP7B | AR | 13:52511697:G:A | rs758355520 | 2078 | 1 | 0 | 0 | 1.0 | 0.0 | 0.0 | 1 | 1 | 0.048 | missense_variant | - | - | 4.730000019073486 | 0.0 | 1.0 | 29.700000762939453 | 189139 | Pathogenic | P | NO, b) |
| Genes related to miscellaneous phenotypes | Wilson disease | ATP7B | AR | 13:52511706:T:C | rs121907990 | 2078 | 1 | 0 | 0 | 1.0 | 0.0 | 0.0 | 1 | 1 | 0.048 | missense_variant | - | - | 4.730000019073486 | 0.0 | 0.999 | 24.100000381469727 | 3859 | Pathogenic | P | NO, b) |
| Genes related to miscellaneous phenotypes | Wilson disease | ATP7B | AR | 13:52524267:C:T | rs775553302 | 2078 | 1 | 0 | 0 | 1.0 | 0.0 | 0.0 | 1 | 0 | 0.048 | missense_variant | - | - | 5.690000057220459 | 0.0 | 0.994 | 32.0 | 1525062 | Likely pathogenic | LP | NO, b) |
| Genes related to miscellaneous phenotypes | Wilson disease | ATP7B | AR | 13:52532466:C:T | rs137853283 | 2078 | 1 | 0 | 0 | 1.0 | 0.0 | 0.0 | 1 | 0 | 0.048 | stop_gained | - | - | 5.389999866485596 | - | - | 43.0 | 156284 | Pathogenic | P | NO, b) |
| Genes related to miscellaneous phenotypes | Wilson disease | ATP7B | AR | 13:52585454:TG:- | rs749363958 | 2078 | 1 | 0 | 0 | 1.0 | 0.0 | 0.0 | 1 | 0 | 0.048 | frameshift_variant | - | - | 2.869999885559082 | - | - | - | 419611 | Conflicting interpretations of p | LP | NO, b) |
| Genes related to cardiovascular phenotypes | Hypertrophic cardiomyopathy | MYH7 | AD | 14:23888731:C:T | rs730880906 | 2080 | 1 | 0 | 0 | 1.0 | 0.0 | 0.0 | 1 | 0 | 0.048 | missense_variant | - | - | 4.989999771118164 | 0.02 | 0.983 | 35.0 | 181385 | Conflicting interpretations of p | LP | YES, a) |
| Genes related to cardiovascular phenotypes | Hypertrophic cardiomyopathy | MYH7 | AD | 14:23899049:T:A | rs606231316 | 2036 | 1 | 0 | 0 | 1.0 | 0.0 | 0.0 | 1 | 1 | 0.049 | missense_variant | - | - | 3.940000057220459 | 0.0 | 0.996 | 28.700000762939453 | 132933 | Pathogenic | P | YES, a) |
| Genes related to cancer phenotypes | Hereditary paraganglioma–pheochromocytoma syndrome | MAX | AD | 14:65560530:CAGC:- | - | 2007 | 1 | 0 | 2 | 1.0 | 0.0 | 0.0 | 1 | 0 | 0.050 | frameshift_variant | - | - | 6.039999961853027 | - | - | - | - | - | LP | YES, a) |
| Genes related to cardiovascular phenotypes | Aortopathies | FBN1 | AD | 15:48719962:C:A | rs1199524458 | 2080 | 1 | 0 | 0 | 1.0 | 0.0 | 0.0 | 1 | 0 | 0.048 | stop_gained | - | - | 5.440000057220459 | - | - | 47.0 | 521084 | Pathogenic | P | YES, a) |
| Genes related to cardiovascular phenotypes | Aortopathies | FBN1 | AD | 15:48758044:-:T | - | 2080 | 1 | 0 | 0 | 1.0 | 0.0 | 0.0 | 1 | 0 | 0.048 | frameshift_variant | - | - | 5.739999771118164 | - | - | - | - | - | LP | YES, a) |
| Genes related to cardiovascular phenotypes | Aortopathies | FBN1 | AD | 15:48760618:-:A | - | 2080 | 1 | 0 | 0 | 1.0 | 0.0 | 0.0 | 1 | 1 | 0.048 | frameshift_variant | - | - | 5.489999771118164 | - | - | - | - | - | LP | YES, a) |
| Genes related to cardiovascular phenotypes | Aortopathies | SMAD3 | AD | 15:67457380:-:C | - | 2080 | 1 | 0 | 0 | 1.0 | 0.0 | 0.0 | 1 | 1 | 0.048 | frameshift_variant | - | - | 4.389999866485596 | - | - | - | - | - | LP | YES, a) |
| Genes related to cardiovascular phenotypes | Aortopathies | MYH11 | AD | 16:15815335:TTTTGTT | - | 2061 | 1 | 0 | 7 | 1.0 | 0.0 | 0.0 | 1 | 0 | 0.048 | frameshift_variant | - | - | 3.7200000286102295 | - | - | - | - | - | LP | YES, a) |
| Genes related to cardiovascular phenotypes | Aortopathies | MYH11 | AD | 16:15878561:AAAAG:- | - | 2080 | 1 | 0 | 0 | 1.0 | 0.0 | 0.0 | 1 | 0 | 0.048 | frameshift_variant | - | - | 5.489999771118164 | - | - | - | - | - | LP | YES, a) |
| Genes related to cardiovascular phenotypes | Aortopathies | MYH11 | AD | 16:15878567:-:CCCTC | - | 2080 | 1 | 0 | 0 | 1.0 | 0.0 | 0.0 | 1 | 0 | 0.048 | frameshift_variant | - | - | 0.6499999761581421 | - | - | - | - | - | LP | YES, a) |
| Genes related to cancer phenotypes | Tuberous sclerosis complex | TSC2 | AD | 16:2115609:CG:- | - | 2002 | 1 | 0 | 7 | 1.0 | 0.0 | 0.0 | 1 | 0 | 0.050 | frameshift_variant | - | - | 4.329999923706055 | - | - | - | - | - | LP | YES, a) |
| Genes related to cancer phenotypes | Tuberous sclerosis complex | TSC2 | AD | 16:2115614:-:TT | - | 2002 | 1 | 0 | 7 | 1.0 | 0.0 | 0.0 | 1 | 0 | 0.050 | frameshift_variant | - | - | 4.349999904632568 | - | - | - | - | - | LP | YES, a) |
| Genes related to cancer phenotypes | Tuberous sclerosis complex | TSC2 | AD | 16:2126142:C:T | rs45517258 | 2021 | 1 | 0 | 0 | 1.0 | 0.0 | 0.0 | 1 | 0 | 0.049 | missense_variant | - | - | 4.110000133514404 | 0.0 | 1.0 | 34.0 | 12404 | Pathogenic | P | YES, a) |
| Genes related to cancer phenotypes | Tuberous sclerosis complex | TSC2 | AD | 16:2138321:-:CAGCGG | rs759520580 | 2021 | 1 | 0 | 0 | 1.0 | 0.0 | 0.0 | 1 | 1 | 0.049 | inframe_insertion | - | - | -3.369999885559082 | - | - | - | - | - | LP | YES, a) |
| Genes related to cancer phenotypes | Hereditary breast and/or ovarian cancer | PALB2 | AD | 16:23625369:-:ACGA | - | 2021 | 1 | 0 | 0 | 1.0 | 0.0 | 0.0 | 1 | 0 | 0.049 | frameshift_variant | - | - | 4.860000133514404 | - | - | - | - | - | LP | YES, a) |
| Genes related to cancer phenotypes | Hereditary breast and/or ovarian cancer | PALB2 | AD | 16:23625372:CA:- | - | 2021 | 1 | 0 | 0 | 1.0 | 0.0 | 0.0 | 1 | 0 | 0.049 | frameshift_variant | - | - | -0.14300000667572021 | - | - | - | - | - | LP | YES, a) |
| Genes related to cancer phenotypes | Hereditary breast and/or ovarian cancer | PALB2 | AD | 16:23625383:T:- | rs1555458822 | 2021 | 1 | 0 | 0 | 1.0 | 0.0 | 0.0 | 1 | 0 | 0.049 | frameshift_variant | - | - | 5.949999809265137 | - | - | - | 430596 | Pathogenic | P | YES, a) |
| Genes related to cancer phenotypes | Hereditary breast and/or ovarian cancer | PALB2 | AD | 16:23646568:-:GA | - | 2021 | 1 | 0 | 0 | 1.0 | 0.0 | 0.0 | 1 | 0 | 0.049 | frameshift_variant | - | - | 3.6600000858306885 | - | - | - | - | - | LP | YES, a) |
| Genes related to cancer phenotypes | Hereditary breast and/or ovarian cancer | PALB2 | AD | 16:23646577:-:GA | - | 2021 | 1 | 0 | 0 | 1.0 | 0.0 | 0.0 | 1 | 0 | 0.049 | frameshift_variant | - | - | 5.670000076293945 | - | - | - | - | - | LP | YES, a) |
| Genes related to cancer phenotypes | Hereditary breast and/or ovarian cancer | BRCA1 | AD | 17:41246153:AT:- | - | 2021 | 1 | 0 | 0 | 1.0 | 0.0 | 0.0 | 1 | 0 | 0.049 | frameshift_variant | - | - | 4.570000171661377 | - | - | - | - | - | LP | YES, a) |
| Genes related to cancer phenotypes | Hereditary breast and/or ovarian cancer | BRCA1 | AD | 17:41246159:-:TG | - | 2021 | 1 | 0 | 0 | 1.0 | 0.0 | 0.0 | 1 | 0 | 0.049 | frameshift_variant | - | - | 3.3299999237060547 | - | - | - | - | - | LP | YES, a) |
| Genes related to cancer phenotypes | Hereditary breast and/or ovarian cancer | BRCA1 | AD | 17:41246204:GT:- | - | 2021 | 1 | 0 | 0 | 1.0 | 0.0 | 0.0 | 1 | 0 | 0.049 | frameshift_variant | - | - | -5.010000228881836 | - | - | - | - | - | LP | YES, a) |
| Genes related to cancer phenotypes | Hereditary breast and/or ovarian cancer | BRCA1 | AD | 17:41249289:CTTC:- | - | 2021 | 1 | 0 | 0 | 1.0 | 0.0 | 0.0 | 1 | 0 | 0.049 | frameshift_variant | - | - | 5.489999771118164 | - | - | - | - | - | LP | YES, a) |
| Genes related to cancer phenotypes | Hereditary breast and/or ovarian cancer | BRCA1 | AD | 17:41249305:-:C | - | 2021 | 1 | 0 | 0 | 1.0 | 0.0 | 0.0 | 1 | 0 | 0.049 | frameshift_variant | - | - | 4.960000038146973 | - | - | - | - | - | LP | YES, a) |
| Genes related to cancer phenotypes | Hereditary breast and/or ovarian cancer | BRCA1 | AD | 17:41249306:-:CG | - | 2021 | 1 | 0 | 0 | 1.0 | 0.0 | 0.0 | 1 | 0 | 0.049 | frameshift_variant | - | - | 1.4600000381469727 | - | - | - | - | - | LP | YES, a) |
| Genes related to cancer phenotypes | Hereditary breast and/or ovarian cancer | BRCA1 | AD | 17:41256219:CA:- | - | 2021 | 1 | 0 | 0 | 1.0 | 0.0 | 0.0 | 1 | 0 | 0.049 | frameshift_variant | - | - | -3.0799999237060547 | - | - | - | - | - | LP | YES, a) |
| Genes related to cancer phenotypes | Hereditary breast and/or ovarian cancer | BRCA1 | AD | 17:41256222:-:G | - | 2021 | 1 | 0 | 0 | 1.0 | 0.0 | 0.0 | 1 | 0 | 0.049 | frameshift_variant | - | - | 5.320000171661377 | - | - | - | - | - | LP | YES, a) |
| Genes related to cancer phenotypes | Hereditary breast and/or ovarian cancer | BRCA1 | AD | 17:41256224:-:C | - | 2021 | 1 | 0 | 0 | 1.0 | 0.0 | 0.0 | 1 | 0 | 0.049 | frameshift_variant | - | - | -0.5109999775886536 | - | - | - | - | - | LP | YES, a) |
| Genes related to cancer phenotypes | Hereditary breast and/or ovarian cancer | BRCA1 | AD | 17:41258474:T:C | rs80357382 | 2021 | 1 | 0 | 0 | 1.0 | 0.0 | 0.0 | 1 | 0 | 0.049 | missense_variant | - | - | 4.639999866485596 | 0.0 | 0.985 | 23.5 | 17693 | Pathogenic | P | YES, a) |
| Genes related to cancer phenotypes | Li–Fraumeni syndrome | TP53 | AD | 17:7577035:TGGGG:- | - | 2021 | 1 | 0 | 0 | 1.0 | 0.0 | 0.0 | 1 | 0 | 0.049 | frameshift_variant | - | - | 4.099999904632568 | - | - | - | - | - | LP | YES, a) |
| Genes related to cancer phenotypes | Li–Fraumeni syndrome | TP53 | AD | 17:7579313:G:A | rs786201057 | 2021 | 1 | 0 | 0 | 1.0 | 0.0 | 0.0 | 1 | 0 | 0.049 | missense_variant | - | - | 4.75 | 0.04 | 1.0 | 33.0 | 183748 | Conflicting interpretations of p | LP | YES, a) |
| Genes related to inborn errors of metabolism phenotypes | Pompe disease | GAA | AR | 17:78081665:G:A | rs543300039 | 2078 | 1 | 0 | 0 | 1.0 | 0.0 | 0.0 | 1 | 1 | 0.048 | missense_variant | 0.0009940357 | 0.004672897 | 5.289999961853027 | 0.0 | 1.0 | 34.0 | 188797 | Pathogenic | P | NO, b) |
| Genes related to inborn errors of metabolism phenotypes | Pompe disease | GAA | AR | 17:78090814:G:C | rs752921215 | 2078 | 1 | 0 | 0 | 1.0 | 0.0 | 0.0 | 1 | 0 | 0.048 | missense_variant | - | - | 5.099999904632568 | 0.0 | 0.999 | 33.0 | 188484 | Pathogenic/Likely pathogenic | P/LP | NO, b) |
| Genes related to cardiovascular phenotypes | Arrhythmogenic right ventricular cardiomyopathy | DSC2 | AD | 18:28649032:CC:- | - | 2080 | 1 | 0 | 0 | 1.0 | 0.0 | 0.0 | 1 | 0 | 0.048 | frameshift_variant | - | - | 4.309999942779541 | - | - | - | - | - | LP | YES, a) |
| Genes related to cardiovascular phenotypes | Arrhythmogenic right ventricular cardiomyopathy | DSC2 | AD | 18:28649035:-:GAAAT | - | 2080 | 1 | 0 | 0 | 1.0 | 0.0 | 0.0 | 1 | 0 | 0.048 | frameshift_variant | - | - | -6.010000228881836 | - | - | - | - | - | LP | YES, a) |
| Genes related to cardiovascular phenotypes | Arrhythmogenic right ventricular cardiomyopathy | DSC2 | AD | 18:28659882:T:- | - | 2080 | 1 | 0 | 0 | 1.0 | 0.0 | 0.0 | 1 | 0 | 0.048 | frameshift_variant | - | - | 1.5800000429153442 | - | - | - | - | - | LP | YES, a) |
| Genes related to cardiovascular phenotypes | Arrhythmogenic right ventricular cardiomyopathy | DSC2 | AD | 18:28666552:-:A | - | 2080 | 1 | 0 | 0 | 1.0 | 0.0 | 0.0 | 1 | 0 | 0.048 | frameshift_variant | - | - | 2.799999952316284 | - | - | - | - | - | LP | YES, a) |
| Genes related to cardiovascular phenotypes | Arrhythmogenic right ventricular cardiomyopathy | DSG2 | AD | 18:29098200:A:C | - | 1108 | 1 | 0 | 0 | 1.0 | 0.0 | 0.0 | 1 | 1 | 0.090 | splice_acceptor_variant | - | - | 5.070000171661377 | - | - | 23.899999618530273 | - | - | P | YES, a) |
| Genes related to cardiovascular phenotypes | Arrhythmogenic right ventricular cardiomyopathy | DSG2 | AD | 18:29102213:G:A | rs750176752 | 971 | 1 | 0 | 0 | 0.999 | 0.001 | 0.001 | 1 | 1 | 0.103 | splice_donor_variant | - | - | 5.53000020980835 | 0.62 | 0.0 | 27.100000381469727 | 410373 | Pathogenic | P | YES, a) |
| Genes related to cardiovascular phenotypes | Arrhythmogenic right ventricular cardiomyopathy | DSG2 | AD | 18:29115245:AGAAG:- | - | 2080 | 1 | 0 | 0 | 1.0 | 0.0 | 0.0 | 1 | 0 | 0.048 | frameshift_variant | - | - | 5.659999847412109 | - | - | - | - | - | LP | YES, a) |
| Genes related to cardiovascular phenotypes | Arrhythmogenic right ventricular cardiomyopathy | DSG2 | AD | 18:29115251:-:TCCTC | - | 2080 | 1 | 0 | 0 | 1.0 | 0.0 | 0.0 | 1 | 0 | 0.048 | frameshift_variant | - | - | 5.659999847412109 | - | - | - | - | - | LP | YES, a) |
| Genes related to miscellaneous phenotypes | Hereditary transthyretin-related amyloidosis | TTR | AD | 18:29172937:G:A | rs28933979 | 2078 | 1 | 0 | 0 | 1.0 | 0.0 | 0.0 | 1 | 0 | 0.048 | missense_variant | - | - | 5.710000038146973 | 0.01 | 0.966 | 25.299999237060547 | 13417 | Pathogenic | P | YES, a) |
| Genes related to cancer phenotypes | Juvenile polyposis syndrome | SMAD4 | AD | 18:48591833:T:- | - | 2021 | 1 | 0 | 0 | 1.0 | 0.0 | 0.0 | 1 | 0 | 0.049 | frameshift_variant | - | - | 2.25 | - | - | - | - | - | LP | YES, a) |
| Genes related to cancer phenotypes | Juvenile polyposis syndrome | SMAD4 | AD | 18:48591835:-:CA | - | 2021 | 1 | 0 | 0 | 1.0 | 0.0 | 0.0 | 1 | 0 | 0.049 | frameshift_variant | - | - | 5.989999771118164 | - | - | - | - | - | LP | YES, a) |
| Genes related to cardiovascular phenotypes | Familial hypercholesterolemia | LDLR | AD | 19:11213340:T:A | rs774016801 | 2078 | 1 | 0 | 0 | 1.0 | 0.0 | 0.0 | 1 | 1 | 0.048 | stop_gained | - | - | -8.15999984741211 | - | - | 23.0 | 375776 | Pathogenic | P | YES, a) |
| Genes related to cardiovascular phenotypes | Familial hypercholesterolemia | LDLR | AD | 19:11215925:C:T | rs774723292 | 2078 | 1 | 0 | 0 | 1.0 | 0.0 | 0.0 | 1 | 1 | 0.048 | missense_variant | - | - | 5.329999923706055 | 0.01 | 0.984 | 33.0 | 251162 | Likely pathogenic | LP | YES, a) |
| Genes related to cardiovascular phenotypes | Familial hypercholesterolemia | LDLR | AD | 19:11224082:A:T | rs1057519670 | 2078 | 1 | 0 | 0 | 1.0 | 0.0 | 0.0 | 1 | 0 | 0.048 | missense_variant | - | - | 4.869999885559082 | 0.01 | 0.992 | 27.5 | 375813 | Likely pathogenic | LP | YES, a) |
| Genes related to cardiovascular phenotypes | Familial hypercholesterolemia | LDLR | AD | 19:11231137:G:A | rs551528700 | 2078 | 1 | 0 | 0 | 1.0 | 0.0 | 0.0 | 1 | 1 | 0.048 | synonymous_variant | 0.0009940357 | 0.004672897 | 2.740000009536743 | - | - | 9.770000457763672 | 430794 | Likely pathogenic | LP | YES, a) |
| Genes related to cancer phenotypes | Peutz–Jeghers syndrome | STK11 | AD | 19:1207153:A:- | rs1599915235 | 2021 | 1 | 0 | 0 | 1.0 | 0.0 | 0.0 | 1 | 0 | 0.049 | frameshift_variant | - | - | 2.859999895095825 | - | - | - | 821260 | Pathogenic | P | YES, a) |
| Genes related to cancer phenotypes | Peutz–Jeghers syndrome | STK11 | AD | 19:1207157:-:G | - | 2021 | 1 | 0 | 0 | 1.0 | 0.0 | 0.0 | 1 | 0 | 0.049 | frameshift_variant | - | - | 3.9000000953674316 | - | - | - | - | - | LP | YES, a) |
| Genes related to miscellaneous phenotypes | Malignant hyperthermia | RYR1 | AD | 19:38993222:A:- | - | 2095 | 1 | 0 | 0 | 1.0 | 0.0 | 0.0 | 1 | 0 | 0.048 | frameshift_variant | - | - | 3.990000009536743 | - | - | - | - | - | LP | YES, a) |
| Genes related to miscellaneous phenotypes | Malignant hyperthermia | RYR1 | AD | 19:38993226:-:G | - | 2095 | 1 | 0 | 0 | 1.0 | 0.0 | 0.0 | 1 | 0 | 0.048 | frameshift_variant | - | - | 3.990000009536743 | - | - | - | - | - | LP | YES, a) |
| Genes related to miscellaneous phenotypes | Malignant hyperthermia | RYR1 | AD | 19:38993313:C:A | - | 2095 | 1 | 0 | 0 | 1.0 | 0.0 | 0.0 | 1 | 0 | 0.048 | stop_gained | - | - | 4.079999923706055 | - | - | 42.0 | - | - | P | YES, a) |
| Genes related to miscellaneous phenotypes | Malignant hyperthermia | RYR1 | AD | 19:39015987:C:T | - | 1439 | 1 | 0 | 0 | 1.0 | 0.0 | 0.0 | 1 | 1 | 0.069 | stop_gained | - | - | 4.300000190734863 | - | - | 55.0 | 1073108 | Pathogenic | P | YES, a) |
| Genes related to miscellaneous phenotypes | Malignant hyperthermia | RYR1 | AD | 19:39026643:-:TA | - | 2095 | 1 | 0 | 0 | 1.0 | 0.0 | 0.0 | 1 | 0 | 0.048 | frameshift_variant | - | - | 4.380000114440918 | - | - | - | - | - | LP | YES, a) |
| Genes related to miscellaneous phenotypes | Malignant hyperthermia | RYR1 | AD | 19:39055617:C:T | - | 2095 | 1 | 0 | 0 | 1.0 | 0.0 | 0.0 | 1 | 0 | 0.048 | stop_gained | - | - | 3.069999933242798 | - | - | 57.0 | - | - | P | YES, a) |
| Genes related to miscellaneous phenotypes | Malignant hyperthermia | RYR1 | AD | 19:39070656:CC:- | - | 2095 | 1 | 0 | 0 | 1.0 | 0.0 | 0.0 | 1 | 0 | 0.048 | frameshift_variant | - | - | -1.440000057220459 | - | - | - | - | - | LP | YES, a) |

| | | | | | | | | | | | | | | | | | | | | | | | | | | |
|---|---|---|---|---|---|---|---|---|---|---|---|---|---|---|---|---|---|---|---|---|---|---|---|---|---|---|
| Genes related to miscellaneous phenotypes | Malignant hyperthermia | RYR1 | AD | 19:39077205:-:GTCTT | - | 2095 | 1 | 0 | 0 | 1.0 | 0.0 | 0.0 | 1 | 0 | 0.048 | frameshift_variant | - | - | 1.190000057220459 | - | - | - | - | - | LP | YES, a) |
| Genes related to cardiovascular phenotypes | Dilated cardiomyopathy | TTN | AD | 2:179399104:CT:- | - | 2080 | 1 | 0 | 0 | 1.0 | 0.0 | 0.0 | 1 | 1 | 0.048 | frameshift_variant | - | - | -4.199999809265137 | - | - | - | - | - | LP | YES, a) |
| Genes related to cardiovascular phenotypes | Dilated cardiomyopathy | TTN | AD | 2:179418514:T:A | - | 2080 | 1 | 0 | 0 | 1.0 | 0.0 | 0.0 | 1 | 1 | 0.048 | stop_gained | - | - | 5.610000133514404 | - | - | 67.0 | - | - | P | YES, a) |
| Genes related to cardiovascular phenotypes | Dilated cardiomyopathy | TTN | AD | 2:179421842:T:- | - | 2080 | 1 | 0 | 0 | 1.0 | 0.0 | 0.0 | 1 | 0 | 0.048 | frameshift_variant | - | - | 5.809999942779541 | - | - | - | - | - | LP | YES, a) |
| Genes related to cardiovascular phenotypes | Dilated cardiomyopathy | TTN | AD | 2:179429537:-:C | - | 2080 | 1 | 0 | 0 | 1.0 | 0.0 | 0.0 | 1 | 0 | 0.048 | frameshift_variant | - | - | 6.019999980926514 | - | - | - | - | - | LP | YES, a) |
| Genes related to cardiovascular phenotypes | Dilated cardiomyopathy | TTN | AD | 2:179429538:-:T | - | 2080 | 1 | 0 | 0 | 1.0 | 0.0 | 0.0 | 1 | 0 | 0.048 | frameshift_variant | - | - | 5.130000114440918 | - | - | - | - | - | LP | YES, a) |
| Genes related to cardiovascular phenotypes | Dilated cardiomyopathy | TTN | AD | 2:179429538:G:T | rs557312035 | 2080 | 1 | 0 | 0 | 1.0 | 0.0 | 0.0 | 1 | 1 | 0.048 | stop_gained | 0.0009940357 | 0.004672897 | 4.21999979019165 | - | - | 65.0 | - | - | P | YES, a) |
| Genes related to cardiovascular phenotypes | Dilated cardiomyopathy | TTN | AD | 2:179449550:-:TGGGT | - | 2080 | 1 | 0 | 0 | 1.0 | 0.0 | 0.0 | 1 | 0 | 0.048 | frameshift_variant | - | - | 6.170000076293945 | - | - | - | - | - | LP | YES, a) |
| Genes related to cardiovascular phenotypes | Dilated cardiomyopathy | TTN | AD | 2:179458065:G:A | rs1553649171 | 2080 | 1 | 0 | 0 | 1.0 | 0.0 | 0.0 | 1 | 0 | 0.048 | stop_gained | - | - | 3.0799999237060547 | - | - | 62.0 | 466646 | Pathogenic/Likely pathogenic | P/LP | YES, a) |
| Genes related to cardiovascular phenotypes | Dilated cardiomyopathy | TTN | AD | 2:179462513:CTGACAC | - | 2080 | 1 | 0 | 0 | 1.0 | 0.0 | 0.0 | 1 | 0 | 0.048 | frameshift_variant | - | - | 2.759999990463257 | - | - | - | - | - | LP | YES, a) |
| Genes related to cardiovascular phenotypes | Dilated cardiomyopathy | TTN | AD | 2:179469713:C:T | rs756339648 | 1836 | 1 | 0 | 0 | 1.0 | 0.0 | 0.0 | 1 | 0 | 0.054 | splice_donor_variant | - | - | 5.949999809265137 | - | - | 24.100000381469727 | 287661 | Likely pathogenic | LP | YES, a) |
| Genes related to cardiovascular phenotypes | Dilated cardiomyopathy | TTN | AD | 2:179471777:-:GG | - | 2064 | 1 | 0 | 4 | 1.0 | 0.0 | 0.0 | 1 | 0 | 0.048 | frameshift_variant | - | - | 4.159999847412109 | - | - | - | - | - | LP | YES, a) |
| Genes related to cardiovascular phenotypes | Dilated cardiomyopathy | TTN | AD | 2:179471779:-:A | - | 2064 | 1 | 0 | 4 | 1.0 | 0.0 | 0.0 | 1 | 0 | 0.048 | frameshift_variant | - | - | 5.349999904632568 | - | - | - | - | - | LP | YES, a) |
| Genes related to cardiovascular phenotypes | Dilated cardiomyopathy | TTN | AD | 2:179474991:C:- | - | 2080 | 1 | 0 | 0 | 1.0 | 0.0 | 0.0 | 1 | 0 | 0.048 | frameshift_variant | - | - | 5.630000114440918 | - | - | - | - | - | LP | YES, a) |
| Genes related to cardiovascular phenotypes | Dilated cardiomyopathy | TTN | AD | 2:179474995:-:C | - | 2080 | 1 | 0 | 0 | 1.0 | 0.0 | 0.0 | 1 | 0 | 0.048 | frameshift_variant | - | - | 5.630000114440918 | - | - | - | - | - | LP | YES, a) |
| Genes related to cardiovascular phenotypes | Dilated cardiomyopathy | TTN | AD | 2:179485989:C:T | - | 2080 | 1 | 0 | 0 | 1.0 | 0.0 | 0.0 | 1 | 0 | 0.048 | stop_gained | - | - | 6.170000076293945 | - | - | 62.0 | - | - | P | YES, a) |
| Genes related to cardiovascular phenotypes | Dilated cardiomyopathy | TTN | AD | 2:179497711:CCAG:- | - | 2080 | 1 | 0 | 0 | 1.0 | 0.0 | 0.0 | 1 | 1 | 0.048 | frameshift_variant | - | - | 3.9600000381469727 | - | - | - | - | - | LP | YES, a) |
| Genes related to cardiovascular phenotypes | Dilated cardiomyopathy | TTN | AD | 2:179498145:-:GGAGT | - | 2080 | 1 | 0 | 0 | 1.0 | 0.0 | 0.0 | 1 | 0 | 0.048 | frameshift_variant | - | - | -4.289999961853027 | - | - | - | - | - | LP | YES, a) |
| Genes related to cardiovascular phenotypes | Dilated cardiomyopathy | TTN | AD | 2:179500946:-:G | - | 2080 | 1 | 0 | 0 | 1.0 | 0.0 | 0.0 | 1 | 0 | 0.048 | frameshift_variant | - | - | 5.619999885559082 | - | - | - | - | - | LP | YES, a) |
| Genes related to cardiovascular phenotypes | Dilated cardiomyopathy | TTN | AD | 2:179526487:T:- | - | 833 | 1 | 0 | 0 | 0.999 | 0.001 | 0.001 | 1 | 1 | 0.120 | frameshift_variant | - | - | 4.320000171661377 | - | - | - | - | - | LP | YES, a) |
| Genes related to cardiovascular phenotypes | Dilated cardiomyopathy | TTN | AD | 2:179532392:G:A | - | 1775 | 1 | 0 | 0 | 1.0 | 0.0 | 0.0 | 1 | 1 | 0.056 | stop_gained | - | - | 3.799999952316284 | - | - | 58.0 | - | - | P | YES, a) |
| Genes related to cardiovascular phenotypes | Dilated cardiomyopathy | TTN | AD | 2:179539765:C:T | rs577363824 | 2080 | 1 | 0 | 0 | 1.0 | 0.0 | 0.0 | 1 | 1 | 0.048 | splice_donor_variant | - | - | 5.630000114440918 | - | - | 25.600000381469727 | 404922 | Conflicting interpretations of p | P | YES, a) |
| Genes related to cardiovascular phenotypes | Dilated cardiomyopathy | TTN | AD | 2:179548772:T:- | - | 2080 | 1 | 0 | 0 | 1.0 | 0.0 | 0.0 | 1 | 0 | 0.048 | frameshift_variant | - | - | 0.013299999758601189 | - | - | - | - | - | LP | YES, a) |
| Genes related to cardiovascular phenotypes | Dilated cardiomyopathy | TTN | AD | 2:179548779:-:C | - | 2080 | 1 | 0 | 0 | 1.0 | 0.0 | 0.0 | 1 | 0 | 0.048 | frameshift_variant | - | - | 5.579999923706055 | - | - | - | - | - | LP | YES, a) |
| Genes related to cardiovascular phenotypes | Dilated cardiomyopathy | TTN | AD | 2:179549674:GG:- | - | 2080 | 1 | 0 | 0 | 1.0 | 0.0 | 0.0 | 1 | 0 | 0.048 | frameshift_variant | - | - | 5.730000019073486 | - | - | - | - | - | LP | YES, a) |
| Genes related to cardiovascular phenotypes | Dilated cardiomyopathy | TTN | AD | 2:179549678:-:GG | - | 2080 | 1 | 0 | 0 | 1.0 | 0.0 | 0.0 | 1 | 0 | 0.048 | frameshift_variant | - | - | 3.380000114440918 | - | - | - | - | - | LP | YES, a) |
| Genes related to cardiovascular phenotypes | Dilated cardiomyopathy | TTN | AD | 2:179557260:GAAC:- | - | 2080 | 1 | 0 | 0 | 1.0 | 0.0 | 0.0 | 1 | 0 | 0.048 | frameshift_variant | - | - | 5.739999771118164 | - | - | - | - | - | LP | YES, a) |
| Genes related to cardiovascular phenotypes | Dilated cardiomyopathy | TTN | AD | 2:179557265:-:GGTG | - | 2080 | 1 | 0 | 0 | 1.0 | 0.0 | 0.0 | 1 | 0 | 0.048 | frameshift_variant | - | - | 3.240000009536743 | - | - | - | - | - | LP | YES, a) |
| Genes related to cardiovascular phenotypes | Dilated cardiomyopathy | TTN | AD | 2:179580231:A:C | - | 2024 | 1 | 0 | 0 | 1.0 | 0.0 | 0.0 | 1 | 1 | 0.049 | stop_gained | - | - | 5.480000019073486 | - | - | 59.0 | - | - | P | YES, a) |
| Genes related to cardiovascular phenotypes | Dilated cardiomyopathy | TTN | AD | 2:179582869:AT:- | - | 2080 | 1 | 0 | 0 | 1.0 | 0.0 | 0.0 | 1 | 0 | 0.048 | frameshift_variant | - | - | 5.059999942779541 | - | - | - | - | - | LP | YES, a) |
| Genes related to cardiovascular phenotypes | Dilated cardiomyopathy | TTN | AD | 2:179582874:CT:- | - | 2080 | 1 | 0 | 0 | 1.0 | 0.0 | 0.0 | 1 | 0 | 0.048 | frameshift_variant | - | - | 3.8299999237060547 | - | - | - | - | - | LP | YES, a) |
| Genes related to cardiovascular phenotypes | Dilated cardiomyopathy | TTN | AD | 2:179587389:G:A | - | 2080 | 1 | 0 | 0 | 1.0 | 0.0 | 0.0 | 1 | 0 | 0.048 | stop_gained | - | - | 5.949999809265137 | - | - | 46.0 | - | - | P | YES, a) |
| Genes related to cardiovascular phenotypes | Dilated cardiomyopathy | TTN | AD | 2:179587478:-:CT | - | 2080 | 0 | 1 | 0 | 1.0 | 0.0 | 0.0 | 1 | 1 | 0.048 | frameshift_variant | - | - | 3.569999933242798 | - | - | - | - | - | LP | YES, a) |
| Genes related to cardiovascular phenotypes | Dilated cardiomyopathy | TTN | AD | 2:179590368:T:- | - | 2080 | 1 | 0 | 0 | 1.0 | 0.0 | 0.0 | 1 | 0 | 0.048 | frameshift_variant | - | - | 4.699999809265137 | - | - | - | - | - | LP | YES, a) |
| Genes related to cardiovascular phenotypes | Dilated cardiomyopathy | TTN | AD | 2:179604789:G:A | - | 2080 | 1 | 0 | 0 | 1.0 | 0.0 | 0.0 | 1 | 0 | 0.048 | stop_gained | - | - | 1.2699999809265137 | - | - | 33.0 | - | - | LP | YES, a) |
| Genes related to cardiovascular phenotypes | Dilated cardiomyopathy | TTN | AD | 2:179611187:-:CT | - | 2080 | 1 | 0 | 0 | 1.0 | 0.0 | 0.0 | 1 | 0 | 0.048 | frameshift_variant | - | - | 5.880000114440918 | - | - | - | - | - | LP | YES, a) |
| Genes related to cardiovascular phenotypes | Dilated cardiomyopathy | TTN | AD | 2:179611194:-:A | - | 2080 | 1 | 0 | 0 | 1.0 | 0.0 | 0.0 | 1 | 0 | 0.048 | frameshift_variant | - | - | 2.930000066757202 | - | - | - | - | - | LP | YES, a) |
| Genes related to cardiovascular phenotypes | Dilated cardiomyopathy | TTN | AD | 2:179613050:-:GT | - | 2080 | 1 | 0 | 0 | 1.0 | 0.0 | 0.0 | 1 | 0 | 0.048 | frameshift_variant | - | - | -1.7300000190734863 | - | - | - | - | - | LP | YES, a) |
| Genes related to cardiovascular phenotypes | Dilated cardiomyopathy | TTN | AD | 2:179613052:-:T | - | 2080 | 1 | 0 | 0 | 1.0 | 0.0 | 0.0 | 1 | 0 | 0.048 | frameshift_variant | - | - | -8.789999961853027 | - | - | - | - | - | LP | YES, a) |
| Genes related to cardiovascular phenotypes | Dilated cardiomyopathy | TTN | AD | 2:179613198:TATTT:- | - | 2080 | 1 | 0 | 0 | 1.0 | 0.0 | 0.0 | 1 | 0 | 0.048 | frameshift_variant | - | - | 4.730000019073486 | - | - | - | - | - | LP | YES, a) |
| Genes related to cardiovascular phenotypes | Dilated cardiomyopathy | TTN | AD | 2:179613203:-:CGCCC | - | 2080 | 1 | 0 | 0 | 1.0 | 0.0 | 0.0 | 1 | 0 | 0.048 | frameshift_variant | - | - | 4.730000019073486 | - | - | - | - | - | LP | YES, a) |
| Genes related to cardiovascular phenotypes | Dilated cardiomyopathy | TTN | AD | 2:179613629:TTTG:- | - | 2080 | 1 | 0 | 0 | 1.0 | 0.0 | 0.0 | 1 | 0 | 0.048 | frameshift_variant | - | - | 4.21999979019165 | - | - | - | - | - | LP | YES, a) |
| Genes related to cardiovascular phenotypes | Dilated cardiomyopathy | TTN | AD | 2:179613634:-:CCAT | - | 2080 | 1 | 0 | 0 | 1.0 | 0.0 | 0.0 | 1 | 0 | 0.048 | frameshift_variant | - | - | -3.059999942779541 | - | - | - | - | - | LP | YES, a) |
| Genes related to cardiovascular phenotypes | Dilated cardiomyopathy | TTN | AD | 2:179614145:-:CTCCT | - | 2080 | 1 | 0 | 0 | 1.0 | 0.0 | 0.0 | 1 | 0 | 0.048 | frameshift_variant | - | - | 0.11900000274181366 | - | - | - | - | - | LP | YES, a) |
| Genes related to cardiovascular phenotypes | Dilated cardiomyopathy | TTN | AD | 2:179615904:A:- | - | 2080 | 1 | 0 | 0 | 1.0 | 0.0 | 0.0 | 1 | 0 | 0.048 | frameshift_variant | - | - | -3.549999952316284 | - | - | - | - | - | LP | YES, a) |
| Genes related to cardiovascular phenotypes | Dilated cardiomyopathy | TTN | AD | 2:179616345:T:- | rs777963995 | 2080 | 1 | 0 | 0 | 1.0 | 0.0 | 0.0 | 1 | 1 | 0.048 | frameshift_variant | - | - | 2.309999942779541 | - | - | - | - | - | LP | YES, a) |
| Genes related to cardiovascular phenotypes | Dilated cardiomyopathy | TTN | AD | 2:179621020:-:C | rs778172350 | 2080 | 1 | 0 | 0 | 1.0 | 0.0 | 0.0 | 1 | 1 | 0.048 | frameshift_variant | - | - | 2.319999933242798 | - | - | - | 202507 | Conflicting interpretations of p | LP | YES, a) |
| Genes related to cardiovascular phenotypes | Dilated cardiomyopathy | TTN | AD | 2:179633535:-:CGGGT | - | 2080 | 1 | 0 | 0 | 1.0 | 0.0 | 0.0 | 1 | 0 | 0.048 | frameshift_variant | - | - | 5.559999942779541 | - | - | - | - | - | LP | YES, a) |
| Genes related to cardiovascular phenotypes | Dilated cardiomyopathy | TTN | AD | 2:179641516:C:- | - | 2080 | 1 | 0 | 0 | 1.0 | 0.0 | 0.0 | 1 | 0 | 0.048 | frameshift_variant | - | - | 5.329999923706055 | - | - | - | - | - | LP | YES, a) |
| Genes related to cardiovascular phenotypes | Dilated cardiomyopathy | TTN | AD | 2:179641522:-:C | - | 2080 | 1 | 0 | 0 | 1.0 | 0.0 | 0.0 | 1 | 0 | 0.048 | frameshift_variant | - | - | 1.649999976158142 | - | - | - | - | - | LP | YES, a) |
| Genes related to cardiovascular phenotypes | Dilated cardiomyopathy | TTN | AD | 2:179644933:C:A | - | 958 | 1 | 0 | 0 | 0.999 | 0.001 | 0.001 | 1 | 1 | 0.104 | splice_acceptor_variant | - | - | 5.789999961853027 | - | - | 27.899999618530273 | - | - | P | YES, a) |
| Genes related to cardiovascular phenotypes | Dilated cardiomyopathy | TTN | AD | 2:179645859:AAGG:- | - | 2080 | 1 | 0 | 0 | 1.0 | 0.0 | 0.0 | 1 | 1 | 0.048 | frameshift_variant | - | - | 5.909999847412109 | - | - | - | - | - | LP | YES, a) |
| Genes related to cardiovascular phenotypes | Dilated cardiomyopathy | TTN | AD | 2:179647754:ACAGA:- | - | 2080 | 1 | 0 | 0 | 1.0 | 0.0 | 0.0 | 1 | 0 | 0.048 | frameshift_variant | - | - | 6.170000076293945 | - | - | - | - | - | LP | YES, a) |
| Genes related to cardiovascular phenotypes | Dilated cardiomyopathy | TTN | AD | 2:179647759:-:TGTCT | - | 2080 | 1 | 0 | 0 | 1.0 | 0.0 | 0.0 | 1 | 0 | 0.048 | frameshift_variant | - | - | 6.170000076293945 | - | - | - | - | - | LP | YES, a) |
| Genes related to cardiovascular phenotypes | Dilated cardiomyopathy | TTN | AD | 2:179650383:TGAAACA | - | 2036 | 1 | 0 | 0 | 1.0 | 0.0 | 0.0 | 1 | 0 | 0.049 | frameshift_variant | - | - | -11.5 | - | - | - | - | - | LP | YES, a) |
| Genes related to cardiovascular phenotypes | Ehlers–Danlos syndrome, vascular type | COL3A1 | AD | 2:189850397:CCTGG:- | - | 2080 | 1 | 0 | 0 | 1.0 | 0.0 | 0.0 | 1 | 0 | 0.048 | frameshift_variant | - | - | 4.619999885559082 | - | - | - | - | - | LP | YES, a) |
| Genes related to cardiovascular phenotypes | Ehlers–Danlos syndrome, vascular type | COL3A1 | AD | 2:189850402:-:TTCAA | - | 2080 | 1 | 0 | 0 | 1.0 | 0.0 | 0.0 | 1 | 0 | 0.048 | frameshift_variant | - | - | 4.619999885559082 | - | - | - | - | - | LP | YES, a) |
| Genes related to cardiovascular phenotypes | Ehlers–Danlos syndrome, vascular type | COL3A1 | AD | 2:189855741:ACGAA:- | - | 2080 | 1 | 0 | 0 | 1.0 | 0.0 | 0.0 | 1 | 0 | 0.048 | frameshift_variant | - | - | 4.840000152587891 | - | - | - | - | - | LP | YES, a) |
| Genes related to cardiovascular phenotypes | Ehlers–Danlos syndrome, vascular type | COL3A1 | AD | 2:189855747:-:GCTTT | - | 2080 | 1 | 0 | 0 | 1.0 | 0.0 | 0.0 | 1 | 0 | 0.048 | frameshift_variant | - | - | 5.980000019073486 | - | - | - | - | - | LP | YES, a) |
| Genes related to cardiovascular phenotypes | Ehlers–Danlos syndrome, vascular type | COL3A1 | AD | 2:189860906:T:G | - | 1108 | 1 | 0 | 0 | 1.0 | 0.0 | 0.0 | 1 | 1 | 0.090 | splice_donor_variant | - | - | 5.610000133514404 | - | - | 25.100000381469727 | - | - | P | YES, a) |
| Genes related to cardiovascular phenotypes | Ehlers–Danlos syndrome, vascular type | COL3A1 | AD | 2:189864064:-:AA | - | 2068 | 1 | 0 | 0 | 1.0 | 0.0 | 0.0 | 1 | 0 | 0.048 | frameshift_variant | - | - | 4.059999942779541 | - | - | - | - | - | LP | YES, a) |
| Genes related to cardiovascular phenotypes | Ehlers–Danlos syndrome, vascular type | COL3A1 | AD | 2:189864075:-:A | - | 2066 | 1 | 0 | 2 | 1.0 | 0.0 | 0.0 | 1 | 0 | 0.048 | frameshift_variant | - | - | 4.960000038146973 | - | - | - | - | - | LP | YES, a) |
| Genes related to cardiovascular phenotypes | Familial hypercholesterolemia | APOB | AD | 2:21227953:ATTT:- | - | 2032 | 1 | 0 | 0 | 1.0 | 0.0 | 0.0 | 1 | 0 | 0.049 | frameshift_variant | - | - | -7.119999885559082 | - | - | - | - | - | LP | YES, a) |
| Genes related to cardiovascular phenotypes | Familial hypercholesterolemia | APOB | AD | 2:21227959:-:GGCT | - | 2078 | 1 | 0 | 0 | 1.0 | 0.0 | 0.0 | 1 | 0 | 0.048 | frameshift_variant | - | - | 3.9200000762939453 | - | - | - | - | - | LP | YES, a) |
| Genes related to cardiovascular phenotypes | Familial hypercholesterolemia | APOB | AD | 2:21232140:G:A | rs145143533 | 2078 | 1 | 0 | 0 | 1.0 | 0.0 | 0.0 | 1 | 1 | 0.048 | stop_gained | - | - | 2.3499999046325684 | - | - | 35.0 | 895430 | Conflicting interpretations of p | P | YES, a) |
| Genes related to cardiovascular phenotypes | Familial hypercholesterolemia | APOB | AD | 2:21249778:CC:- | - | 2078 | 1 | 0 | 0 | 1.0 | 0.0 | 0.0 | 1 | 0 | 0.048 | frameshift_variant | - | - | 5.699999809265137 | - | - | - | - | - | LP | YES, a) |
| Genes related to cardiovascular phenotypes | Familial hypercholesterolemia | APOB | AD | 2:21249782:-:CC | - | 2078 | 1 | 0 | 0 | 1.0 | 0.0 | 0.0 | 1 | 0 | 0.048 | frameshift_variant | - | - | 4.550000190734863 | - | - | - | - | - | LP | YES, a) |
| Genes related to cardiovascular phenotypes | Dilated cardiomyopathy | DES | AD | 2:220283700:GC:- | rs769096434 | 2080 | 1 | 0 | 0 | 1.0 | 0.0 | 0.0 | 1 | 1 | 0.048 | frameshift_variant | - | - | 3.6700000762939453 | - | - | - | 1069497 | Pathogenic | P | YES, a) |
| Genes related to cancer phenotypes | Lynch syndrome | MSH2 | AD | 2:47637443:CA:- | - | 2021 | 1 | 0 | 0 | 1.0 | 0.0 | 0.0 | 1 | 0 | 0.049 | frameshift_variant | - | - | 4.590000152587891 | - | - | - | - | - | LP | YES, a) |
| Genes related to cancer phenotypes | Lynch syndrome | MSH2 | AD | 2:47637446:A:- | - | 2021 | 1 | 0 | 0 | 1.0 | 0.0 | 0.0 | 1 | 0 | 0.049 | frameshift_variant | - | - | 3.2899999618530273 | - | - | - | - | - | LP | YES, a) |
| Genes related to cancer phenotypes | Lynch syndrome | MSH6 | AD | 2:48026290:G:- | rs753796271 | 1416 | 1 | 0 | 0 | 1.0 | 0.0 | 0.0 | 1 | 1 | 0.071 | frameshift_variant | - | - | 4.630000114440918 | - | - | - | 428404 | Pathogenic/Likely pathogenic | P/LP | YES, a) |
| Genes related to cancer phenotypes | Lynch syndrome | MSH6 | AD | 2:48030653:-:A | rs1553331434 | 1416 | 1 | 0 | 0 | 1.0 | 0.0 | 0.0 | 1 | 1 | 0.071 | frameshift_variant | - | - | 5.380000114440918 | - | - | - | 372412 | Pathogenic | P | YES, a) |
| Genes related to cancer phenotypes | Hereditary paraganglioma–pheochromocytoma syndrome | TMEM127 | AD | 2:96931034:C:- | - | 2021 | 1 | 0 | 0 | 1.0 | 0.0 | 0.0 | 1 | 0 | 0.049 | frameshift_variant | - | - | 4.71999979019165 | - | - | - | - | - | LP | YES, a) |
| Genes related to cancer phenotypes | Neurofibromatosis type 2 | NF2 | AD | 22:30050682:TTTTT:- | - | 2021 | 1 | 0 | 0 | 1.0 | 0.0 | 0.0 | 1 | 0 | 0.049 | frameshift_variant | - | - | 5.989999771118164 | - | - | - | - | - | LP | YES, a) |
| Genes related to cancer phenotypes | Neurofibromatosis type 2 | NF2 | AD | 22:30057219:TTGGA:- | - | 2021 | 1 | 0 | 0 | 1.0 | 0.0 | 0.0 | 1 | 0 | 0.049 | frameshift_variant | - | - | 0.9210000038146973 | - | - | - | - | - | LP | YES, a) |
| Genes related to cancer phenotypes | Neurofibromatosis type 2 | NF2 | AD | 22:30057224:-:CCAAG | - | 2021 | 1 | 0 | 0 | 1.0 | 0.0 | 0.0 | 1 | 0 | 0.049 | frameshift_variant | - | - | 0.9210000038146973 | - | - | - | - | - | LP | YES, a) |
| Genes related to cardiovascular phenotypes | Arrhythmogenic right ventricular cardiomyopathy | TMEM43 | AD | 3:14174056:G:T | - | 2080 | 1 | 0 | 0 | 1.0 | 0.0 | 0.0 | 1 | 1 | 0.048 | stop_gained | - | - | 4.989999771118164 | - | - | 40.0 | - | - | P | YES, a) |
| Genes related to cardiovascular phenotypes | Arrhythmogenic right ventricular cardiomyopathy | TMEM43 | AD | 3:14180678:A:G | rs756115936 | 915 | 1 | 0 | 0 | 0.999 | 0.001 | 0.001 | 1 | 1 | 0.109 | splice_acceptor_variant | - | - | 5.25 | - | - | 24.0 | - | - | P | YES, a) |
| Genes related to inborn errors of metabolism phenotypes | Biotinidase deficiency | BTD | AR | 3:15676986:GG:- | rs765906887 | 2078 | 1 | 0 | 0 | 1.0 | 0.0 | 0.0 | 1 | 1 | 0.048 | frameshift_variant | - | - | 2.859999895095825 | - | - | - | 373906 | Pathogenic | P | YES, b) * |
| Genes related to inborn errors of metabolism phenotypes | Biotinidase deficiency | BTD | AR | 3:15676989:TG:- | rs750965140 | 2078 | 1 | 0 | 0 | 1.0 | 0.0 | 0.0 | 1 | 1 | 0.048 | frameshift_variant | - | - | 3.700000047683716 | - | - | - | 801942 | Pathogenic | P | YES, b) * |
| Genes related to inborn errors of metabolism phenotypes | Biotinidase deficiency | BTD | AR | 3:15685958:G:A | rs397514375 | 2078 | 1 | 0 | 0 | 1.0 | 0.0 | 0.0 | 1 | 0 | 0.048 | missense_variant | - | - | 5.079999923706055 | 0.0 | 0.984 | 24.700000762939453 | 25026 | Pathogenic/Likely pathogenic | P/LP | NO, b) |
| Genes related to inborn errors of metabolism phenotypes | Biotinidase deficiency | BTD | AR | 3:15686128:C:G | rs1575029719 | 2078 | 1 | 0 | 0 | 1.0 | 0.0 | 0.0 | 1 | 0 | 0.048 | missense_variant | - | - | 0.2680000066757202 | 0.04 | 0.555 | 7.050000190734863 | 801946 | Likely pathogenic | LP | NO, b) |
| Genes related to inborn errors of metabolism phenotypes | Biotinidase deficiency | BTD | AR | 3:15686731:A:C | rs80338685 | 2078 | 1 | 0 | 0 | 1.0 | 0.0 | 0.0 | 1 | 0 | 0.048 | missense_variant | 0.0009940357 | - | -2.259999990463257 | 0.0 | 0.993 | 22.899999618530273 | 1902 | Pathogenic/Likely pathogenic | P/LP | NO, b) |
| Genes related to inborn errors of metabolism phenotypes | Biotinidase deficiency | BTD | AR | 3:15686975:C:T | rs80338686 | 2078 | 1 | 0 | 0 | 1.0 | 0.0 | 0.0 | 1 | 1 | 0.048 | missense_variant | - | - | 4.690000057220459 | 0.0 | 1.0 | 34.0 | 1898 | Pathogenic | P | NO, b) |
| Genes related to cardiovascular phenotypes | Aortopathies | TGFBR2 | AD | 3:30664711:A:- | - | 2053 | 1 | 0 | 0 | 1.0 | 0.0 | 0.0 | 1 | 0 | 0.049 | frameshift_variant | - | - | 2.9000000953674316 | - | - | - | - | - | LP | YES, a) |
| Genes related to cardiovascular phenotypes | Aortopathies | TGFBR2 | AD | 3:30664716:T:- | - | 2053 | 1 | 0 | 0 | 1.0 | 0.0 | 0.0 | 1 | 0 | 0.049 | frameshift_variant | - | - | -2.759999990463257 | - | - | - | - | - | LP | YES, a) |
| Genes related to cardiovascular phenotypes | Aortopathies | TGFBR2 | AD | 3:30664718:-:CA | - | 2053 | 1 | 0 | 0 | 1.0 | 0.0 | 0.0 | 1 | 0 | 0.049 | frameshift_variant | - | - | 2.4100000858306885 | - | - | - | - | - | LP | YES, a) |
| Genes related to cancer phenotypes | Lynch syndrome | MLH1 | AD | 3:37035041:-:G | - | 2021 | 1 | 0 | 0 | 1.0 | 0.0 | 0.0 | 1 | 1 | 0.049 | frameshift_variant | - | - | 5.869999885559082 | - | - | - | - | - | LP | YES, a) |
| Genes related to cardiovascular phenotypes | Hypertrophic cardiomyopathy | MYL3 | AD | 3:46899743:C:- | - | 2080 | 1 | 0 | 0 | 1.0 | 0.0 | 0.0 | 1 | 1 | 0.048 | frameshift_variant | - | - | 4.0 | - | - | - | - | - | LP | YES, a) |
| Genes related to cardiovascular phenotypes | Hypertrophic cardiomyopathy | MYL3 | AD | 3:46902303:G:T | rs139794067 | 1434 | 1 | 0 | 0 | 1.0 | 0.0 | 0.0 | 1 | 1 | 0.070 | missense_variant | - | - | 4.360000133514404 | 0.0 | 0.996 | 29.799999237060547 | 43121 | Conflicting interpretations of p | LP | YES, a) |
| Genes related to cancer phenotypes | Familial adenomatous polyposis | APC | AD | 5:112090668:AGAAG:- | - | 2021 | 1 | 0 | 0 | 1.0 | 0.0 | 0.0 | 1 | 0 | 0.049 | frameshift_variant | - | - | 5.670000076293945 | - | - | - | - | - | LP | YES, a) |
| Genes related to cancer phenotypes | Familial adenomatous polyposis | APC | AD | 5:112090674:-:TCCTC | - | 2021 | 1 | 0 | 0 | 1.0 | 0.0 | 0.0 | 1 | 0 | 0.049 | frameshift_variant | - | - | 5.670000076293945 | - | - | - | - | - | LP | YES, a) |
| Genes related to cancer phenotypes | Familial adenomatous polyposis | APC | AD | 5:112111328:C:A | - | 2021 | 1 | 0 | 0 | 1.0 | 0.0 | 0.0 | 1 | 1 | 0.049 | stop_gained | - | - | 5.449999809265137 | - | - | 36.0 | - | - | P | YES, a) |
| Genes related to cancer phenotypes | Familial adenomatous polyposis | APC | AD | 5:112154723:C:T | rs775126020 | 1416 | 1 | 0 | 0 | 1.0 | 0.0 | 0.0 | 1 | 1 | 0.071 | stop_gained | - | - | 4.840000152587891 | - | - | 37.0 | 184937 | Pathogenic | P | YES, a) |
| Genes related to cardiovascular phenotypes | Catecholaminergic polymorphic ventricular tachycardia | TRDN | AR | 6:123759209:T:- | rs753514580 | 2079 | 1 | 0 | 1 | 1.0 | 0.0 | 0.0 | 1 | 0 | 0.048 | frameshift_variant | - | - | 4.329999923706055 | - | - | - | 1325231 | Likely pathogenic | LP | NO, b) |
| Genes related to cardiovascular phenotypes | Catecholaminergic polymorphic ventricular tachycardia | TRDN | AR | 6:123824979:T:- | - | 2080 | 1 | 0 | 0 | 1.0 | 0.0 | 0.0 | 1 | 1 | 0.048 | frameshift_variant | - | - | 0.6209999918937683 | - | - | - | - | - | LP | NO, b) |
| Genes related to cardiovascular phenotypes | Catecholaminergic polymorphic ventricular tachycardia | TRDN | AR | 6:123868496:-:T | - | 2080 | 1 | 0 | 0 | 1.0 | 0.0 | 0.0 | 1 | 0 | 0.048 | frameshift_variant | - | - | 1.940000057220459 | - | - | - | - | - | LP | NO, b) |
| Genes related to cardiovascular phenotypes | Arrhythmogenic right ventricular cardiomyopathy | DSP | AD | 6:7555992:-:CT | - | 2080 | 1 | 0 | 0 | 1.0 | 0.0 | 0.0 | 1 | 0 | 0.048 | frameshift_variant | - | - | 4.570000171661377 | - | - | - | - | - | LP | YES, a) |
| Genes related to cardiovascular phenotypes | Arrhythmogenic right ventricular cardiomyopathy | DSP | AD | 6:7556002:-:C | - | 2080 | 1 | 0 | 0 | 1.0 | 0.0 | 0.0 | 1 | 0 | 0.048 | frameshift_variant | - | - | 3.2200000286102295 | - | - | - | - | - | LP | YES, a) |
| Genes related to cardiovascular phenotypes | Arrhythmogenic right ventricular cardiomyopathy | DSP | AD | 6:7578019:A:- | - | 2080 | 1 | 0 | 0 | 1.0 | 0.0 | 0.0 | 1 | 0 | 0.048 | frameshift_variant | - | - | 5.550000190734863 | - | - | - | - | - | LP | YES, a) |
| Genes related to cardiovascular phenotypes | Arrhythmogenic right ventricular cardiomyopathy | DSP | AD | 6:7578024:-:T | - | 2080 | 1 | 0 | 0 | 1.0 | 0.0 | 0.0 | 1 | 0 | 0.048 | frameshift_variant | - | - | -5.340000152587891 | - | - | - | - | - | LP | YES, a) |
| Genes related to cardiovascular phenotypes | Dilated cardiomyopathy | FLNC | AD | 7:128490537:G:T | - | 2068 | 1 | 0 | 0 | 1.0 | 0.0 | 0.0 | 1 | 1 | 0.048 | stop_gained | - | - | 5.409999847412109 | - | - | 47.0 | - | - | P | YES, a) |
| Genes related to cardiovascular phenotypes | Long QT syndrome types 1 and 2 | KCNH2 | AD | 7:150656679:-:G | rs761863251 | 2068 | 1 | 0 | 0 | 1.0 | 0.0 | 0.0 | 1 | 1 | 0.048 | frameshift_variant | - | - | 2.0799999237060547 | - | - | - | 503669 | Conflicting interpretations of p | LP | YES, a) |
| Genes related to cardiovascular phenotypes | Hypertrophic cardiomyopathy | PRKAG2 | AD | 7:151262466:-:GGTC | - | 2080 | 1 | 0 | 0 | 1.0 | 0.0 | 0.0 | 1 | 0 | 0.048 | frameshift_variant | - | - | 5.429999828338623 | - | - | - | - | - | LP | YES, a) |
| Genes related to miscellaneous phenotypes | Hereditary hemorrhagic telangiectasia | ENG | AD | 9:130605480:CGGGG:- | - | 2080 | 1 | 0 | 0 | 1.0 | 0.0 | 0.0 | 1 | 0 | 0.048 | frameshift_variant | - | - | 4.230000019073486 | - | - | - | - | - | LP | YES, a) |
| Genes related to miscellaneous phenotypes | Hereditary hemorrhagic telangiectasia | ENG | AD | 9:130605485:-:TAAAA | - | 2080 | 1 | 0 | 0 | 1.0 | 0.0 | 0.0 | 1 | 0 | 0.048 | frameshift_variant | - | - | 4.230000019073486 | - | - | - | - | - | LP | YES, a) |
| Genes related to cancer phenotypes | Tuberous sclerosis complex | TSC1 | AD | 9:135772877:G:- | - | 2021 | 1 | 0 | 0 | 1.0 | 0.0 | 0.0 | 1 | 0 | 0.049 | frameshift_variant | - | - | -0.6129999756813049 | - | - | - | - | - | LP | YES, a) |
| Genes related to cancer phenotypes | Tuberous sclerosis complex | TSC1 | AD | 9:135772880:GA:- | - | 2021 | 1 | 0 | 0 | 1.0 | 0.0 | 0.0 | 1 | 0 | 0.049 | frameshift_variant | - | - | 4.449999809265137 | - | - | - | - | - | LP | YES, a) |
| Genes related to cancer phenotypes | Tuberous sclerosis complex | TSC1 | AD | 9:135772998:C:A | - | 890 | 1 | 0 | 1 | 0.999 | 0.001 | 0.001 | 1 | 1 | 0.112 | splice_acceptor_variant | - | - | 5.789999961853027 | - | - | 25.399999618530273 | - | - | P | YES, a) |
| Genes related to cancer phenotypes | Tuberous sclerosis complex | TSC1 | AD | 9:135772999:T:A | rs118203717 | 882 | 1 | 0 | 4 | 0.999 | 0.001 | 0.001 | 1 | 1 | 0.113 | splice_acceptor_variant | - | - | 4.639999866485596 | - | - | 23.200000762939453 | - | - | P | YES, a) |
| Genes related to cancer phenotypes | Tuberous sclerosis complex | TSC1 | AD | 9:135781092:CA:- | - | 2021 | 1 | 0 | 0 | 1.0 | 0.0 | 0.0 | 1 | 0 | 0.049 | frameshift_variant | - | - | -10.699999809265137 | - | - | - | - | - | LP | YES, a) |
| Genes related to cancer phenotypes | Tuberous sclerosis complex | TSC1 | AD | 9:135781098:-:C | - | 2021 | 1 | 0 | 0 | 1.0 | 0.0 | 0.0 | 1 | 0 | 0.049 | frameshift_variant | - | - | 5.940000057220459 | - | - | - | - | - | LP | YES, a) |
| Genes related to cancer phenotypes | Tuberous sclerosis complex | TSC1 | AD | 9:135781104:-:G | - | 2021 | 1 | 0 | 0 | 1.0 | 0.0 | 0.0 | 1 | 0 | 0.049 | frameshift_variant | - | - | 5.039999961853027 | - | - | - | - | - | LP | YES, a) |
| Genes related to inborn errors of metabolism phenotypes | Fabry disease | GLA | XL | X:100656740:C:T | rs104894845 | 1192 | 0 | 1 | 0 | 0.999 | 0.001 | 0.001 | 1 | 1 | 0.084 | missense_variant | - | - | 5.090000152587891 | 0.02 | 0.647 | 29.899999618530273 | 10748 | Conflicting interpretations of p | LP | YES, c) |
| Genes related to miscellaneous phenotypes | Malignant hyperthermia | CACNA1S | AD | 1:201060844:CATA:- | - | 2094 | 2 | 0 | 0 | 1.0 | 0.0 | 0.0 | 2 | 0 | 0.095 | frameshift_variant | - | - | -0.5609999895095825 | - | - | - | - | - | LP | YES, a) |
| Genes related to cardiovascular phenotypes | Catecholaminergic polymorphic ventricular tachycardia | RYR2 | AD | 1:237670019:C:- | - | 2079 | 2 | 0 | 0 | 1.0 | 0.0 | 0.0 | 2 | 0 | 0.096 | frameshift_variant | - | - | 4.670000076293945 | - | - | - | - | - | LP | YES, a) |
| Genes related to cardiovascular phenotypes | Catecholaminergic polymorphic ventricular tachycardia | RYR2 | AD | 1:237756778:C:- | - | 2079 | 2 | 0 | 0 | 1.0 | 0.0 | 0.0 | 2 | 0 | 0.096 | frameshift_variant | - | - | 1.0399999618530273 | - | - | - | - | - | LP | YES, a) |
| Genes related to cardiovascular phenotypes | Catecholaminergic polymorphic ventricular tachycardia | RYR2 | AD | 1:237923143:A:- | - | 2079 | 2 | 0 | 0 | 1.0 | 0.0 | 0.0 | 2 | 0 | 0.096 | frameshift_variant | - | - | 5.789999961853027 | - | - | - | - | - | LP | YES, a) |
| Genes related to cancer phenotypes | MUTYH-associated polyposis | MUTYH | AR | 1:45797187:-:CC | rs587780078 | 2020 | 2 | 0 | 0 | 1.0 | 0.0 | 0.0 | 2 | 1 | 0.099 | frameshift_variant | - | - | 2.890000104904175 | - | - | - | 127831 | Pathogenic/Likely pathogenic | P/LP | NO, b) |
| Genes related to miscellaneous phenotypes | RPE65-related retinopathy | RPE65 | AR | 1:68910522:CTTT:- | - | 1977 | 2 | 0 | 1 | 0.999 | 0.001 | 0.001 | 2 | 0 | 0.101 | frameshift_variant | - | - | 4.880000114440918 | - | - | - | - | - | LP | NO, b) |
| Genes related to miscellaneous phenotypes | RPE65-related retinopathy | RPE65 | AR | 1:68910527:-:CCCC | - | 1977 | 2 | 0 | 1 | 0.999 | 0.001 | 0.001 | 2 | 0 | 0.101 | frameshift_variant | - | - | 4.880000114440918 | - | - | - | - | - | LP | NO, b) |
| Genes related to cancer phenotypes | Familial medullary thyroid cancer | RET | AD | 10:43610093:CCTTC:- | - | 2020 | 2 | 0 | 0 | 1.0 | 0.0 | 0.0 | 2 | 0 | 0.099 | frameshift_variant | - | - | 3.8399999141693115 | - | - | - | - | - | LP | YES, a) |
| Genes related to cancer phenotypes | Familial medullary thyroid cancer | RET | AD | 10:43610099:-:AGGAA | - | 2020 | 2 | 0 | 0 | 1.0 | 0.0 | 0.0 | 2 | 0 | 0.099 | frameshift_variant | - | - | 4.75 | - | - | - | - | - | LP | YES, a) |
| Genes related to cancer phenotypes | Familial medullary thyroid cancer | RET | AD | 10:43614996:G:A | rs79658334 | 2008 | 2 | 0 | 0 | 1.0 | 0.0 | 0.0 | 2 | 0 | 0.100 | missense_variant | - | - | 5.360000133514404 | 0.03 | 1.0 | 26.700000762939453 | 37102 | Pathogenic/Likely pathogenic | P/LP | YES, a) |
| Genes related to cardiovascular phenotypes | Hypertrophic cardiomyopathy | MYBPC3 | AD | 11:47354178:T:- | - | 2079 | 2 | 0 | 0 | 1.0 | 0.0 | 0.0 | 2 | 0 | 0.096 | frameshift_variant | - | - | 5.099999904632568 | - | - | - | - | - | LP | YES, a) |
| Genes related to cardiovascular phenotypes | Hypertrophic cardiomyopathy | MYBPC3 | AD | 11:47354183:-:A | - | 2079 | 2 | 0 | 0 | 1.0 | 0.0 | 0.0 | 2 | 0 | 0.096 | frameshift_variant | - | - | 1.2899999618530273 | - | - | - | - | - | LP | YES, a) |
| Genes related to cardiovascular phenotypes | Hypertrophic cardiomyopathy | MYL2 | AD | 12:111352011:-:C | - | 2079 | 2 | 0 | 0 | 1.0 | 0.0 | 0.0 | 2 | 0 | 0.096 | frameshift_variant | - | - | 4.880000114440918 | - | - | - | - | - | LP | YES, a) |
| Genes related to miscellaneous phenotypes | Maturity-onset diabetes of the young | HNF1A | AD | 12:121432117:-:C | rs766191969 | 2077 | 2 | 0 | 0 | 1.0 | 0.0 | 0.0 | 2 | 0 | 0.096 | frameshift_variant | - | - | 4.670000076293945 | - | - | - | 1328238 | Pathogenic | P | YES, a) |
| Genes related to cancer phenotypes | Hereditary breast and/or ovarian cancer | BRCA2 | AD | 13:32905080:CATG:- | - | 2020 | 2 | 0 | 0 | 1.0 | 0.0 | 0.0 | 2 | 0 | 0.099 | frameshift_variant | - | - | 2.130000114440918 | - | - | - | - | - | LP | YES, a) |
| Genes related to cancer phenotypes | Hereditary breast and/or ovarian cancer | BRCA2 | AD | 13:32905085:-:CGTC | - | 2020 | 2 | 0 | 0 | 1.0 | 0.0 | 0.0 | 2 | 0 | 0.099 | frameshift_variant | - | - | 3.390000104904175 | - | - | - | - | - | LP | YES, a) |
| Genes related to cancer phenotypes | Hereditary breast and/or ovarian cancer | BRCA2 | AD | 13:32911093:TACT:- | - | 2020 | 2 | 0 | 0 | 1.0 | 0.0 | 0.0 | 2 | 0 | 0.099 | frameshift_variant | - | - | 2.75 | - | - | - | - | - | LP | YES, a) |
| Genes related to cancer phenotypes | Hereditary breast and/or ovarian cancer | BRCA2 | AD | 13:32911100:ATTT:- | - | 2020 | 2 | 0 | 0 | 1.0 | 0.0 | 0.0 | 2 | 0 | 0.099 | frameshift_variant | - | - | 3.0399999618530273 | - | - | - | - | - | LP | YES, a) |
| Genes related to cancer phenotypes | Hereditary breast and/or ovarian cancer | BRCA2 | AD | 13:32911104:-:GGAGG( | - | 2020 | 2 | 0 | 0 | 1.0 | 0.0 | 0.0 | 2 | 0 | 0.099 | frameshift_variant | - | - | 3.0399999618530273 | - | - | - | - | - | LP | YES, a) |
| Genes related to cancer phenotypes | Hereditary breast and/or ovarian cancer | BRCA2 | AD | 13:32911772:AA:- | - | 2020 | 2 | 0 | 0 | 1.0 | 0.0 | 0.0 | 2 | 0 | 0.099 | frameshift_variant | - | - | 3.240000009536743 | - | - | - | - | - | LP | YES, a) |
| Genes related to cancer phenotypes | Hereditary breast and/or ovarian cancer | BRCA2 | AD | 13:32911797:-:T | rs1555283160 | 2020 | 2 | 0 | 0 | 1.0 | 0.0 | 0.0 | 2 | 0 | 0.099 | frameshift_variant | - | - | 4.5 | - | - | - | 959854 | Pathogenic | P | YES, a) |
| Genes related to cancer phenotypes | Hereditary breast and/or ovarian cancer | BRCA2 | AD | 13:32912471:GC:- | - | 2020 | 2 | 0 | 0 | 1.0 | 0.0 | 0.0 | 2 | 0 | 0.099 | frameshift_variant | - | - | 1.850000023841858 | - | - | - | - | - | LP | YES, a) |
| Genes related to cancer phenotypes | Hereditary breast and/or ovarian cancer | BRCA2 | AD | 13:32912476:-:TC | - | 2020 | 2 | 0 | 0 | 1.0 | 0.0 | 0.0 | 2 | 0 | 0.099 | frameshift_variant | - | - | -3.0 | - | - | - | - | - | LP | YES, a) |
| Genes related to cancer phenotypes | Hereditary breast and/or ovarian cancer | BRCA2 | AD | 13:32913528:CTTCT:- | - | 2020 | 2 | 0 | 0 | 1.0 | 0.0 | 0.0 | 2 | 0 | 0.099 | frameshift_variant | - | - | 5.760000228881836 | - | - | - | - | - | LP | YES, a) |
| Genes related to cancer phenotypes | Hereditary breast and/or ovarian cancer | BRCA2 | AD | 13:32913533:-:AGGGA | - | 2020 | 2 | 0 | 0 | 1.0 | 0.0 | 0.0 | 2 | 0 | 0.099 | frameshift_variant | - | - | 5.760000228881836 | - | - | - | - | - | LP | YES, a) |
| Genes related to cancer phenotypes | Hereditary breast and/or ovarian cancer | BRCA2 | AD | 13:32915296:-:GA | - | 2020 | 2 | 0 | 0 | 1.0 | 0.0 | 0.0 | 2 | 0 | 0.099 | frameshift_variant | - | - | 5.710000038146973 | - | - | - | - | - | LP | YES, a) |
| Genes related to miscellaneous phenotypes | Wilson disease | ATP7B | AR | 13:52516532:G:- | rs137853281 | 2077 | 2 | 0 | 0 | 1.0 | 0.0 | 0.0 | 2 | 0 | 0.096 | frameshift_variant | - | - | -2.0199999809265137 | - | - | - | 88958 | Pathogenic | P | NO, b) |
| Genes related to cardiovascular phenotypes | Aortopathies | SMAD3 | AD | 15:67358548:AGAAG:- | - | 2079 | 2 | 0 | 0 | 1.0 | 0.0 | 0.0 | 2 | 0 | 0.096 | frameshift_variant | - | - | 0.382999986410141 | - | - | - | - | - | LP | YES, a) |
| Genes related to cardiovascular phenotypes | Aortopathies | SMAD3 | AD | 15:67358553:-:TCTTC | - | 2079 | 2 | 0 | 0 | 1.0 | 0.0 | 0.0 | 2 | 0 | 0.096 | frameshift_variant | - | - | 0.382999986410141 | - | - | - | - | - | LP | YES, a) |
| Genes related to cancer phenotypes | Tuberous sclerosis complex | TSC2 | AD | 16:2098688:GAGG:- | - | 2020 | 2 | 0 | 0 | 1.0 | 0.0 | 0.0 | 2 | 0 | 0.099 | frameshift_variant | - | - | -0.16699999570846558 | - | - | - | - | - | LP | YES, a) |
| Genes related to cancer phenotypes | Tuberous sclerosis complex | TSC2 | AD | 16:2098693:-:TCCG | - | 2020 | 2 | 0 | 0 | 1.0 | 0.0 | 0.0 | 2 | 0 | 0.099 | frameshift_variant | - | - | 4.199999809265137 | - | - | - | - | - | LP | YES, a) |
| Genes related to cancer phenotypes | Hereditary breast and/or ovarian cancer | BRCA1 | AD | 17:41245426:AACCA:- | - | 2020 | 2 | 0 | 0 | 1.0 | 0.0 | 0.0 | 2 | 0 | 0.099 | frameshift_variant | - | - | 2.950000047683716 | - | - | - | - | - | LP | YES, a) |
| Genes related to cancer phenotypes | Hereditary breast and/or ovarian cancer | BRCA1 | AD | 17:41246064:-:TA | - | 2020 | 2 | 0 | 0 | 1.0 | 0.0 | 0.0 | 2 | 0 | 0.099 | frameshift_variant | - | - | 1.1399999856948853 | - | - | - | - | - | LP | YES, a) |
| Genes related to cancer phenotypes | Hereditary breast and/or ovarian cancer | BRCA1 | AD | 17:41246066:-:T | - | 2020 | 2 | 0 | 0 | 1.0 | 0.0 | 0.0 | 2 | 0 | 0.099 | frameshift_variant | - | - | 3.049999952316284 | - | - | - | - | - | LP | YES, a) |
| Genes related to cancer phenotypes | Hereditary breast and/or ovarian cancer | BRCA1 | AD | 17:41246071:T:- | rs786203982 | 2020 | 2 | 0 | 0 | 1.0 | 0.0 | 0.0 | 2 | 0 | 0.099 | frameshift_variant | - | - | -1.5800000429153442 | - | - | - | 187763 | Pathogenic | P | YES, a) |
| Genes related to cancer phenotypes | Hereditary breast and/or ovarian cancer | BRCA1 | AD | 17:41246075:-:A | - | 2020 | 2 | 0 | 0 | 1.0 | 0.0 | 0.0 | 2 | 0 | 0.099 | frameshift_variant | - | - | 0.8450000286102295 | - | - | - | - | - | LP | YES, a) |
| Genes related to inborn errors of metabolism phenotypes | Pompe disease | GAA | AR | 17:78079569:C:T | rs771258854 | 2077 | 2 | 0 | 0 | 1.0 | 0.0 | 0.0 | 2 | 1 | 0.096 | missense_variant | - | - | 5.050000190734863 | 0.0 | 0.999 | 31.0 | 645336 | Pathogenic | P | NO, b) |
| Genes related to inborn errors of metabolism phenotypes | Pompe disease | GAA | AR | 17:78086420:C:T | rs764670084 | 2077 | 2 | 0 | 0 | 1.0 | 0.0 | 0.0 | 2 | 1 | 0.096 | missense_variant | - | - | 2.630000114440918 | 0.0 | 1.0 | 33.0 | 640911 | Pathogenic | P | NO, b) |

| | | | | | | | | | | | | | | | | | | | | | | | | | | |
|---|---|---|---|---|---|---|---|---|---|---|---|---|---|---|---|---|---|---|---|---|---|---|---|---|---|---|
| Genes related to miscellaneous phenotypes | Malignant hyperthermia | RYR1 | AD | 19:38934252:C:T | rs118192173 | 2048 | 2 | 0 | 0 | 1.0 | 0.0 | 0.0 | 2 | 1 | 0.098 | missense_variant | - | - | 4.079999923706055 | 0.0 | 0.994 | 34.0 | 12988 | Pathogenic/Likely pathogenic | P/LP | YES, a) |
| Genes related to miscellaneous phenotypes | Malignant hyperthermia | RYR1 | AD | 19:38948185:C:T | rs118192172 | 2094 | 2 | 0 | 0 | 1.0 | 0.0 | 0.0 | 2 | 1 | 0.095 | missense_variant | 0.0009940357 | 0.004672897 | 3.759999990463257 | 0.0 | 1.0 | 35.0 | 12964 | Pathogenic; drug response | P | YES, a) |
| Genes related to miscellaneous phenotypes | Malignant hyperthermia | RYR1 | AD | 19:39077179:ATTTG:- | - | 2094 | 2 | 0 | 0 | 1.0 | 0.0 | 0.0 | 2 | 0 | 0.095 | frameshift_variant | - | - | 3.5799999237060547 | - | - | - | - | - | LP | YES, a) |
| Genes related to miscellaneous phenotypes | Malignant hyperthermia | RYR1 | AD | 19:39077185:-:CCCAG | - | 2094 | 2 | 0 | 0 | 1.0 | 0.0 | 0.0 | 2 | 0 | 0.095 | frameshift_variant | - | - | 4.610000133514404 | - | - | - | - | - | LP | YES, a) |
| Genes related to cardiovascular phenotypes | Dilated cardiomyopathy | TTN | AD | 2:179464123:T:- | - | 2079 | 2 | 0 | 0 | 1.0 | 0.0 | 0.0 | 2 | 0 | 0.096 | frameshift_variant | - | - | -0.052799999713897705 | - | - | - | - | - | LP | YES, a) |
| Genes related to cardiovascular phenotypes | Dilated cardiomyopathy | TTN | AD | 2:179471792:-:A | - | 2061 | 2 | 0 | 6 | 1.0 | 0.0 | 0.0 | 2 | 0 | 0.097 | frameshift_variant | - | - | 1.440000057220459 | - | - | - | - | - | LP | YES, a) |
| Genes related to cardiovascular phenotypes | Dilated cardiomyopathy | TTN | AD | 2:179471813:-:C | - | 2079 | 2 | 0 | 0 | 1.0 | 0.0 | 0.0 | 2 | 0 | 0.096 | frameshift_variant | - | - | 3.440000057220459 | - | - | - | - | - | LP | YES, a) |
| Genes related to cardiovascular phenotypes | Dilated cardiomyopathy | TTN | AD | 2:179477264:A:- | - | 2079 | 2 | 0 | 0 | 1.0 | 0.0 | 0.0 | 2 | 0 | 0.096 | frameshift_variant | - | - | 5.710000038146973 | - | - | - | - | - | LP | YES, a) |
| Genes related to cardiovascular phenotypes | Dilated cardiomyopathy | TTN | AD | 2:179495023:-:AA | - | 2079 | 2 | 0 | 0 | 1.0 | 0.0 | 0.0 | 2 | 0 | 0.096 | frameshift_variant | - | - | 6.039999961853027 | - | - | - | - | - | LP | YES, a) |
| Genes related to cardiovascular phenotypes | Dilated cardiomyopathy | TTN | AD | 2:179495025:-:A | - | 2079 | 2 | 0 | 0 | 1.0 | 0.0 | 0.0 | 2 | 0 | 0.096 | frameshift_variant | - | - | 6.039999961853027 | - | - | - | - | - | LP | YES, a) |
| Genes related to cardiovascular phenotypes | Dilated cardiomyopathy | TTN | AD | 2:179588002:-:CC | - | 2079 | 2 | 0 | 0 | 1.0 | 0.0 | 0.0 | 2 | 0 | 0.096 | frameshift_variant | - | - | 3.3499999046325684 | - | - | - | - | - | LP | YES, a) |
| Genes related to cardiovascular phenotypes | Dilated cardiomyopathy | TTN | AD | 2:179594589:CTGTT:- | - | 2079 | 2 | 0 | 0 | 1.0 | 0.0 | 0.0 | 2 | 0 | 0.096 | frameshift_variant | - | - | 0.44999998807907104 | - | - | - | - | - | LP | YES, a) |
| Genes related to cardiovascular phenotypes | Dilated cardiomyopathy | TTN | AD | 2:179594594:-:GACAA | - | 2079 | 2 | 0 | 0 | 1.0 | 0.0 | 0.0 | 2 | 0 | 0.096 | frameshift_variant | - | - | 0.44999998807907104 | - | - | - | - | - | LP | YES, a) |
| Genes related to cardiovascular phenotypes | Dilated cardiomyopathy | TTN | AD | 2:179613032:AAAT:- | - | 2079 | 2 | 0 | 0 | 1.0 | 0.0 | 0.0 | 2 | 0 | 0.096 | frameshift_variant | - | - | -2.109999895095825 | - | - | - | - | - | LP | YES, a) |
| Genes related to cardiovascular phenotypes | Dilated cardiomyopathy | TTN | AD | 2:179613037:-:GGCA | - | 2079 | 2 | 0 | 0 | 1.0 | 0.0 | 0.0 | 2 | 0 | 0.096 | frameshift_variant | - | - | 1.25 | - | - | - | - | - | LP | YES, a) |
| Genes related to cardiovascular phenotypes | Familial hypercholesterolemia | APOB | AD | 2:21236032:C:A | - | 2013 | 2 | 0 | 6 | 1.0 | 0.0 | 0.0 | 2 | 0 | 0.099 | stop_gained | - | - | 5.210000038146973 | - | - | 39.0 | - | - | P | YES, a) |
| Genes related to cancer phenotypes | Lynch syndrome | MSH2 | AD | 2:47672724:-:TT | - | 2020 | 2 | 0 | 0 | 1.0 | 0.0 | 0.0 | 2 | 0 | 0.099 | frameshift_variant | - | - | 5.559999942779541 | - | - | - | - | - | LP | YES, a) |
| Genes related to cancer phenotypes | Neurofibromatosis type 2 | NF2 | AD | 22:30057234:TGGGG:- | - | 2020 | 2 | 0 | 0 | 1.0 | 0.0 | 0.0 | 2 | 0 | 0.099 | frameshift_variant | - | - | -2.930000066757202 | - | - | - | - | - | LP | YES, a) |
| Genes related to cancer phenotypes | Neurofibromatosis type 2 | NF2 | AD | 22:30057239:-:CAAAA | - | 2020 | 2 | 0 | 0 | 1.0 | 0.0 | 0.0 | 2 | 0 | 0.099 | frameshift_variant | - | - | -2.930000066757202 | - | - | - | - | - | LP | YES, a) |
| Genes related to cancer phenotypes | Neurofibromatosis type 2 | NF2 | AD | 22:30070859:CA:- | - | 2020 | 2 | 0 | 0 | 1.0 | 0.0 | 0.0 | 2 | 0 | 0.099 | frameshift_variant | - | - | 5.599999904632568 | - | - | - | - | - | LP | YES, a) |
| Genes related to cancer phenotypes | Neurofibromatosis type 2 | NF2 | AD | 22:30070864:-:AG | - | 2020 | 2 | 0 | 0 | 1.0 | 0.0 | 0.0 | 2 | 0 | 0.099 | frameshift_variant | - | - | 5.599999904632568 | - | - | - | - | - | LP | YES, a) |
| Genes related to inborn errors of metabolism phenotypes | Biotinidase deficiency | BTD | AR | 3:15683446:G:T | rs375712490 | 2077 | 2 | 0 | 0 | 1.0 | 0.0 | 0.0 | 2 | 2 | 0.096 | missense_variant | - | - | 5.820000171661377 | 0.0 | 0.999 | 31.0 | 24999 | Pathogenic/Likely pathogenic | P/LP | NO, b) |
| Genes related to inborn errors of metabolism phenotypes | Biotinidase deficiency | BTD | AR | 3:15685994:C:T | rs372844636 | 2077 | 2 | 0 | 0 | 1.0 | 0.0 | 0.0 | 2 | 0 | 0.096 | missense_variant | - | - | 5.619999885559082 | 0.0 | 0.989 | 34.0 | 25028 | Pathogenic | P | NO, b) |
| Genes related to inborn errors of metabolism phenotypes | Biotinidase deficiency | BTD | AR | 3:15686296:T:- | rs397514395 | 2077 | 2 | 0 | 0 | 1.0 | 0.0 | 0.0 | 2 | 1 | 0.096 | frameshift_variant | - | - | -1.940000057220459 | - | - | - | 25052 | Pathogenic/Likely pathogenic | P/LP | NO, b) |
| Genes related to cardiovascular phenotypes | Long QT syndrome 3; Brugada syndrome | SCN5A | AD | 3:38595909:G:- | - | 2079 | 2 | 0 | 0 | 1.0 | 0.0 | 0.0 | 2 | 2 | 0.096 | frameshift_variant | - | - | 2.7799999713897705 | - | - | - | - | - | LP | YES, a) |
| Genes related to cardiovascular phenotypes | Catecholaminergic polymorphic ventricular tachycardia | TRDN | AR | 6:123687285:TTAA:- | - | 2079 | 2 | 0 | 0 | 1.0 | 0.0 | 0.0 | 2 | 0 | 0.096 | frameshift_variant | - | - | 4.679999828338623 | - | - | - | - | - | LP | NO, b) |
| Genes related to cardiovascular phenotypes | Catecholaminergic polymorphic ventricular tachycardia | TRDN | AR | 6:123687292:-:TCCA | - | 2079 | 2 | 0 | 0 | 1.0 | 0.0 | 0.0 | 2 | 0 | 0.096 | frameshift_variant | - | - | 5.550000190734863 | - | - | - | - | - | LP | NO, b) |
| Genes related to cardiovascular phenotypes | Catecholaminergic polymorphic ventricular tachycardia | TRDN | AR | 6:123818381:ACAGA:- | - | 2079 | 2 | 0 | 0 | 1.0 | 0.0 | 0.0 | 2 | 0 | 0.096 | frameshift_variant | - | - | 6.03000020980835 | - | - | - | - | - | LP | NO, b) |
| Genes related to cardiovascular phenotypes | Catecholaminergic polymorphic ventricular tachycardia | TRDN | AR | 6:123818387:-:GTCTT | - | 2079 | 2 | 0 | 0 | 1.0 | 0.0 | 0.0 | 2 | 0 | 0.096 | frameshift_variant | - | - | 6.03000020980835 | - | - | - | - | - | LP | NO, b) |
| Genes related to cardiovascular phenotypes | Catecholaminergic polymorphic ventricular tachycardia | TRDN | AR | 6:123868486:T:- | rs757805966 | 2079 | 2 | 0 | 0 | 1.0 | 0.0 | 0.0 | 2 | 0 | 0.096 | frameshift_variant | - | - | 4.380000114440918 | - | - | - | 1074440 | Pathogenic | P | NO, b) |
| Genes related to cardiovascular phenotypes | Arrhythmogenic right ventricular cardiomyopathy | DSP | AD | 6:7577247:T:- | - | 2079 | 2 | 0 | 0 | 1.0 | 0.0 | 0.0 | 2 | 0 | 0.096 | frameshift_variant | - | - | 3.3399999141693115 | - | - | - | - | - | LP | YES, a) |
| Genes related to cardiovascular phenotypes | Arrhythmogenic right ventricular cardiomyopathy | DSP | AD | 6:7577251:T:- | - | 2079 | 2 | 0 | 0 | 1.0 | 0.0 | 0.0 | 2 | 0 | 0.096 | frameshift_variant | - | - | -3.3299999237060547 | - | - | - | - | - | LP | YES, a) |
| Genes related to cardiovascular phenotypes | Arrhythmogenic right ventricular cardiomyopathy | DSP | AD | 6:7577253:-:AC | - | 2079 | 2 | 0 | 0 | 1.0 | 0.0 | 0.0 | 2 | 0 | 0.096 | frameshift_variant | - | - | 5.769999980926514 | - | - | - | - | - | LP | YES, a) |
| Genes related to cardiovascular phenotypes | Arrhythmogenic right ventricular cardiomyopathy | DSP | AD | 6:7580369:-:A | - | 2079 | 2 | 0 | 0 | 1.0 | 0.0 | 0.0 | 2 | 0 | 0.096 | frameshift_variant | - | - | 4.400000095367432 | - | - | - | - | - | LP | YES, a) |
| Genes related to cardiovascular phenotypes | Arrhythmogenic right ventricular cardiomyopathy | DSP | AD | 6:7581780:AATT:- | - | 2079 | 2 | 0 | 0 | 1.0 | 0.0 | 0.0 | 2 | 0 | 0.096 | frameshift_variant | - | - | 6.019999980926514 | - | - | - | - | - | LP | YES, a) |
| Genes related to cardiovascular phenotypes | Arrhythmogenic right ventricular cardiomyopathy | DSP | AD | 6:7581784:-:GGCC | - | 2079 | 2 | 0 | 0 | 1.0 | 0.0 | 0.0 | 2 | 0 | 0.096 | frameshift_variant | - | - | 6.019999980926514 | - | - | - | - | - | LP | YES, a) |
| Genes related to cardiovascular phenotypes | Dilated cardiomyopathy | FLNC | AD | 7:128482372:A:T | - | 2079 | 2 | 0 | 0 | 1.0 | 0.0 | 0.0 | 2 | 0 | 0.096 | stop_gained | - | - | 5.539999961853027 | - | - | 41.0 | - | - | P | YES, a) |
| Genes related to cardiovascular phenotypes | Dilated cardiomyopathy | FLNC | AD | 7:128482880:-:AATAC | - | 2079 | 2 | 0 | 0 | 1.0 | 0.0 | 0.0 | 2 | 0 | 0.096 | frameshift_variant | - | - | -11.399999618530273 | - | - | - | - | - | LP | YES, a) |
| Genes related to cardiovascular phenotypes | Dilated cardiomyopathy | FLNC | AD | 7:128482881:GC:- | - | 2079 | 2 | 0 | 0 | 1.0 | 0.0 | 0.0 | 2 | 0 | 0.096 | frameshift_variant | - | - | -3.140000104904175 | - | - | - | - | - | LP | YES, a) |
| Genes related to cardiovascular phenotypes | Long QT syndrome types 1 and 2 | KCNH2 | AD | 7:150644579:TGTTG:- | - | 2079 | 2 | 0 | 0 | 1.0 | 0.0 | 0.0 | 2 | 0 | 0.096 | frameshift_variant | - | - | 4.980000019073486 | - | - | - | - | - | LP | YES, a) |
| Genes related to cardiovascular phenotypes | Long QT syndrome types 1 and 2 | KCNH2 | AD | 7:150644584:-:CACCA | - | 2079 | 2 | 0 | 0 | 1.0 | 0.0 | 0.0 | 2 | 0 | 0.096 | frameshift_variant | - | - | 4.980000019073486 | - | - | - | - | - | LP | YES, a) |
| Genes related to cardiovascular phenotypes | Long QT syndrome types 1 and 2 | KCNH2 | AD | 7:150648737:G:A | rs121912508 | 2079 | 2 | 0 | 0 | 1.0 | 0.0 | 0.0 | 2 | 0 | 0.096 | missense_variant | - | - | 3.490000009536743 | 0.0 | 0.993 | 26.0 | 14428 | Pathogenic | P | YES, a) |
| Genes related to cancer phenotypes | Tuberous sclerosis complex | TSC1 | AD | 9:135772912:-:A | - | 2020 | 2 | 0 | 0 | 1.0 | 0.0 | 0.0 | 2 | 0 | 0.099 | frameshift_variant | - | - | 1.090000033378601 | - | - | - | - | - | LP | YES, a) |
| Genes related to cancer phenotypes | Tuberous sclerosis complex | TSC1 | AD | 9:135797360:C:- | - | 1999 | 2 | 0 | 9 | 1.0 | 0.0 | 0.0 | 2 | 0 | 0.100 | frameshift_variant | - | - | 6.079999923706055 | - | - | - | - | - | LP | YES, a) |
| Genes related to miscellaneous phenotypes | Malignant hyperthermia | CACNA1S | AD | 1:201060849:-:CGCC | - | 2093 | 3 | 0 | 0 | 0.999 | 0.001 | 0.001 | 3 | 0 | 0.143 | frameshift_variant | - | - | 5.039999961853027 | - | - | - | - | - | LP | YES, a) |
| Genes related to cardiovascular phenotypes | Catecholaminergic polymorphic ventricular tachycardia | RYR2 | AD | 1:237948243:CCCA:- | - | 2078 | 3 | 0 | 0 | 0.999 | 0.001 | 0.001 | 3 | 0 | 0.144 | frameshift_variant | - | - | -7.349999904632568 | - | - | - | - | - | LP | YES, a) |
| Genes related to cardiovascular phenotypes | Catecholaminergic polymorphic ventricular tachycardia | RYR2 | AD | 1:237948248:-:TTGC | - | 2078 | 3 | 0 | 0 | 0.999 | 0.001 | 0.001 | 3 | 0 | 0.144 | frameshift_variant | - | - | 4.230000019073486 | - | - | - | - | - | LP | YES, a) |
| Genes related to cardiovascular phenotypes | Hypertrophic cardiomyopathy | MYBPC3 | AD | 11:47364239:T:- | - | 2078 | 3 | 0 | 0 | 0.999 | 0.001 | 0.001 | 3 | 0 | 0.144 | frameshift_variant | - | - | 4.630000114440918 | - | - | - | - | - | LP | YES, a) |
| Genes related to cardiovascular phenotypes | Hypertrophic cardiomyopathy | MYBPC3 | AD | 11:47364243:TG:- | - | 2078 | 3 | 0 | 0 | 0.999 | 0.001 | 0.001 | 3 | 0 | 0.144 | frameshift_variant | - | - | 1.4600000381469727 | - | - | - | - | - | LP | YES, a) |
| Genes related to cancer phenotypes | Hereditary breast and/or ovarian cancer | BRCA2 | AD | 13:32906553:CTAAATG | - | 2019 | 3 | 0 | 0 | 0.999 | 0.001 | 0.001 | 3 | 0 | 0.148 | frameshift_variant | - | - | 0.3409999907016754 | - | - | - | - | - | LP | YES, a) |
| Genes related to cancer phenotypes | Hereditary breast and/or ovarian cancer | BRCA2 | AD | 13:32913593:CAACC:- | - | 2019 | 3 | 0 | 0 | 0.999 | 0.001 | 0.001 | 3 | 0 | 0.148 | frameshift_variant | - | - | -0.9390000104904175 | - | - | - | - | - | LP | YES, a) |
| Genes related to cancer phenotypes | Hereditary breast and/or ovarian cancer | BRCA2 | AD | 13:32913970:CA:- | - | 2019 | 3 | 0 | 0 | 0.999 | 0.001 | 0.001 | 3 | 0 | 0.148 | frameshift_variant | - | - | -2.0799999237060547 | - | - | - | - | - | LP | YES, a) |
| Genes related to cancer phenotypes | Hereditary breast and/or ovarian cancer | BRCA2 | AD | 13:32913975:-:AT | - | 2019 | 3 | 0 | 0 | 0.999 | 0.001 | 0.001 | 3 | 0 | 0.148 | frameshift_variant | - | - | -3.1600000858306885 | - | - | - | - | - | LP | YES, a) |
| Genes related to cancer phenotypes | Hereditary breast and/or ovarian cancer | BRCA2 | AD | 13:32914437:GTGG:- | - | 2019 | 3 | 0 | 0 | 0.999 | 0.001 | 0.001 | 3 | 0 | 0.148 | frameshift_variant | - | - | 5.71999979019165 | - | - | - | - | - | LP | YES, a) |
| Genes related to cancer phenotypes | Hereditary breast and/or ovarian cancer | BRCA2 | AD | 13:32914442:-:CAAG | - | 2019 | 3 | 0 | 0 | 0.999 | 0.001 | 0.001 | 3 | 0 | 0.148 | frameshift_variant | - | - | -0.10899999737739563 | - | - | - | - | - | LP | YES, a) |
| Genes related to cancer phenotypes | Hereditary breast and/or ovarian cancer | BRCA2 | AD | 13:32915130:CTAC:- | - | 2019 | 3 | 0 | 0 | 0.999 | 0.001 | 0.001 | 3 | 0 | 0.148 | frameshift_variant | - | - | 4.199999809265137 | - | - | - | - | - | LP | YES, a) |
| Genes related to cancer phenotypes | Hereditary breast and/or ovarian cancer | BRCA2 | AD | 13:32915135:-:CGTC | - | 2019 | 3 | 0 | 0 | 0.999 | 0.001 | 0.001 | 3 | 0 | 0.148 | frameshift_variant | - | - | 2.5999999046325684 | - | - | - | - | - | LP | YES, a) |
| Genes related to cancer phenotypes | Hereditary breast and/or ovarian cancer | BRCA2 | AD | 13:32918763:TTAAA:- | - | 2019 | 3 | 0 | 0 | 0.999 | 0.001 | 0.001 | 3 | 0 | 0.148 | frameshift_variant | - | - | 2.4000000953674316 | - | - | - | - | - | LP | YES, a) |
| Genes related to cancer phenotypes | Hereditary breast and/or ovarian cancer | BRCA2 | AD | 13:32969046:CAACAAA | - | 2019 | 3 | 0 | 0 | 0.999 | 0.001 | 0.001 | 3 | 0 | 0.148 | frameshift_variant | - | - | -3.7100000381469727 | - | - | - | - | - | LP | YES, a) |
| Genes related to cancer phenotypes | Retinoblastoma | RB1 | AD | 13:48916753:AAAAA:- | - | 2019 | 3 | 0 | 0 | 0.999 | 0.001 | 0.001 | 3 | 0 | 0.148 | frameshift_variant | - | - | 5.389999866485596 | - | - | - | - | - | LP | YES, a) |
| Genes related to cancer phenotypes | Retinoblastoma | RB1 | AD | 13:48916758:-:GGGGG | - | 2019 | 3 | 0 | 0 | 0.999 | 0.001 | 0.001 | 3 | 0 | 0.148 | frameshift_variant | - | - | 5.389999866485596 | - | - | - | - | - | LP | YES, a) |
| Genes related to cancer phenotypes | Retinoblastoma | RB1 | AD | 13:49039440:-:TGTTTC | - | 2019 | 3 | 0 | 0 | 0.999 | 0.001 | 0.001 | 3 | 0 | 0.148 | frameshift_variant | - | - | -0.10499999672174454 | - | - | - | - | - | LP | YES, a) |
| Genes related to cancer phenotypes | Retinoblastoma | RB1 | AD | 13:49039442:GAAGAGT | - | 2019 | 3 | 0 | 0 | 0.999 | 0.001 | 0.001 | 3 | 0 | 0.148 | frameshift_variant | - | - | 5.869999885559082 | - | - | - | - | - | LP | YES, a) |
| Genes related to miscellaneous phenotypes | Wilson disease | ATP7B | AR | 13:52518281:G:T | rs76151636 | 2064 | 3 | 0 | 0 | 0.999 | 0.001 | 0.001 | 3 | 0 | 0.145 | missense_variant | 0.0009940357 | - | 2.319999933242798 | 0.0 | 1.0 | 25.100000381469727 | 3848 | Pathogenic | P | NO, b) |
| Genes related to cardiovascular phenotypes | Aortopathies | FBN1 | AD | 15:48726797:A:G | rs1555395001 | 2078 | 3 | 0 | 0 | 0.999 | 0.001 | 0.001 | 3 | 0 | 0.144 | missense_variant | - | - | 6.070000171661377 | 0.0 | 0.999 | 29.799999237060547 | 527154 | Pathogenic/Likely pathogenic | P/LP | YES, a) |
| Genes related to cardiovascular phenotypes | Aortopathies | FBN1 | AD | 15:48905256:-:GTTGT | - | 2078 | 3 | 0 | 0 | 0.999 | 0.001 | 0.001 | 3 | 0 | 0.144 | frameshift_variant | - | - | 5.119999885559082 | - | - | - | - | - | LP | YES, a) |
| Genes related to cardiovascular phenotypes | Aortopathies | SMAD3 | AD | 15:67479723:A:- | - | 2078 | 3 | 0 | 0 | 0.999 | 0.001 | 0.001 | 3 | 0 | 0.144 | frameshift_variant | - | - | 5.409999847412109 | - | - | - | - | - | LP | YES, a) |
| Genes related to cardiovascular phenotypes | Aortopathies | SMAD3 | AD | 15:67479729:-:A | - | 2078 | 3 | 0 | 0 | 0.999 | 0.001 | 0.001 | 3 | 0 | 0.144 | frameshift_variant | - | - | 3.5299999713897705 | - | - | - | - | - | LP | YES, a) |
| Genes related to cancer phenotypes | Hereditary breast and/or ovarian cancer | PALB2 | AD | 16:23625379:CA:- | - | 2019 | 3 | 0 | 0 | 0.999 | 0.001 | 0.001 | 3 | 0 | 0.148 | frameshift_variant | - | - | 5.949999809265137 | - | - | - | - | - | LP | YES, a) |
| Genes related to cancer phenotypes | Hereditary breast and/or ovarian cancer | PALB2 | AD | 16:23625382:-:G | - | 2019 | 3 | 0 | 0 | 0.999 | 0.001 | 0.001 | 3 | 0 | 0.148 | frameshift_variant | - | - | 3.7200000286102295 | - | - | - | - | - | LP | YES, a) |
| Genes related to cancer phenotypes | Hereditary breast and/or ovarian cancer | BRCA1 | AD | 17:41245431:-:GGTTG | - | 2019 | 3 | 0 | 0 | 0.999 | 0.001 | 0.001 | 3 | 0 | 0.148 | frameshift_variant | - | - | 2.950000047683716 | - | - | - | - | - | LP | YES, a) |
| Genes related to cancer phenotypes | Hereditary breast and/or ovarian cancer | BRCA1 | AD | 17:41246201:G:- | rs886039945 | 2019 | 3 | 0 | 0 | 0.999 | 0.001 | 0.001 | 3 | 0 | 0.148 | frameshift_variant | - | - | 3.5799999237060547 | - | - | - | 266159 | Pathogenic | P | YES, a) |
| Genes related to cancer phenotypes | Juvenile polyposis syndrome | SMAD4 | AD | 18:48573481:GTTTGAT | - | 2019 | 3 | 0 | 0 | 0.999 | 0.001 | 0.001 | 3 | 0 | 0.148 | frameshift_variant | - | - | 5.860000133514404 | - | - | - | - | - | LP | YES, a) |
| Genes related to cardiovascular phenotypes | Familial hypercholesterolemia | LDLR | AD | 19:11216228:T:- | rs879254609 | 2029 | 3 | 0 | 1 | 0.999 | 0.001 | 0.001 | 3 | 0 | 0.148 | frameshift_variant | - | - | 5.599999904632568 | - | - | - | 251344 | Pathogenic | P | YES, a) |
| Genes related to cardiovascular phenotypes | Familial hypercholesterolemia | LDLR | AD | 19:11216232:-:T | rs879254614 | 2029 | 3 | 0 | 1 | 0.999 | 0.001 | 0.001 | 3 | 0 | 0.148 | frameshift_variant | - | - | 5.599999904632568 | - | - | - | 251350 | Pathogenic | P | YES, a) |
| Genes related to cardiovascular phenotypes | Dilated cardiomyopathy | TTN | AD | 2:179428045:TAGC:- | - | 2078 | 3 | 0 | 0 | 0.999 | 0.001 | 0.001 | 3 | 0 | 0.144 | frameshift_variant | - | - | 5.739999771118164 | - | - | - | - | - | LP | YES, a) |
| Genes related to cardiovascular phenotypes | Dilated cardiomyopathy | TTN | AD | 2:179464120:AT:- | - | 2078 | 3 | 0 | 0 | 0.999 | 0.001 | 0.001 | 3 | 0 | 0.144 | frameshift_variant | - | - | 5.940000057220459 | - | - | - | - | - | LP | YES, a) |
| Genes related to cardiovascular phenotypes | Dilated cardiomyopathy | TTN | AD | 2:179466241:-:GTTG | - | 2078 | 3 | 0 | 0 | 0.999 | 0.001 | 0.001 | 3 | 0 | 0.144 | frameshift_variant | - | - | 5.739999771118164 | - | - | - | - | - | LP | YES, a) |
| Genes related to cardiovascular phenotypes | Dilated cardiomyopathy | TTN | AD | 2:179481859:CA:- | - | 2078 | 3 | 0 | 0 | 0.999 | 0.001 | 0.001 | 3 | 0 | 0.144 | frameshift_variant | - | - | -3.190000057220459 | - | - | - | - | - | LP | YES, a) |
| Genes related to cardiovascular phenotypes | Dilated cardiomyopathy | TTN | AD | 2:179481862:-:T | - | 2078 | 3 | 0 | 0 | 0.999 | 0.001 | 0.001 | 3 | 0 | 0.144 | frameshift_variant | - | - | 5.760000228881836 | - | - | - | - | - | LP | YES, a) |
| Genes related to cardiovascular phenotypes | Dilated cardiomyopathy | TTN | AD | 2:179481864:-:C | - | 2078 | 3 | 0 | 0 | 0.999 | 0.001 | 0.001 | 3 | 0 | 0.144 | frameshift_variant | - | - | -6.659999847412109 | - | - | - | - | - | LP | YES, a) |
| Genes related to cardiovascular phenotypes | Dilated cardiomyopathy | TTN | AD | 2:179576833:-:CG | - | 2078 | 3 | 0 | 0 | 0.999 | 0.001 | 0.001 | 3 | 0 | 0.144 | frameshift_variant | - | - | 5.650000095367432 | - | - | - | - | - | LP | YES, a) |
| Genes related to cardiovascular phenotypes | Dilated cardiomyopathy | TTN | AD | 2:179576835:-:C | - | 2078 | 3 | 0 | 0 | 0.999 | 0.001 | 0.001 | 3 | 0 | 0.144 | frameshift_variant | - | - | 3.2300000190734863 | - | - | - | - | - | LP | YES, a) |
| Genes related to cardiovascular phenotypes | Dilated cardiomyopathy | TTN | AD | 2:179587997:-:A | - | 2078 | 3 | 0 | 0 | 0.999 | 0.001 | 0.001 | 3 | 0 | 0.144 | frameshift_variant | - | - | -0.39800000190734863 | - | - | - | - | - | LP | YES, a) |
| Genes related to cardiovascular phenotypes | Dilated cardiomyopathy | TTN | AD | 2:179596082:GC:- | - | 2078 | 3 | 0 | 0 | 0.999 | 0.001 | 0.001 | 3 | 0 | 0.144 | frameshift_variant | - | - | 5.71999979019165 | - | - | - | - | - | LP | YES, a) |
| Genes related to cardiovascular phenotypes | Dilated cardiomyopathy | TTN | AD | 2:179596087:-:AC | - | 2078 | 3 | 0 | 0 | 0.999 | 0.001 | 0.001 | 3 | 0 | 0.144 | frameshift_variant | - | - | 4.800000190734863 | - | - | - | - | - | LP | YES, a) |
| Genes related to cardiovascular phenotypes | Dilated cardiomyopathy | TTN | AD | 2:179615332:-:GGCCC | - | 2076 | 3 | 0 | 2 | 0.999 | 0.001 | 0.001 | 3 | 0 | 0.144 | frameshift_variant | - | - | 3.059999942779541 | - | - | - | - | - | LP | YES, a) |
| Genes related to cardiovascular phenotypes | Dilated cardiomyopathy | TTN | AD | 2:179621451:-:G | - | 2078 | 3 | 0 | 0 | 0.999 | 0.001 | 0.001 | 3 | 0 | 0.144 | frameshift_variant | - | - | 3.6600000858306885 | - | - | - | - | - | LP | YES, a) |
| Genes related to cancer phenotypes | Lynch syndrome | MSH2 | AD | 2:47703532:T:- | - | 2019 | 3 | 0 | 0 | 0.999 | 0.001 | 0.001 | 3 | 0 | 0.148 | frameshift_variant | - | - | 6.070000171661377 | - | - | - | - | - | LP | YES, a) |
| Genes related to cancer phenotypes | Lynch syndrome | MSH2 | AD | 2:47703537:-:A | - | 2019 | 3 | 0 | 0 | 0.999 | 0.001 | 0.001 | 3 | 0 | 0.148 | frameshift_variant | - | - | 6.070000171661377 | - | - | - | - | - | LP | YES, a) |
| Genes related to cancer phenotypes | Lynch syndrome | MSH6 | AD | 2:48033453:GT:- | - | 2019 | 3 | 0 | 0 | 0.999 | 0.001 | 0.001 | 3 | 0 | 0.148 | frameshift_variant | - | - | 5.5 | - | - | - | - | - | LP | YES, a) |
| Genes related to cancer phenotypes | Lynch syndrome | MSH6 | AD | 2:48033456:GAAGATT: | - | 2019 | 3 | 0 | 0 | 0.999 | 0.001 | 0.001 | 3 | 0 | 0.148 | frameshift_variant | - | - | 5.5 | - | - | - | - | - | LP | YES, a) |
| Genes related to cancer phenotypes | Neurofibromatosis type 2 | NF2 | AD | 22:30032830:AAGGA:- | - | 2019 | 3 | 0 | 0 | 0.999 | 0.001 | 0.001 | 3 | 0 | 0.148 | frameshift_variant | - | - | 3.9100000858306885 | - | - | - | - | - | LP | YES, a) |
| Genes related to cardiovascular phenotypes | Arrhythmogenic right ventricular cardiomyopathy | DSP | AD | 6:7578014:-:AG | - | 2078 | 3 | 0 | 0 | 0.999 | 0.001 | 0.001 | 3 | 0 | 0.144 | frameshift_variant | - | - | 5.550000190734863 | - | - | - | - | - | LP | YES, a) |
| Genes related to cardiovascular phenotypes | Catecholaminergic polymorphic ventricular tachycardia | RYR2 | AD | 1:237777621:G:- | - | 2077 | 4 | 0 | 0 | 0.999 | 0.001 | 0.001 | 4 | 0 | 0.192 | frameshift_variant | - | - | 3.2899999618530273 | - | - | - | - | - | LP | YES, a) |
| Genes related to cardiovascular phenotypes | Catecholaminergic polymorphic ventricular tachycardia | RYR2 | AD | 1:237777626:-:G | - | 2077 | 4 | 0 | 0 | 0.999 | 0.001 | 0.001 | 4 | 0 | 0.192 | frameshift_variant | - | - | 5.429999828338623 | - | - | - | - | - | LP | YES, a) |
| Genes related to cardiovascular phenotypes | Catecholaminergic polymorphic ventricular tachycardia | RYR2 | AD | 1:237796943:TAGGCAC | - | 2077 | 4 | 0 | 0 | 0.999 | 0.001 | 0.001 | 4 | 0 | 0.192 | frameshift_variant | - | - | 4.769999980926514 | - | - | - | - | - | LP | YES, a) |
| Genes related to cardiovascular phenotypes | Catecholaminergic polymorphic ventricular tachycardia | RYR2 | AD | 1:237796953:-:CCGCTC | - | 2077 | 4 | 0 | 0 | 0.999 | 0.001 | 0.001 | 4 | 0 | 0.192 | frameshift_variant | - | - | 4.900000095367432 | - | - | - | - | - | LP | YES, a) |
| Genes related to cancer phenotypes | Familial medullary thyroid cancer | RET | AD | 10:43622072:A:- | - | 2018 | 4 | 0 | 0 | 0.999 | 0.001 | 0.001 | 4 | 0 | 0.198 | frameshift_variant | - | - | 5.090000152587891 | - | - | - | - | - | LP | YES, a) |
| Genes related to cancer phenotypes | Familial medullary thyroid cancer | RET | AD | 10:43622075:A:- | - | 2018 | 4 | 0 | 0 | 0.999 | 0.001 | 0.001 | 4 | 0 | 0.198 | frameshift_variant | - | - | 5.090000152587891 | - | - | - | - | - | LP | YES, a) |
| Genes related to cancer phenotypes | Familial medullary thyroid cancer | RET | AD | 10:43622077:-:TG | - | 2018 | 4 | 0 | 0 | 0.999 | 0.001 | 0.001 | 4 | 0 | 0.198 | frameshift_variant | - | - | -8.34000015258789 | - | - | - | - | - | LP | YES, a) |
| Genes related to cancer phenotypes | Hereditary breast and/or ovarian cancer | BRCA2 | AD | 13:32906561:-:CGCCCA | - | 2018 | 4 | 0 | 0 | 0.999 | 0.001 | 0.001 | 4 | 0 | 0.198 | frameshift_variant | - | - | 1.7999999523162842 | - | - | - | - | - | LP | YES, a) |
| Genes related to cancer phenotypes | Hereditary breast and/or ovarian cancer | BRCA2 | AD | 13:32969044:-:CCTGGC | - | 2018 | 4 | 0 | 0 | 0.999 | 0.001 | 0.001 | 4 | 0 | 0.198 | frameshift_variant | - | - | -10.300000190734863 | - | - | - | - | - | LP | YES, a) |
| Genes related to cardiovascular phenotypes | Aortopathies | FBN1 | AD | 15:48704807:TC:- | - | 2077 | 4 | 0 | 0 | 0.999 | 0.001 | 0.001 | 4 | 0 | 0.192 | frameshift_variant | - | - | 4.449999809265137 | - | - | - | - | - | LP | YES, a) |
| Genes related to cancer phenotypes | Hereditary breast and/or ovarian cancer | BRCA1 | AD | 17:41245862:AATA:- | - | 2017 | 4 | 0 | 1 | 0.999 | 0.001 | 0.001 | 4 | 0 | 0.198 | frameshift_variant | - | - | 5.070000171661377 | - | - | - | - | - | LP | YES, a) |
| Genes related to cancer phenotypes | Hereditary breast and/or ovarian cancer | BRCA1 | AD | 17:41245867:-:GCGA | - | 2018 | 4 | 0 | 0 | 0.999 | 0.001 | 0.001 | 4 | 0 | 0.198 | frameshift_variant | - | - | 0.40700000524520874 | - | - | - | - | - | LP | YES, a) |
| Genes related to inborn errors of metabolism phenotypes | Pompe disease | GAA | AR | 17:78090815:G:C | rs1800312 | 2075 | 4 | 0 | 0 | 0.999 | 0.001 | 0.001 | 4 | 2 | 0.192 | missense_variant | 0.0009940357 | - | 5.099999904632568 | 0.0 | 1.0 | 33.0 | 265160 | Pathogenic | P | NO, b) |
| Genes related to cardiovascular phenotypes | Arrhythmogenic right ventricular cardiomyopathy | DSG2 | AD | 18:29115333:CAAAA:- | - | 2077 | 4 | 0 | 0 | 0.999 | 0.001 | 0.001 | 4 | 0 | 0.192 | frameshift_variant | - | - | 5.659999847412109 | - | - | - | - | - | LP | YES, a) |
| Genes related to cardiovascular phenotypes | Arrhythmogenic right ventricular cardiomyopathy | DSG2 | AD | 18:29115338:-:TGGGG | - | 2077 | 4 | 0 | 0 | 0.999 | 0.001 | 0.001 | 4 | 0 | 0.192 | frameshift_variant | - | - | 5.659999847412109 | - | - | - | - | - | LP | YES, a) |
| Genes related to cardiovascular phenotypes | Arrhythmogenic right ventricular cardiomyopathy | DSG2 | AD | 18:29125706:A:- | rs1064793103 | 2056 | 4 | 0 | 9 | 0.999 | 0.001 | 0.001 | 4 | 0 | 0.194 | frameshift_variant | - | - | 4.820000171661377 | - | - | - | 418168 | Likely pathogenic | LP | YES, a) |
| Genes related to miscellaneous phenotypes | Malignant hyperthermia | RYR1 | AD | 19:38951151:CCCC:- | - | 2092 | 4 | 0 | 0 | 0.999 | 0.001 | 0.001 | 4 | 0 | 0.191 | frameshift_variant | - | - | 3.069999933242798 | - | - | - | - | - | LP | YES, a) |
| Genes related to miscellaneous phenotypes | Malignant hyperthermia | RYR1 | AD | 19:38951155:-:GGGG | - | 2091 | 4 | 0 | 1 | 0.999 | 0.001 | 0.001 | 4 | 0 | 0.191 | frameshift_variant | - | - | 3.069999933242798 | - | - | - | - | - | LP | YES, a) |
| Genes related to miscellaneous phenotypes | Malignant hyperthermia | RYR1 | AD | 19:39039059:-:G | - | 2075 | 4 | 0 | 5 | 0.999 | 0.001 | 0.001 | 4 | 0 | 0.192 | frameshift_variant | - | - | 4.360000133514404 | - | - | - | - | - | LP | YES, a) |
| Genes related to cardiovascular phenotypes | Dilated cardiomyopathy | TTN | AD | 2:179396328:TG:- | - | 2077 | 4 | 0 | 0 | 0.999 | 0.001 | 0.001 | 4 | 0 | 0.192 | frameshift_variant | - | - | 4.679999828338623 | - | - | - | - | - | LP | YES, a) |
| Genes related to cardiovascular phenotypes | Dilated cardiomyopathy | TTN | AD | 2:179396332:-:TG | - | 2077 | 4 | 0 | 0 | 0.999 | 0.001 | 0.001 | 4 | 0 | 0.192 | frameshift_variant | - | - | 5.559999942779541 | - | - | - | - | - | LP | YES, a) |
| Genes related to cardiovascular phenotypes | Dilated cardiomyopathy | TTN | AD | 2:179468902:CCTGG:- | - | 2077 | 4 | 0 | 0 | 0.999 | 0.001 | 0.001 | 4 | 0 | 0.192 | frameshift_variant | - | - | 6.159999847412109 | - | - | - | - | - | LP | YES, a) |
| Genes related to cardiovascular phenotypes | Dilated cardiomyopathy | TTN | AD | 2:179468907:-:TTCAA | - | 2077 | 4 | 0 | 0 | 0.999 | 0.001 | 0.001 | 4 | 0 | 0.192 | frameshift_variant | - | - | 6.159999847412109 | - | - | - | - | - | LP | YES, a) |
| Genes related to cardiovascular phenotypes | Dilated cardiomyopathy | TTN | AD | 2:179507024:CTTT:- | - | 2077 | 4 | 0 | 0 | 0.999 | 0.001 | 0.001 | 4 | 0 | 0.192 | frameshift_variant | - | - | 5.550000190734863 | - | - | - | - | - | LP | YES, a) |
| Genes related to cardiovascular phenotypes | Dilated cardiomyopathy | TTN | AD | 2:179507033:-:CCCC | - | 2077 | 4 | 0 | 0 | 0.999 | 0.001 | 0.001 | 4 | 0 | 0.192 | frameshift_variant | - | - | 5.550000190734863 | - | - | - | - | - | LP | YES, a) |
| Genes related to cardiovascular phenotypes | Dilated cardiomyopathy | TTN | AD | 2:179580244:T:- | - | 2009 | 4 | 0 | 12 | 0.999 | 0.001 | 0.001 | 4 | 0 | 0.199 | frameshift_variant | - | - | 5.480000019073486 | - | - | - | - | - | LP | YES, a) |
| Genes related to cardiovascular phenotypes | Dilated cardiomyopathy | TTN | AD | 2:179580247:T:- | - | 2009 | 4 | 0 | 12 | 0.999 | 0.001 | 0.001 | 4 | 0 | 0.199 | frameshift_variant | - | - | 5.480000019073486 | - | - | - | - | - | LP | YES, a) |
| Genes related to cardiovascular phenotypes | Dilated cardiomyopathy | TTN | AD | 2:179580249:-:AC | - | 2009 | 4 | 0 | 12 | 0.999 | 0.001 | 0.001 | 4 | 0 | 0.199 | frameshift_variant | - | - | -10.699999809265137 | - | - | - | - | - | LP | YES, a) |
| Genes related to cardiovascular phenotypes | Dilated cardiomyopathy | TTN | AD | 2:179596298:AA:- | - | 2077 | 4 | 0 | 0 | 0.999 | 0.001 | 0.001 | 4 | 0 | 0.192 | frameshift_variant | - | - | 5.590000152587891 | - | - | - | - | - | LP | YES, a) |
| Genes related to cardiovascular phenotypes | Dilated cardiomyopathy | TTN | AD | 2:179605709:AAAGC:- | - | 2077 | 4 | 0 | 0 | 0.999 | 0.001 | 0.001 | 4 | 0 | 0.192 | frameshift_variant | - | - | 1.590000033378601 | - | - | - | - | - | LP | YES, a) |
| Genes related to cardiovascular phenotypes | Dilated cardiomyopathy | TTN | AD | 2:179640402:AGTT:- | - | 2077 | 4 | 0 | 0 | 0.999 | 0.001 | 0.001 | 4 | 0 | 0.192 | frameshift_variant | - | - | -1.5 | - | - | - | - | - | LP | YES, a) |
| Genes related to cancer phenotypes | Neurofibromatosis type 2 | NF2 | AD | 22:30032778:CC:- | - | 2018 | 4 | 0 | 0 | 0.999 | 0.001 | 0.001 | 4 | 0 | 0.198 | frameshift_variant | - | - | 2.5299999713897705 | - | - | - | - | - | LP | YES, a) |
| Genes related to cardiovascular phenotypes | Catecholaminergic polymorphic ventricular tachycardia | RYR2 | AD | 1:237817685:TGGGG:- | - | 2076 | 5 | 0 | 0 | 0.999 | 0.001 | 0.001 | 5 | 0 | 0.240 | frameshift_variant | - | - | 5.860000133514404 | - | - | - | - | - | LP | YES, a) |
| Genes related to cardiovascular phenotypes | Catecholaminergic polymorphic ventricular tachycardia | RYR2 | AD | 1:237817690:-:CAAAA | - | 2076 | 5 | 0 | 0 | 0.999 | 0.001 | 0.001 | 5 | 0 | 0.240 | frameshift_variant | - | - | 5.860000133514404 | - | - | - | - | - | LP | YES, a) |
| Genes related to cancer phenotypes | MUTYH-associated polyposis | MUTYH | AR | 1:45798475:T:C | rs34612342 | 2017 | 5 | 0 | 0 | 0.999 | 0.001 | 0.001 | 5 | 3 | 0.247 | missense_variant | - | - | 5.010000228881836 | 0.0 | 1.0 | 24.700000762939453 | 5293 | Pathogenic/Likely pathogenic | P/LP | NO, b) |
| Genes related to cancer phenotypes | Hereditary paraganglioma–pheochromocytoma syndrome | SDHAF2 | AD | 11:61205475:G:C | - | 2017 | 5 | 0 | 0 | 0.999 | 0.001 | 0.001 | 5 | 0 | 0.247 | splice_acceptor_variant | - | - | 5.0 | - | - | 23.799999237060547 | - | - | P | YES, a) |
| Genes related to cancer phenotypes | Hereditary breast and/or ovarian cancer | BRCA2 | AD | 13:32911776:AGGA:- | - | 2017 | 5 | 0 | 0 | 0.999 | 0.001 | 0.001 | 5 | 0 | 0.247 | frameshift_variant | - | - | 3.309999942779541 | - | - | - | - | - | LP | YES, a) |
| Genes related to cancer phenotypes | Hereditary breast and/or ovarian cancer | BRCA2 | AD | 13:32912662:GGAA:- | - | 2017 | 5 | 0 | 0 | 0.999 | 0.001 | 0.001 | 5 | 0 | 0.247 | frameshift_variant | - | - | 3.5 | - | - | - | - | - | LP | YES, a) |
| Genes related to cancer phenotypes | Hereditary breast and/or ovarian cancer | BRCA2 | AD | 13:32912668:-:CCTG | - | 2017 | 5 | 0 | 0 | 0.999 | 0.001 | 0.001 | 5 | 0 | 0.247 | frameshift_variant | - | - | 3.5899999141693115 | - | - | - | - | - | LP | YES, a) |
| Genes related to cardiovascular phenotypes | Aortopathies | FBN1 | AD | 15:48802346:-:A | - | 2076 | 5 | 0 | 0 | 0.999 | 0.001 | 0.001 | 5 | 0 | 0.240 | frameshift_variant | - | - | 5.5 | - | - | - | - | - | LP | YES, a) |
| Genes related to cardiovascular phenotypes | Aortopathies | MYH11 | AD | 16:15815365:CTTCC:- | - | 2076 | 5 | 0 | 0 | 0.999 | 0.001 | 0.001 | 5 | 0 | 0.240 | frameshift_variant | - | - | 1.7799999713897705 | - | - | - | - | - | LP | YES, a) |
| Genes related to cancer phenotypes | Juvenile polyposis syndrome | SMAD4 | AD | 18:48591841:-:AA | - | 2017 | 5 | 0 | 0 | 0.999 | 0.001 | 0.001 | 5 | 0 | 0.247 | frameshift_variant | - | - | 5.989999771118164 | - | - | - | - | - | LP | YES, a) |
| Genes related to cardiovascular phenotypes | Dilated cardiomyopathy | TTN | AD | 2:179406260:-:TA | - | 2076 | 5 | 0 | 0 | 0.999 | 0.001 | 0.001 | 5 | 0 | 0.240 | frameshift_variant | - | - | 2.740000009536743 | - | - | - | - | - | LP | YES, a) |
| Genes related to cardiovascular phenotypes | Dilated cardiomyopathy | TTN | AD | 2:179406261:-:G | - | 2076 | 5 | 0 | 0 | 0.999 | 0.001 | 0.001 | 5 | 0 | 0.240 | frameshift_variant | - | - | 5.559999942779541 | - | - | - | - | - | LP | YES, a) |
| Genes related to cardiovascular phenotypes | Catecholaminergic polymorphic ventricular tachycardia | RYR2 | AD | 1:237433889:T:- | - | 2075 | 6 | 0 | 0 | 0.999 | 0.001 | 0.001 | 6 | 0 | 0.288 | frameshift_variant | - | - | -1.0199999809265137 | - | - | - | - | - | LP | YES, a) |
| Genes related to cancer phenotypes | Retinoblastoma | RB1 | AD | 13:48881456:T:- | - | 2016 | 6 | 0 | 0 | 0.999 | 0.001 | 0.001 | 6 | 0 | 0.297 | frameshift_variant | - | - | 3.6500000953674316 | - | - | - | - | - | LP | YES, a) |
| Genes related to cancer phenotypes | Retinoblastoma | RB1 | AD | 13:48881457:T:A | - | 2016 | 6 | 0 | 0 | 0.999 | 0.001 | 0.001 | 6 | 0 | 0.297 | stop_gained | - | - | 4.829999923706055 | - | - | 36.0 | - | - | P | YES, a) |
| Genes related to cardiovascular phenotypes | Hypertrophic cardiomyopathy | ACTC1 | AD | 15:35085762:CATA:- | - | 2047 | 6 | 0 | 16 | 0.999 | 0.001 | 0.001 | 6 | 0 | 0.292 | frameshift_variant | - | - | 2.4100000858306885 | - | - | - | - | - | LP | YES, a) |
| Genes related to cardiovascular phenotypes | Hypertrophic cardiomyopathy | ACTC1 | AD | 15:35085767:-:CGCC | - | 2047 | 6 | 0 | 16 | 0.999 | 0.001 | 0.001 | 6 | 0 | 0.292 | frameshift_variant | - | - | 5.489999771118164 | - | - | - | - | - | LP | YES, a) |
| Genes related to cardiovascular phenotypes | Aortopathies | FBN1 | AD | 15:48786443:-:GTTGT | - | 2075 | 6 | 0 | 0 | 0.999 | 0.001 | 0.001 | 6 | 0 | 0.288 | frameshift_variant | - | - | 5.78000020980835 | - | - | - | - | - | LP | YES, a) |
| Genes related to cardiovascular phenotypes | Aortopathies | FBN1 | AD | 15:48905283:A:- | - | 2075 | 6 | 0 | 0 | 0.999 | 0.001 | 0.001 | 6 | 0 | 0.288 | frameshift_variant | - | - | 5.119999885559082 | - | - | - | - | - | LP | YES, a) |
| Genes related to cancer phenotypes | Tuberous sclerosis complex | TSC2 | AD | 16:2129352:G:- | - | 2016 | 6 | 0 | 0 | 0.999 | 0.001 | 0.001 | 6 | 0 | 0.297 | frameshift_variant | - | - | 1.5499999523162842 | - | - | - | - | - | LP | YES, a) |
| Genes related to cancer phenotypes | Tuberous sclerosis complex | TSC2 | AD | 16:2129355:G:- | - | 2016 | 6 | 0 | 0 | 0.999 | 0.001 | 0.001 | 6 | 0 | 0.297 | frameshift_variant | - | - | -6.070000171661377 | - | - | - | - | - | LP | YES, a) |
| Genes related to cancer phenotypes | Juvenile polyposis syndrome | SMAD4 | AD | 18:48591831:-:GGCA | - | 2016 | 6 | 0 | 0 | 0.999 | 0.001 | 0.001 | 6 | 0 | 0.297 | frameshift_variant | - | - | 5.989999771118164 | - | - | - | - | - | LP | YES, a) |
| Genes related to cancer phenotypes | Juvenile polyposis syndrome | SMAD4 | AD | 18:48591836:-:C | - | 2016 | 6 | 0 | 0 | 0.999 | 0.001 | 0.001 | 6 | 0 | 0.297 | frameshift_variant | - | - | 5.989999771118164 | - | - | - | - | - | LP | YES, a) |
| Genes related to cardiovascular phenotypes | Dilated cardiomyopathy | TTN | AD | 2:179401193:-:C | - | 2075 | 6 | 0 | 0 | 0.999 | 0.001 | 0.001 | 6 | 0 | 0.288 | frameshift_variant | - | - | 5.760000228881836 | - | - | - | - | - | LP | YES, a) |
| Genes related to cardiovascular phenotypes | Dilated cardiomyopathy | TTN | AD | 2:179425854:TG:- | - | 2075 | 6 | 0 | 0 | 0.999 | 0.001 | 0.001 | 6 | 0 | 0.288 | frameshift_variant | - | - | 5.75 | - | - | - | - | - | LP | YES, a) |
| Genes related to cardiovascular phenotypes | Dilated cardiomyopathy | TTN | AD | 2:179428064:CA:- | - | 2075 | 6 | 0 | 0 | 0.999 | 0.001 | 0.001 | 6 | 0 | 0.288 | frameshift_variant | - | - | -4.400000095367432 | - | - | - | - | - | LP | YES, a) |

| | | | | | | | | | | | | | | | | | | | | | | | | | | |
|---|---|---|---|---|---|---|---|---|---|---|---|---|---|---|---|---|---|---|---|---|---|---|---|---|---|---|
| Genes related to cardiovascular phenotypes | Dilated cardiomyopathy | TTN | AD | 2:179500950:ACAAA:- | - | 2075 | 6 | 0 | 0 | 0.999 | 0.001 | 0.001 | 6 | 0 | 0.288 | frameshift_variant | - | - | 5.619999885559082 | - | - | - | - | - | LP | YES, a) |
| Genes related to cardiovascular phenotypes | Dilated cardiomyopathy | TTN | AD | 2:179641467:T:- | - | 2075 | 6 | 0 | 0 | 0.999 | 0.001 | 0.001 | 6 | 0 | 0.288 | frameshift_variant | - | - | 0.38100001215934753 | - | - | - | - | - | LP | YES, a) |
| Genes related to cardiovascular phenotypes | Dilated cardiomyopathy | TTN | AD | 2:179647295:CACG:- | - | 2075 | 6 | 0 | 0 | 0.999 | 0.001 | 0.001 | 6 | 0 | 0.288 | frameshift_variant | - | - | -6.929999828338623 | - | - | - | - | - | LP | YES, a) |
| Genes related to cardiovascular phenotypes | Dilated cardiomyopathy | TTN | AD | 2:179647690:-:GGT | - | 2075 | 6 | 0 | 0 | 0.999 | 0.001 | 0.001 | 6 | 0 | 0.288 | inframe_insertion | - | - | 6.170000076293945 | - | - | - | - | - | LP | YES, a) |
| Genes related to cardiovascular phenotypes | Arrhythmogenic right ventricular cardiomyopathy | PKP2 | AD | 12:33031456:CATAA:- | - | 2074 | 7 | 0 | 0 | 0.998 | 0.002 | 0.002 | 7 | 0 | 0.336 | frameshift_variant | - | - | -6.320000171661377 | - | - | - | - | - | LP | YES, a) |
| Genes related to cardiovascular phenotypes | Arrhythmogenic right ventricular cardiomyopathy | PKP2 | AD | 12:33031461:-:TGCGG | - | 2074 | 7 | 0 | 0 | 0.998 | 0.002 | 0.002 | 7 | 0 | 0.336 | frameshift_variant | - | - | -6.320000171661377 | - | - | - | - | - | LP | YES, a) |
| Genes related to cancer phenotypes | Hereditary breast and/or ovarian cancer | BRCA1 | AD | 17:41245877:-:AAGGG | - | 2015 | 7 | 0 | 0 | 0.998 | 0.002 | 0.002 | 7 | 0 | 0.346 | frameshift_variant | - | - | -0.40799999237060547 | - | - | - | - | - | LP | YES, a) |
| Genes related to cancer phenotypes | Juvenile polyposis syndrome | SMAD4 | AD | 18:48591826:AAAT:- | - | 2015 | 7 | 0 | 0 | 0.998 | 0.002 | 0.002 | 7 | 0 | 0.346 | frameshift_variant | - | - | 5.989999771118164 | - | - | - | - | - | LP | YES, a) |
| Genes related to cardiovascular phenotypes | Dilated cardiomyopathy | TTN | AD | 2:179411893:AATT:- | - | 2074 | 7 | 0 | 0 | 0.998 | 0.002 | 0.002 | 7 | 0 | 0.336 | frameshift_variant | - | - | 3.240000009536743 | - | - | - | - | - | LP | YES, a) |
| Genes related to cardiovascular phenotypes | Dilated cardiomyopathy | TTN | AD | 2:179477751:-:CAGC | - | 2074 | 7 | 0 | 0 | 0.998 | 0.002 | 0.002 | 7 | 0 | 0.336 | frameshift_variant | - | - | -1.25 | - | - | - | - | - | LP | YES, a) |
| Genes related to cardiovascular phenotypes | Dilated cardiomyopathy | TTN | AD | 2:179644868:A:- | - | 2074 | 7 | 0 | 0 | 0.998 | 0.002 | 0.002 | 7 | 0 | 0.336 | frameshift_variant | - | - | 2.109999895095825 | - | - | - | - | - | LP | YES, a) |
| Genes related to cardiovascular phenotypes | Dilated cardiomyopathy | TTN | AD | 2:179644872:GC:- | - | 2074 | 7 | 0 | 0 | 0.998 | 0.002 | 0.002 | 7 | 0 | 0.336 | frameshift_variant | - | - | 3.9800000190734863 | - | - | - | - | - | LP | YES, a) |
| Genes related to cardiovascular phenotypes | Dilated cardiomyopathy | TTN | AD | 2:179647300:-:TGCC | - | 2074 | 7 | 0 | 0 | 0.998 | 0.002 | 0.002 | 7 | 0 | 0.336 | frameshift_variant | - | - | 4.559999942779541 | - | - | - | - | - | LP | YES, a) |
| Genes related to cardiovascular phenotypes | Dilated cardiomyopathy | TTN | AD | 2:179647764:CTTCT:- | - | 2074 | 7 | 0 | 0 | 0.998 | 0.002 | 0.002 | 7 | 0 | 0.336 | frameshift_variant | - | - | -2.259999990463257 | - | - | - | - | - | LP | YES, a) |
| Genes related to cardiovascular phenotypes | Arrhythmogenic right ventricular cardiomyopathy | DSP | AD | 6:7581764:AGACA:- | - | 2074 | 7 | 0 | 0 | 0.998 | 0.002 | 0.002 | 7 | 0 | 0.336 | frameshift_variant | - | - | 6.019999980926514 | - | - | - | - | - | LP | YES, a) |
| Genes related to cancer phenotypes | PTEN hamartoma tumor syndrome | PTEN | AD | 10:89690818:T:- | rs1564828960 | 2014 | 8 | 0 | 0 | 0.998 | 0.002 | 0.002 | 8 | 0 | 0.396 | frameshift_variant | - | - | 5.619999885559082 | - | - | - | 623213 | Likely pathogenic | LP | YES, a) |
| Genes related to cancer phenotypes | PTEN hamartoma tumor syndrome | PTEN | AD | 10:89690822:-:A | - | 2014 | 8 | 0 | 0 | 0.998 | 0.002 | 0.002 | 8 | 0 | 0.396 | frameshift_variant | - | - | 5.619999885559082 | - | - | - | - | - | LP | YES, a) |
| Genes related to cancer phenotypes | Tuberous sclerosis complex | TSC2 | AD | 16:2129336:AC:- | - | 2014 | 8 | 0 | 0 | 0.998 | 0.002 | 0.002 | 8 | 0 | 0.396 | frameshift_variant | - | - | -0.10599999874830246 | - | - | - | - | - | LP | YES, a) |
| Genes related to cancer phenotypes | Tuberous sclerosis complex | TSC2 | AD | 16:2129342:-:TA | - | 2014 | 8 | 0 | 0 | 0.998 | 0.002 | 0.002 | 8 | 0 | 0.396 | frameshift_variant | - | - | 2.7100000381469727 | - | - | - | - | - | LP | YES, a) |
| Genes related to cancer phenotypes | Lynch syndrome | MSH2 | AD | 2:47641423:CT:- | - | 2014 | 8 | 0 | 0 | 0.998 | 0.002 | 0.002 | 8 | 0 | 0.396 | frameshift_variant | - | - | 5.239999771118164 | - | - | - | - | - | LP | YES, a) |
| Genes related to cancer phenotypes | Lynch syndrome | MSH2 | AD | 2:47641428:-:AG | - | 2014 | 8 | 0 | 0 | 0.998 | 0.002 | 0.002 | 8 | 0 | 0.396 | frameshift_variant | - | - | 3.450000047683716 | - | - | - | - | - | LP | YES, a) |
| Genes related to cardiovascular phenotypes | Catecholaminergic polymorphic ventricular tachycardia | TRDN | AR | 6:123892252:-:A | - | 2073 | 8 | 0 | 0 | 0.998 | 0.002 | 0.002 | 8 | 0 | 0.384 | frameshift_variant | - | - | 5.820000171661377 | - | - | - | - | - | LP | NO, b) |
| Genes related to cardiovascular phenotypes | Arrhythmogenic right ventricular cardiomyopathy | DSP | AD | 6:7581774:-:CCTTT | - | 2073 | 8 | 0 | 0 | 0.998 | 0.002 | 0.002 | 8 | 0 | 0.384 | frameshift_variant | - | - | 6.019999980926514 | - | - | - | - | - | LP | YES, a) |
| Genes related to cancer phenotypes | Lynch syndrome | PMS2 | AD | 7:6038890:-:GTTC | - | 2014 | 8 | 0 | 0 | 0.998 | 0.002 | 0.002 | 8 | 0 | 0.396 | frameshift_variant | - | - | 0.24500000476837158 | - | - | - | - | - | LP | YES, a) |
| Genes related to cancer phenotypes | Lynch syndrome | PMS2 | AD | 7:6038891:C:- | - | 2014 | 8 | 0 | 0 | 0.998 | 0.002 | 0.002 | 8 | 0 | 0.396 | frameshift_variant | - | - | 1.5399999618530273 | - | - | - | - | - | LP | YES, a) |
| Genes related to cancer phenotypes | Hereditary paraganglioma–pheochromocytoma syndrome | SDHC | AD | 1:161326481:G:- | - | 1941 | 9 | 0 | 14 | 0.998 | 0.002 | 0.002 | 9 | 0 | 0.462 | frameshift_variant | - | - | 5.239999771118164 | - | - | - | - | - | LP | YES, a) |
| Genes related to cancer phenotypes | Hereditary paraganglioma–pheochromocytoma syndrome | SDHC | AD | 1:161326484:AT:- | - | 1941 | 9 | 0 | 14 | 0.998 | 0.002 | 0.002 | 9 | 0 | 0.462 | frameshift_variant | - | - | 5.239999771118164 | - | - | - | - | - | LP | YES, a) |
| Genes related to cardiovascular phenotypes | Dilated cardiomyopathy | TTN | AD | 2:179462325:-:A | - | 2072 | 9 | 0 | 0 | 0.998 | 0.002 | 0.002 | 9 | 0 | 0.432 | frameshift_variant | - | - | 6.059999942779541 | - | - | - | - | - | LP | YES, a) |
| Genes related to cardiovascular phenotypes | Dilated cardiomyopathy | TTN | AD | 2:179585900:CTTCT:- | - | 2072 | 9 | 0 | 0 | 0.998 | 0.002 | 0.002 | 9 | 0 | 0.432 | frameshift_variant | - | - | -2.0999999046325684 | - | - | - | - | - | LP | YES, a) |
| Genes related to cancer phenotypes | Lynch syndrome | MSH2 | AD | 2:47641413:AGTT:- | - | 1967 | 9 | 0 | 0 | 0.998 | 0.002 | 0.002 | 9 | 0 | 0.455 | frameshift_variant | - | - | 1.5 | - | - | - | - | - | LP | YES, a) |
| Genes related to cancer phenotypes | Lynch syndrome | MSH2 | AD | 2:47641418:-:CAAA | - | 1967 | 9 | 0 | 0 | 0.998 | 0.002 | 0.002 | 9 | 0 | 0.455 | frameshift_variant | - | - | 1.149999976158142 | - | - | - | - | - | LP | YES, a) |
| Genes related to cardiovascular phenotypes | Aortopathies | FBN1 | AD | 15:48758030:-:CT | - | 2071 | 10 | 0 | 0 | 0.998 | 0.002 | 0.002 | 10 | 0 | 0.481 | frameshift_variant | - | - | 5.739999771118164 | - | - | - | - | - | LP | YES, a) |
| Genes related to cardiovascular phenotypes | Dilated cardiomyopathy | TTN | AD | 2:179462320:A:- | - | 2071 | 10 | 0 | 0 | 0.998 | 0.002 | 0.002 | 10 | 0 | 0.481 | frameshift_variant | - | - | 4.869999885559082 | - | - | - | - | - | LP | YES, a) |
| Genes related to cardiovascular phenotypes | Dilated cardiomyopathy | TTN | AD | 2:179615322:AAAA:- | - | 2070 | 10 | 0 | 1 | 0.998 | 0.002 | 0.002 | 10 | 0 | 0.481 | frameshift_variant | - | - | 3.1600000858306885 | - | - | - | - | - | LP | YES, a) |
| Genes related to cancer phenotypes | Neurofibromatosis type 2 | NF2 | AD | 22:30032834:ACAC:- | - | 2012 | 10 | 0 | 0 | 0.998 | 0.002 | 0.002 | 10 | 0 | 0.495 | frameshift_variant | - | - | 5.010000228881836 | - | - | - | - | - | LP | YES, a) |
| Genes related to cancer phenotypes | Neurofibromatosis type 2 | NF2 | AD | 22:30032840:-:TGTC | - | 2012 | 10 | 0 | 0 | 0.998 | 0.002 | 0.002 | 10 | 0 | 0.495 | frameshift_variant | - | - | 3.940000057220459 | - | - | - | - | - | LP | YES, a) |
| Genes related to cardiovascular phenotypes | Catecholaminergic polymorphic ventricular tachycardia | TRDN | AR | 6:123892248:C:- | - | 2071 | 10 | 0 | 0 | 0.998 | 0.002 | 0.002 | 10 | 0 | 0.481 | frameshift_variant | - | - | 5.820000171661377 | - | - | - | - | - | LP | NO, b) |
| Genes related to cancer phenotypes | Hereditary breast and/or ovarian cancer | BRCA2 | AD | 13:32905057:-:G | - | 2011 | 11 | 0 | 0 | 0.997 | 0.003 | 0.003 | 11 | 0 | 0.544 | frameshift_variant | - | - | 2.119999885559082 | - | - | - | - | - | LP | YES, a) |
| Genes related to cardiovascular phenotypes | Aortopathies | FBN1 | AD | 15:48758035:-:A | - | 2070 | 11 | 0 | 0 | 0.997 | 0.003 | 0.003 | 11 | 0 | 0.529 | frameshift_variant | - | - | 5.739999771118164 | - | - | - | - | - | LP | YES, a) |
| Genes related to miscellaneous phenotypes | Malignant hyperthermia | RYR1 | AD | 19:39077195:TGAG:- | - | 2085 | 11 | 0 | 0 | 0.997 | 0.003 | 0.003 | 11 | 0 | 0.525 | frameshift_variant | - | - | 2.5 | - | - | - | - | - | LP | YES, a) |
| Genes related to cardiovascular phenotypes | Dilated cardiomyopathy | TTN | AD | 2:179500956:-:TGGGG | - | 2070 | 11 | 0 | 0 | 0.997 | 0.003 | 0.003 | 11 | 0 | 0.529 | frameshift_variant | - | - | 0.7279999852180481 | - | - | - | - | - | LP | YES, a) |
| Genes related to cardiovascular phenotypes | Long QT syndrome types 1 and 2 | KCNQ1 | AD | 11:2790119:GC:- | - | 2069 | 12 | 0 | 0 | 0.997 | 0.003 | 0.003 | 12 | 0 | 0.577 | frameshift_variant | - | - | 4.260000228881836 | - | - | - | - | - | LP | YES, a) |
| Genes related to cardiovascular phenotypes | Dilated cardiomyopathy | TTN | AD | 2:179571327:-:AGGGA | - | 2069 | 12 | 0 | 0 | 0.997 | 0.003 | 0.003 | 12 | 0 | 0.577 | frameshift_variant | - | - | 3.680000066757202 | - | - | - | - | - | LP | YES, a) |
| Genes related to cardiovascular phenotypes | Dilated cardiomyopathy | TTN | AD | 2:179583305:-:CC | - | 2069 | 12 | 0 | 0 | 0.997 | 0.003 | 0.003 | 12 | 0 | 0.577 | frameshift_variant | - | - | -5.090000152587891 | - | - | - | - | - | LP | YES, a) |
| Genes related to cardiovascular phenotypes | Dilated cardiomyopathy | TTN | AD | 2:179647310:CTTT:- | - | 2025 | 12 | 0 | 0 | 0.997 | 0.003 | 0.003 | 12 | 0 | 0.589 | frameshift_variant | - | - | 3.130000114440918 | - | - | - | - | - | LP | YES, a) |
| Genes related to cardiovascular phenotypes | Dilated cardiomyopathy | TTN | AD | 2:179647315:-:CCCC | - | 1997 | 12 | 0 | 16 | 0.997 | 0.003 | 0.003 | 12 | 0 | 0.597 | frameshift_variant | - | - | 5.539999961853027 | - | - | - | - | - | LP | YES, a) |
| Genes related to cancer phenotypes | Lynch syndrome | MSH6 | AD | 2:48026142:C:- | - | 2010 | 12 | 0 | 0 | 0.997 | 0.003 | 0.003 | 12 | 0 | 0.593 | frameshift_variant | - | - | 3.509999990463257 | - | - | - | - | - | LP | YES, a) |
| Genes related to cancer phenotypes | Lynch syndrome | MSH6 | AD | 2:48026146:-:C | - | 2010 | 12 | 0 | 0 | 0.997 | 0.003 | 0.003 | 12 | 0 | 0.593 | frameshift_variant | - | - | 3.25 | - | - | - | - | - | LP | YES, a) |
| Genes related to cancer phenotypes | Familial adenomatous polyposis | APC | AD | 5:112103024:-:TG | - | 2010 | 12 | 0 | 0 | 0.997 | 0.003 | 0.003 | 12 | 0 | 0.593 | frameshift_variant | - | - | 5.570000171661377 | - | - | - | - | - | LP | YES, a) |
| Genes related to cancer phenotypes | Familial adenomatous polyposis | APC | AD | 5:112103026:-:G | - | 2010 | 12 | 0 | 0 | 0.997 | 0.003 | 0.003 | 12 | 0 | 0.593 | frameshift_variant | - | - | 4.420000076293945 | - | - | - | - | - | LP | YES, a) |
| Genes related to cardiovascular phenotypes | Catecholaminergic polymorphic ventricular tachycardia | TRDN | AR | 6:123892257:T:- | - | 2069 | 12 | 0 | 0 | 0.997 | 0.003 | 0.003 | 12 | 0 | 0.577 | frameshift_variant | - | - | 5.820000171661377 | - | - | - | - | - | LP | NO, b) |
| Genes related to cardiovascular phenotypes | Long QT syndrome types 1 and 2 | KCNQ1 | AD | 11:2790122:G:- | - | 2068 | 13 | 0 | 0 | 0.997 | 0.003 | 0.003 | 13 | 0 | 0.625 | frameshift_variant | - | - | 3.3399999141693115 | - | - | - | - | - | LP | YES, a) |
| Genes related to cancer phenotypes | Hereditary breast and/or ovarian cancer | BRCA2 | AD | 13:32910422:AAAA:- | - | 1976 | 13 | 0 | 21 | 0.997 | 0.003 | 0.003 | 13 | 0 | 0.654 | frameshift_variant | - | - | 5.519999980926514 | - | - | - | - | - | LP | YES, a) |
| Genes related to cancer phenotypes | Hereditary breast and/or ovarian cancer | BRCA2 | AD | 13:32910427:-:GGGA | - | 1976 | 13 | 0 | 21 | 0.997 | 0.003 | 0.003 | 13 | 0 | 0.654 | frameshift_variant | - | - | 0.9670000076293945 | - | - | - | - | - | LP | YES, a) |
| Genes related to cancer phenotypes | Juvenile polyposis syndrome | SMAD4 | AD | 18:48575107:TGGC:- | - | 2009 | 13 | 0 | 0 | 0.997 | 0.003 | 0.003 | 13 | 0 | 0.643 | frameshift_variant | - | - | 5.320000171661377 | - | - | - | - | - | LP | YES, a) |
| Genes related to miscellaneous phenotypes | Malignant hyperthermia | RYR1 | AD | 19:38996553:-:GG | - | 2082 | 13 | 0 | 1 | 0.997 | 0.003 | 0.003 | 13 | 0 | 0.621 | frameshift_variant | - | - | 4.46999979019165 | - | - | - | - | - | LP | YES, a) |
| Genes related to miscellaneous phenotypes | Malignant hyperthermia | RYR1 | AD | 19:38996555:-:G | - | 2082 | 13 | 0 | 1 | 0.997 | 0.003 | 0.003 | 13 | 0 | 0.621 | frameshift_variant | - | - | 3.430000066757202 | - | - | - | - | - | LP | YES, a) |
| Genes related to cardiovascular phenotypes | Catecholaminergic polymorphic ventricular tachycardia | RYR2 | AD | 1:237951397:-:T | - | 2067 | 14 | 0 | 0 | 0.997 | 0.003 | 0.003 | 14 | 0 | 0.673 | frameshift_variant | - | - | -4.389999866485596 | - | - | - | - | - | LP | YES, a) |
| Genes related to cardiovascular phenotypes | Catecholaminergic polymorphic ventricular tachycardia | RYR2 | AD | 1:237951404:-:AG | - | 2067 | 14 | 0 | 0 | 0.997 | 0.003 | 0.003 | 14 | 0 | 0.673 | frameshift_variant | - | - | 4.550000190734863 | - | - | - | - | - | LP | YES, a) |
| Genes related to cancer phenotypes | Hereditary breast and/or ovarian cancer | BRCA2 | AD | 13:32950875:-:C | - | 2008 | 14 | 0 | 0 | 0.997 | 0.003 | 0.003 | 14 | 0 | 0.692 | frameshift_variant | - | - | 1.4600000381469727 | - | - | - | - | - | LP | YES, a) |
| Genes related to cancer phenotypes | Hereditary breast and/or ovarian cancer | BRCA2 | AD | 13:32950876:-:T | - | 2008 | 14 | 0 | 0 | 0.997 | 0.003 | 0.003 | 14 | 0 | 0.692 | frameshift_variant | - | - | 5.320000171661377 | - | - | - | - | - | LP | YES, a) |
| Genes related to cardiovascular phenotypes | Dilated cardiomyopathy | TTN | AD | 2:179577510:-:C | - | 2067 | 14 | 0 | 0 | 0.997 | 0.003 | 0.003 | 14 | 0 | 0.673 | frameshift_variant | - | - | -7.679999828338623 | - | - | - | - | - | LP | YES, a) |
| Genes related to cancer phenotypes | Neurofibromatosis type 2 | NF2 | AD | 22:30050692:-:TTAAA | - | 2008 | 14 | 0 | 0 | 0.997 | 0.003 | 0.003 | 14 | 0 | 0.692 | frameshift_variant | - | - | 5.989999771118164 | - | - | - | - | - | LP | YES, a) |
| Genes related to cardiovascular phenotypes | Catecholaminergic polymorphic ventricular tachycardia | TRDN | AR | 6:123892262:-:T | - | 2067 | 14 | 0 | 0 | 0.997 | 0.003 | 0.003 | 14 | 0 | 0.673 | frameshift_variant | - | - | -1.75 | - | - | - | - | - | LP | NO, b) |
| Genes related to cancer phenotypes | Hereditary breast and/or ovarian cancer | BRCA2 | AD | 13:32950871:AG:- | - | 2007 | 15 | 0 | 0 | 0.996 | 0.004 | 0.004 | 15 | 0 | 0.742 | frameshift_variant | - | - | 5.320000171661377 | - | - | - | - | - | LP | YES, a) |
| Genes related to cardiovascular phenotypes | Aortopathies | FBN1 | AD | 15:48729268:TCCATAT | - | 2066 | 15 | 0 | 0 | 0.996 | 0.004 | 0.004 | 15 | 0 | 0.721 | frameshift_variant | - | - | 5.769999980926514 | - | - | - | - | - | LP | YES, a) |
| Genes related to cardiovascular phenotypes | Dilated cardiomyopathy | TTN | AD | 2:179604087:GGTGT:- | - | 2066 | 15 | 0 | 0 | 0.996 | 0.004 | 0.004 | 15 | 0 | 0.721 | frameshift_variant | - | - | -10.300000190734863 | - | - | - | - | - | LP | YES, a) |
| Genes related to cardiovascular phenotypes | Aortopathies | FBN1 | AD | 15:48729265:-:AGCAG[illegible] | - | 2065 | 16 | 0 | 0 | 0.996 | 0.004 | 0.004 | 16 | 0 | 0.769 | frameshift_variant | - | - | -1.75 | - | - | - | - | - | LP | YES, a) |
| Genes related to cardiovascular phenotypes | Dilated cardiomyopathy | TTN | AD | 2:179604092:-:AACAC | - | 2065 | 16 | 0 | 0 | 0.996 | 0.004 | 0.004 | 16 | 0 | 0.769 | frameshift_variant | - | - | -10.300000190734863 | - | - | - | - | - | LP | YES, a) |
| Genes related to cardiovascular phenotypes | Dilated cardiomyopathy | TTN | AD | 2:179582861:G:- | - | 2064 | 17 | 0 | 0 | 0.996 | 0.004 | 0.004 | 17 | 0 | 0.817 | frameshift_variant | - | - | 6.159999847412109 | - | - | - | - | - | LP | YES, a) |
| Genes related to cardiovascular phenotypes | Dilated cardiomyopathy | TTN | AD | 2:179582865:TT:- | - | 2064 | 17 | 0 | 0 | 0.996 | 0.004 | 0.004 | 17 | 0 | 0.817 | frameshift_variant | - | - | 4.949999809265137 | - | - | - | - | - | LP | YES, a) |
| Genes related to cardiovascular phenotypes | Dilated cardiomyopathy | TTN | AD | 2:179582867:-:ACC | - | 2064 | 17 | 0 | 0 | 0.996 | 0.004 | 0.004 | 17 | 0 | 0.817 | inframe_insertion | - | - | 4.949999809265137 | - | - | - | - | - | LP | YES, a) |
| Genes related to cancer phenotypes | Hereditary breast and/or ovarian cancer | BRCA2 | AD | 13:32915297:-:G | - | 2002 | 20 | 0 | 0 | 0.995 | 0.005 | 0.005 | 20 | 0 | 0.989 | frameshift_variant | - | - | 3.2200000286102295 | - | - | - | - | - | LP | YES, a) |
| Genes related to cardiovascular phenotypes | Arrhythmogenic right ventricular cardiomyopathy | DSP | AD | 6:7572225:GGATG:- | - | 2058 | 23 | 0 | 0 | 0.994 | 0.006 | 0.006 | 23 | 0 | 1.105 | frameshift_variant | - | - | 4.679999828338623 | - | - | - | - | - | LP | YES, a) |
| Genes related to cardiovascular phenotypes | Catecholaminergic polymorphic ventricular tachycardia | RYR2 | AD | 1:237880625:GGGAC:- | - | 1982 | 27 | 0 | 16 | 0.993 | 0.007 | 0.007 | 27 | 0 | 1.344 | frameshift_variant | - | - | 3.4800000190734863 | - | - | - | - | - | LP | YES, a) |
| Genes related to cardiovascular phenotypes | Catecholaminergic polymorphic ventricular tachycardia | RYR2 | AD | 1:237880630:-:TTTCA | - | 1982 | 27 | 0 | 16 | 0.993 | 0.007 | 0.007 | 27 | 0 | 1.344 | frameshift_variant | - | - | 3.4800000190734863 | - | - | - | - | - | LP | YES, a) |
| Genes related to cardiovascular phenotypes | Dilated cardiomyopathy | TTN | AD | 2:179477760:AC:- | - | 2054 | 27 | 0 | 0 | 0.994 | 0.006 | 0.006 | 27 | 0 | 1.297 | frameshift_variant | - | - | -6.840000152587891 | - | - | - | - | - | LP | YES, a) |
| Genes related to cardiovascular phenotypes | Dilated cardiomyopathy | TTN | AD | 2:179477764:-:G | - | 2053 | 28 | 0 | 0 | 0.993 | 0.007 | 0.007 | 28 | 0 | 1.346 | frameshift_variant | - | - | 5.71999979019165 | - | - | - | - | - | LP | YES, a) |
| Genes related to cancer phenotypes | Hereditary breast and/or ovarian cancer | BRCA2 | AD | 13:32913768:AT:- | - | 1993 | 29 | 0 | 0 | 0.993 | 0.007 | 0.007 | 29 | 0 | 1.434 | frameshift_variant | - | - | -1.9600000381469727 | - | - | - | - | - | LP | YES, a) |
| Genes related to cancer phenotypes | Hereditary breast and/or ovarian cancer | BRCA2 | AD | 13:32913773:-:GT | - | 1993 | 29 | 0 | 0 | 0.993 | 0.007 | 0.007 | 29 | 0 | 1.434 | frameshift_variant | - | - | 4.050000190734863 | - | - | - | - | - | LP | YES, a) |
| Genes related to cardiovascular phenotypes | Arrhythmogenic right ventricular cardiomyopathy | DSP | AD | 6:7572230:-:CCTAC | - | 2052 | 29 | 0 | 0 | 0.993 | 0.007 | 0.007 | 29 | 0 | 1.394 | frameshift_variant | - | - | 4.679999828338623 | - | - | - | - | - | LP | YES, a) |
| Genes related to miscellaneous phenotypes | Wilson disease | ATP7B | AR | 13:52535985:A:C | rs121907998 | 2048 | 30 | 0 | 1 | 0.993 | 0.007 | 0.007 | 30 | 19 | 1.444 | missense_variant | 0.0009940357 | 0.004672897 | 1.659999966621399 | 0.44 | 0.008 | 14.09000015258789 | 3862 | Pathogenic/Likely pathogenic | P/LP | NO, b) |
| Genes related to cardiovascular phenotypes | Dilated cardiomyopathy | TTN | AD | 2:179477766:-:C | - | 2051 | 30 | 0 | 0 | 0.993 | 0.007 | 0.007 | 30 | 0 | 1.442 | frameshift_variant | - | - | 4.570000171661377 | - | - | - | - | - | LP | YES, a) |
| Genes related to cancer phenotypes | MUTYH-associated polyposis | MUTYH | AR | 1:45797228:C:T | rs36053993 | 1990 | 31 | 0 | 1 | 0.992 | 0.008 | 0.008 | 31 | 15 | 1.534 | missense_variant | 0.008946322 | 0.018691588 | 5.400000095367432 | 0.0 | 1.0 | 29.399999618530273 | 5294 | Pathogenic/Likely pathogenic | P/LP | NO, b) |
| Genes related to cardiovascular phenotypes | Dilated cardiomyopathy | TTN | AD | 2:179579271:CA:- | - | 2047 | 34 | 0 | 0 | 0.992 | 0.008 | 0.008 | 34 | 0 | 1.634 | frameshift_variant | - | - | 2.3399999141693115 | - | - | - | - | - | LP | YES, a) |
| Genes related to cardiovascular phenotypes | Dilated cardiomyopathy | TTN | AD | 2:179579276:-:AC | - | 2047 | 34 | 0 | 0 | 0.992 | 0.008 | 0.008 | 34 | 0 | 1.634 | frameshift_variant | - | - | -1.3600000143051147 | - | - | - | - | - | LP | YES, a) |
| Genes related to miscellaneous phenotypes | Hereditary hemochromatosis | HFE | AR | 6:26093141:G:A | rs1800562 | 1966 | 110 | 2 | 1 | 0.973 | 0.027 | 0.027 | 112 | 1 | 5.390 | missense_variant | 0.042743538 | 0.042056076 | 5.349999904632568 | 0.0 | 1.0 | 25.700000762939453 | 9 | Conflicting interpretations of p | P | YES, d) |
