## Supplementary Table 2 for "Unveiling the Landscape of Reportable Genetic Secondary Findings in the Spanish Population: A Comprehensive Analysis Using the Collaborative Spanish Variant Server Database"

**Supplementary Table 2. Number of pathogenic or likely pathogenic variants accumulated per gene in both aggregate when analyzed both aggregate or individual CSVS samples, or only individual samples**. Columns are: Phenotype, which contains general phenotype associated with the gene. Inheritance, which contains the inheritance mode for the disease (AD, autosomal dominant; AR, autosomal recessive; SD, semidominant; XL, X-linked). Gene, denotates the gene in which the variant is located. Nvariants_all, which lists the number of variants observed in both aggregate or individual samples. Nvariants_single, which lists the number of variants observed just in individual samples.

| Phenotype | Inheritance | Gene | Nvariants_all | Nvariants_single |
|---|---|---|---|---|
| Hypertrophic cardiomyopathy | AD | ACTC1 | 2 | 0 |
| Hereditary hemorrhagic telangiectasia | AD | ACVRL1 | 2 | 0 |
| Familial adenomatous polyposis | AD | APC | 6 | 2 |
| Familial hypercholesterolemia | AD | APOB | 6 | 1 |
| Wilson disease | AR | ATP7B | 8 | 3 |
| Dilated cardiomyopathy | AD | BAG3 | 3 | 1 |
| Hereditary breast and/or ovarian cancer | AD | BRCA1 | 20 | 0 |
| Hereditary breast and/or ovarian cancer | AD | BRCA2 | 66 | 2 |
| Biotinidase deficiency | AR | BTD | 9 | 5 |
| Malignant hyperthermia | AD | CACNA1S | 10 | 1 |
| Catecholaminergic polymorphic ventricular tachycardia | AR | CASQ2 | 1 | 0 |
| Ehlers–Danlos syndrome, vascular type | AD | COL3A1 | 7 | 1 |
| Dilated cardiomyopathy | AD | DES | 1 | 1 |
| Arrhythmogenic right ventricular cardiomyopathy | AD | DSC2 | 4 | 0 |
| Arrhythmogenic right ventricular cardiomyopathy | AD | DSG2 | 7 | 2 |
| Arrhythmogenic right ventricular cardiomyopathy | AD | DSP | 15 | 0 |
| Hereditary hemorrhagic telangiectasia | AD | ENG | 2 | 0 |
| Aortopathies | AD | FBN1 | 13 | 1 |
| Dilated cardiomyopathy | AD | FLNC | 4 | 1 |
| Pompe disease | AR | GAA | 5 | 4 |
| Fabry disease | XL | GLA | 1 | 1 |
| Hereditary hemochromatosis | AR | HFE | 1 | 1 |
| Maturity-onset diabetes of the young | AD | HNF1A | 1 | 0 |
| Long QT syndrome types 1 and 2 | AD | KCNH2 | 4 | 1 |

| | | | | |
|---|---|---|---|---|
| Long QT syndrome types 1 and 2 | AD | KCNQ1 | 5 | 1 |
| Familial hypercholesterolemia | AD | LDLR | 6 | 3 |
| Hereditary paraganglioma–pheochromocytoma syndrome | AD | MAX | 1 | 0 |
| Lynch syndrome | AD | MLH1 | 1 | 1 |
| Lynch syndrome | AD | MSH2 | 9 | 0 |
| Lynch syndrome | AD | MSH6 | 6 | 2 |
| MUTYH-associated polyposis | AR | MUTYH | 6 | 4 |
| Hypertrophic cardiomyopathy | AD | MYBPC3 | 6 | 2 |
| Aortopathies | AD | MYH11 | 4 | 0 |
| Hypertrophic cardiomyopathy | AD | MYH7 | 2 | 1 |
| Hypertrophic cardiomyopathy | AD | MYL2 | 2 | 0 |
| Hypertrophic cardiomyopathy | AD | MYL3 | 2 | 2 |
| Neurofibromatosis type 2 | AD | NF2 | 12 | 0 |
| Hereditary breast and/or ovarian cancer | AD | PALB2 | 7 | 0 |
| Familial hypercholesterolemia | AD | PCSK9 | 1 | 0 |
| Arrhythmogenic right ventricular cardiomyopathy | AD | PKP2 | 4 | 2 |
| Lynch syndrome | AD | PMS2 | 2 | 0 |
| Hypertrophic cardiomyopathy | AD | PRKAG2 | 1 | 0 |
| PTEN hamartoma tumor syndrome | AD | PTEN | 5 | 0 |
| Retinoblastoma | AD | RB1 | 8 | 0 |
| Dilated cardiomyopathy | AD | RBM20 | 6 | 3 |
| Familial medullary thyroid cancer | AD | RET | 10 | 0 |
| RPE65-related retinopathy | AR | RPE65 | 7 | 2 |
| Malignant hyperthermia | AD | RYR1 | 18 | 3 |
| Catecholaminergic polymorphic ventricular tachycardia | AD | RYR2 | 22 | 0 |
| Long QT syndrome 3; Brugada syndrome | AD | SCN5A | 1 | 1 |
| Hereditary paraganglioma–pheochromocytoma syndrome | AD | SDHAF2 | 2 | 0 |
| Hereditary paraganglioma–pheochromocytoma syndrome | AD | SDHB | 2 | 1 |
| Hereditary paraganglioma–pheochromocytoma syndrome | AD | SDHC | 3 | 1 |
| Aortopathies | AD | SMAD3 | 5 | 1 |
| Juvenile polyposis syndrome | AD | SMAD4 | 8 | 0 |
| Peutz–Jeghers syndrome | AD | STK11 | 2 | 0 |
| Aortopathies | AD | TGFBR2 | 3 | 0 |

| | | | | |
|---|---|---|---|---|
| Hereditary paraganglioma–pheochromocytoma syndrome | AD | TMEM127 | 1 | 0 |
| Arrhythmogenic right ventricular cardiomyopathy | AD | TMEM43 | 2 | 2 |
| Dilated cardiomyopathy | AD | TNNT2 | 2 | 0 |
| Li–Fraumeni syndrome | AD | TP53 | 2 | 0 |
| Catecholaminergic polymorphic ventricular tachycardia | AR | TRDN | 12 | 1 |
| Tuberous sclerosis complex | AD | TSC1 | 9 | 2 |
| Tuberous sclerosis complex | AD | TSC2 | 10 | 1 |
| Dilated cardiomyopathy | AD | TTN | 125 | 13 |
| Hereditary transthyretin-related amyloidosis | AD | TTR | 1 | 0 |
| TOTAL | | | 541 | 77 |
